# Polygenic scores for obstructive sleep apnea based on BMI-adjusted and -unadjusted genetic associations reveal pathways contributing to cardiovascular disease

**DOI:** 10.1101/2024.10.20.24315783

**Authors:** Nuzulul Kurniansyah, Satu J Strausz, Geetha Chittoor, Shreyash Gupta, Anne E Justice, Yana Hrytsenko, Brendan T Keenan, Brian E Cade, Brian W Spitzer, Heming Wang, Jennifer Huffman, Matthew R Moll, Bernhard Haring, Su Yon Jung, Laura M Raffield, Robert Kaplan, Jerome I Rotter, Stephen S Rich, Sina A Gharib, Traci M Bartz, Peter Y Liu, Han Chen, Myriam Fornage, Lifang Hou, Daniel Levy, Alanna Morrison, Heather M Ochs-Balcom, Bruce Psaty, Peter WF Wilson, Kelly Cho, Allan I Pack, Hanna M Ollila, Susan Redline, Daniel J Gottlieb, Tamar Sofer, FinnGen, Trans-Omics in Precision Medicine Consortium, VA Million Veteran Program

## Abstract

**Background:** Obstructive sleep apnea (OSA) is a heterogeneous disease, with obesity a significant risk factor via increased airway collapsibility, reduced lung volumes, and possibly body fat distribution.

**Methods:** Using race/ethnic diverse samples from the Million Veteran Program, FinnGen, TOPMed, All of Us (AoU), Geisinger’s MyCode, MGB Biobank, and the Human Phenotype Project (HPP), we developed, selected, and assessed polygenic scores (PGSs) for OSA, relying on genome-wide association studies both adjusted and unadjusted for BMI: BMIadjOSA- and BMIunadjOSA-PGS. We tested their associations with CVD in AoU.

**Results:** Adjusted odds ratios (ORs) for OSA per 1 standard deviation of the PGSs ranged from 1.38 to 2.75. The associations of BMIadjOSA- and BMIunadjOSA-PGSs with CVD outcomes in AoU shared both common and distinct patterns. For example, BMIunadjOSA-PGS was associated with type 2 diabetes, heart failure, and coronary artery disease, but the associations of BMIadjOSA-PGS with these outcomes were statistically insignificant with estimated OR close to 1. In contrast, both BMIadjOSA- and BMIunadjOSA-PGSs were associated with hypertension and stroke. Sex stratified analyses revealed that BMIadjOSA-PGS association with hypertension was driven by data from OR=1.1, p-value=0.002, OR=1.01 p-value=0.2 in males). OSA PGSs were also associated with dual-energy X-ray absorptiometry (DXA) body fat measures with some sex-specific associations.

**Conclusions:** Distinct components of OSA genetic risk are related to obesity and body fat distribution, and may influence clinical outcomes. These may explain differing OSA risks and associations with cardiometabolic morbidities between sex groups.

## Introduction

Obstructive sleep apnea (OSA) is a common disorder with an estimated prevalence of 17% (women) and 34% (men) in middle aged U.S. adults (1). OSA is associated with increased risk of cardiometabolic diseases, such as type 2 diabetes and hypertension, as well as increased rates of cardiovascular diseases (CVDs) including stroke, ischemic heart disease, and others (2–4). Despite recognition of the high prevalence of OSA, it remains underdiagnosed (5,6), with likely lower rates of diagnosis in women and in individuals who are less symptomatic (i.e. have less comorbidities such as hypertension or diabetes) (7). Major OSA risk factors include obesity, male sex, and older age (8).

Polygenic scores (PGSs) summarize genetic contributions to a trait as linear combinations of many trait-associated genetic alleles. PGSs are being increasingly developed and are being applied to assess genetic relationships between phenotypes (9–11) and identify changes in genetic contributions by age, lifestyle, and environment (12–15). Importantly, PGSs are being studied and sometimes translated into clinical use, such as risk prediction, stratification and classification into risk groups, and for screening (16,17). PGSs have also been used for characterizing etiologic pathways (18,19). PGSs for OSA may help advance the understanding of OSA given the heterogeneity of OSA phenotypes, its complex associations with other chronic diseases, and the incomplete understanding of its underlying mechanisms (20). Ultimately, OSA PGSs may lead to improved risk stratification, screening, and treatment that targets underlying disease mechanisms.

To develop and assess OSA PGSs, we used data from racially and ethnically diverse samples from multiple biobanks and cohort studies, including the Million Veteran Program (MVP, (21)), FinnGen (22), TOPMed, All of Us, Geisinger’s MyCode, MGB Biobank, and the Human Phenotype Project (HPP). Given the known sex differences in OSA (23), and the strong effect of obesity on OSA risk, we use sex stratified summary statistics from genome-wide association studies (GWAS) of OSA without BMI adjustment from three of these cohorts (the MVP, FinnGen, and Han Chinese population GWAS) and after adjusting for BMI for two cohorts (MVP and FinnGen) to develop predictive OSA PGSs. We then constructed the PGSs in other studies. Earlier epidemiological and genetic work has demonstrated that BMI is the strongest causal risk factor for OSA (24). Therefore, it is critical to understand how BMI contributes to comorbidities and relationships between OSA and CVD. Ultimately, this potentially allows us to understand BMI-dependent and BMI-independent cardiovascular consequences of OSA. To achieve these aims, we assessed the associations of the PGSs with OSA, OSA-related measures, sleep phenotypes, and cardio-pulmonary, metabolic and kidney disease outcomes, in multiple independents studies including individuals representing diverse populations.

## Results

A schematic overview of the study design is provided in Figure 1. We used GWAS summary statistics from three sources: the Million Veteran Program (MVP), FinnGen (Release 10), and an OSA GWAS of Han Chinese (Table 1). We applied three modern Bayesian shrinkage methods (LDPred2, PRS-CS, and PRS-CSx) to develop PGSs based on the above GWAS and meta-analysis combinations (ancestry-specific, multi-population, sex-specific), and further developed PGSs as combinations (sums) of other PGSs. Here we use the term “ancestry-specific” to refer to the PGSs, because they were developed based on GWAS summary statistics that were computed within population groups defined to be genetically similar (e.g. the Harmonized Ancestry and Race Ethnicity (HARE) groups of MVP (25,26)). When evaluating the PGSs on groups of individuals, we used the term “population”, because the populations used in data analysis were defined based on self-reported race/ethnicity, rather than based on genetic similarity. We used the Mass General Brigham (MGB) Biobank dataset to train PGS summation weights, the Trans-omics for Precision Medicine (TOPMed) dataset to assess PGS, and then data from All of Us (AoU), Geisinger, and HPP, to further study the selected PGSs in association with OSA and sleep-related measures, as well as other health outcomes.

**Figure 1.**
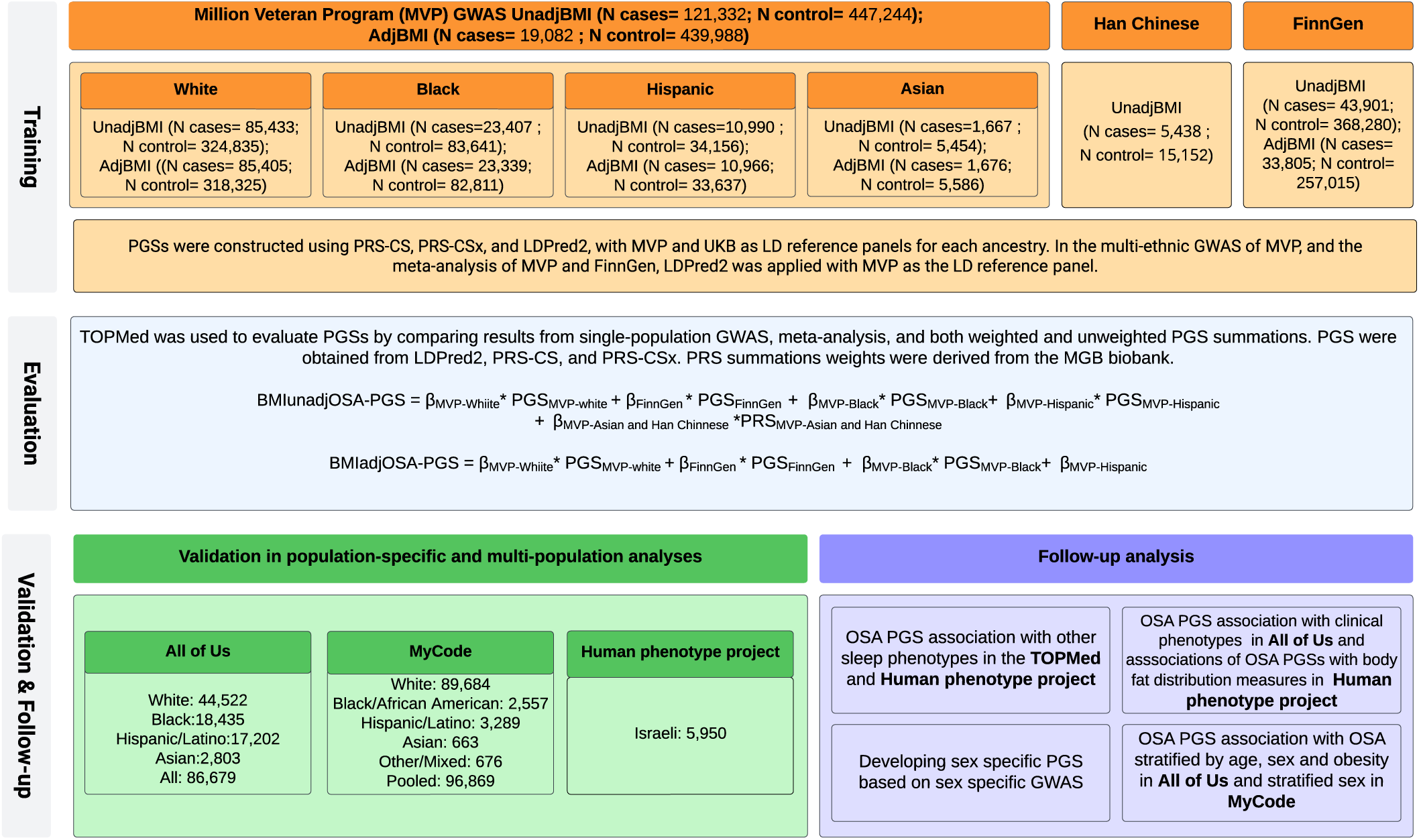
OSA PGS development and assessment. Development and assessment of OSA PGSs. The steps are composed of (a) PGS training using GWAS summary statistics, reference panels, and a separate population for computing PGS summation weights; (b) evaluation step used to select PGS out of multiple candidates; (c) validation of associations with OSA in new independent datasets; and (d) follow up analyses addressing OSA PGS associations with OSA within various strata, associations with related sleep phenotypes, comorbidities, and sequelae of OSA. Analyses in step (d) were in datasets from previous steps.

**Table 1:** GWAS summary statistics used for OSA PGS development.

| GWAS | Reference | Sex group | BMI adjustment | Sex-combined sample size | Study population |
| --- | --- | --- | --- | --- | --- |
| **MVP** | PMID 36989840 | Combined, and sex-stratified | Unadjusted | Cases: 121,332<br>Controls: 447,244 | 91% male, 72% White, 19% Black, 8% Hispanic, 1.2% Asian. |
| **MVP** | PMID 36989840 | Combined, and sex-stratified | Adjusted | Cases: 119,082<br>Controls: 439,988 | 91% male, 72% White, 19% Black, 8% Hispanic, 1.2% Asian. |
| **FinnGen Release 10** | PMID 36653562 | Combined, and sex-stratified | Adjusted | Cases: 33,805;<br>Controls: 257,015 | Finnish Europeans |
| **FinnGen Release 10** | PMID 36653562 | Combined, and sex-stratified | Unadjusted | Cases: 43,901;<br>Controls: 368,280 | Finnish Europeans |
| **Han Chinese** | PMID<br>35819321 | Combined | Unadjusted | Cases: 5,438;<br>Controls:15,152 | Han Chinese |

MVP study population was defined by “HARE group”: groups defined by a harmonized genetic ancestry and self-reported race/ethnicity algorithm (25) designed to create genetically similar groups, with group definitions relying on self-reported race/ethnicity of the majority of group members.

PGS software use summary statistics from a large number of variants, including correlated variants, and require a reference panel of linkage disequilibrium (LD) between the variants. Ideally, the LD reference would match the population from the GWAS corresponding to the summary statistics used. PRS-CS and PRS-CSx provide reference LD panels from the 1000 Genome and from the UK Biobank (UKB) studies. Because these may not adequately correspond to the MVP population, we developed LD reference panels using the MVP dataset. We used a multi-step procedure to assess PGSs where we (a) assessed whether UKB LD reference panel is sufficient when using PRS-CS (compared to using our developed MVP LD reference panel); (b) compared ancestry-specific PRS-CS and LDPred2 PGSs (where LDPred2 used the MVP reference panel); (c) compared PGSs based on multi-ethnic meta-analysis to weighted combination of population-specific PGSs (where the population-specific PGSs relied on methods selected in steps (a) and (b)).

### Evaluation of PGSs in TOPMed

The clinical characteristics of TOPMed participants are provided in Supplementary Table 1. Briefly, participants were self-identified from 4 race/ethnicity groups: White, Black, Hispanic/Latino, and Asian. Characteristics of individuals differed across these groups. For example, the average ages of participants were 68 years (Asian), 63 years (White), 56 years (Black), and 48 years (Hispanic/Latino). Most population groups had balanced sex ratio (50% female), while 58% of Hispanics/Latinos were female. Mean BMI was similar (~ 30 kg/m^2^) in the group of White, Black, and Hispanic/Latino individuals, but lower (~24 kg/m^2^) on average, in the group of Asian participants.

As described in Supplementary Note 1 and Supplementary Figures 1 and 2, using PRS-CS with the public UKB LD reference panel resulted in equivalent or better performance to PRS-CS using the MVP reference panel. Next, we compared LDPred2 (with MVP LD reference panel) to PRS-CS (with UKB reference panel) ancestry-specific PGSs. The results are provided in Supplementary Figures 3 and 4, demonstrating that the results were comparable for both approaches, except for White individuals; in this case, using the PRS-CS with UKB reference panel performed better. Therefore, for ancestry-specific PGSs we proceeded with PRS-CS with UKB LD reference panel. Next, we addressed the multi-ethnic PGS construction by comparing: (i) an LDPred2 PGS based on the multi-ethnic meta-analysis of all GWAS with the multi-population MVP reference panel, multi-PGS summation of either standardized PRS-CS or PRS-CSx ancestry-specific PGSs using either (ii) MGB-inferred weights, or (iii) equal weights. As shown in Supplementary Figures 5 and 6, the best performing PGSs were MGB-inferred weighted combination of PRS-CS PGSs. Characteristics of MGB participants are provided in Supplementary Table 2. Here, the PGS based on BMI-adjusted GWAS, which we dub BMIadjOSA PGS, is the combination is of 5 PGSs, based on GWAS of: MVP White group, FinnGen, MVP Black group, MVP Hispanic/Latino group. The final PGS based on BMI-unadjusted GWAS, called BMIunadjOSA PGS, is a weighted summation of 6 PGSs, based on the BMI-unadjusted versions of the above GWAS, and further including a PGS developed based on MVP Asian group meta-analyzed with the Han Chinese OSA GWAS. The two final PGSs (BMIadjOSA and BMIunadjOSA PGS) were highly correlated: Pearson correlation was 0.96, computed over the TOPMed dataset. Mean and standard deviations (SDs) of these PGSs computed over the TOPMed datasets are provided in Supplementary Table 3.

Figure 2 compares the associations of the selected BMIadjOSA-PGS and BMIunadjOSA-PGS with OSA in TOPMed individuals, stratified by self-reported race/ethnicity, age, BMI, and sex. Here, OSA was defined using the apnea hypopnea index (AHI) or the respiratory event index (REI), depending on availability of the measurement, or self-reported diagnosis of OSA (when overnight sleep study was not available, see Supplementary Table 4 for OSA definition by study). When using AHI or REI, OSA was defined as AHI/REI≥ 15. The association models were adjusted for self-reported race/ethnicity, age, sex, BMI (both linear and squared terms), and the first 11 PCs of genetic ancestry. The OSA associations of the BMIadjOSA-PGS are stronger than the association of the BMIunadjOSA-PGS. In raw association, presented as proportions of individuals in OSA categories (no OSA, mild, moderate, and severe OSA) within quintiles of the PGS, both PGSs result in similar patterns (Figure 2b). Due to the differences in PGS distribution between self-reported race/ethnicity groups (Figure 2a), group-combined raw (quintile-based) associations are not appropriate; however, the PGS associations with OSA are very consistent across different race/ethnicity, sex, and age strata in covariate-adjusted models (Figure 2c).

**Figure 2:**
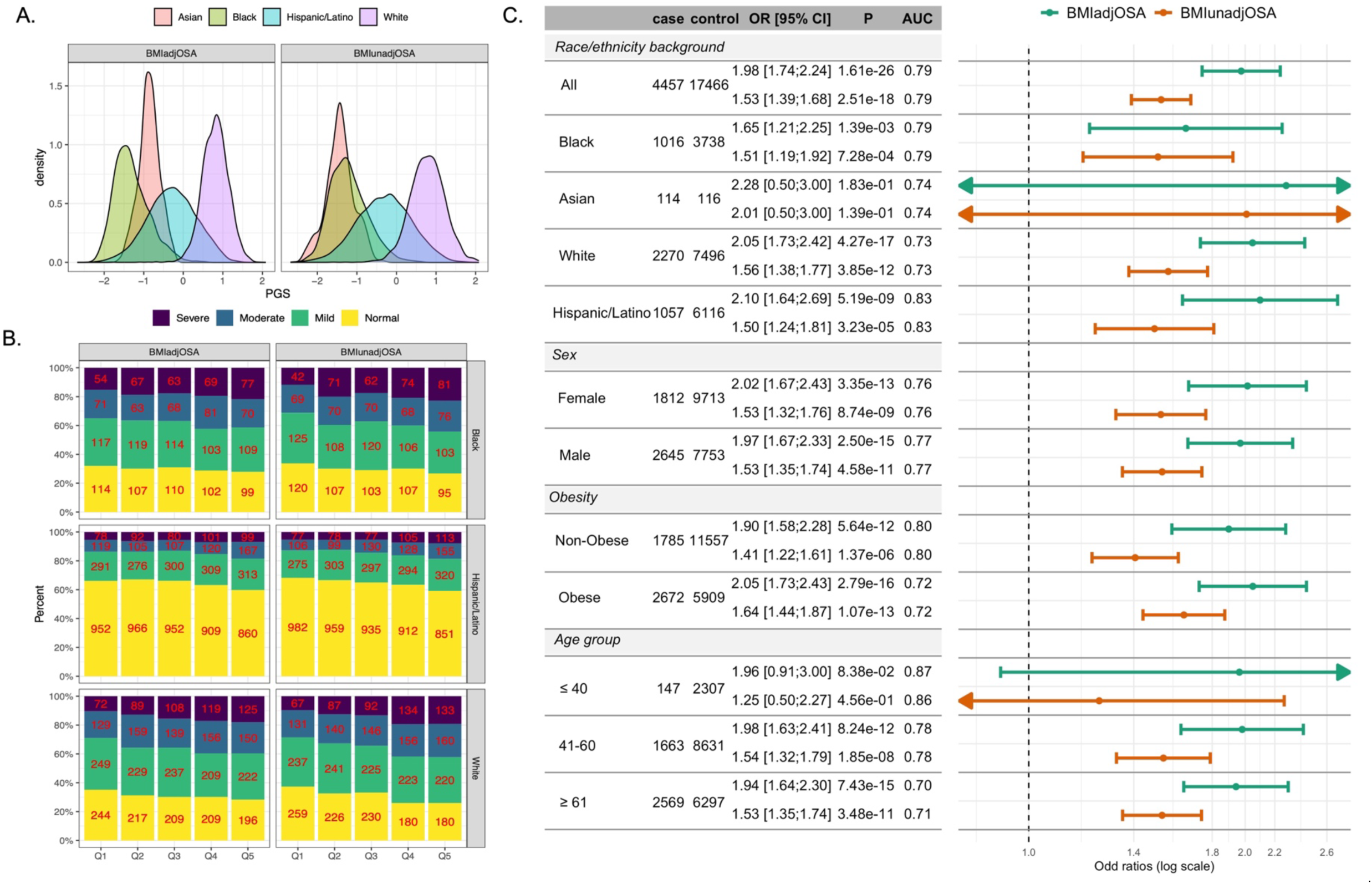
OSA PGS associations with OSA in TOPMed individuals. Panel a: distributions of BMIadjOSA and BMIunadjOSA PGSs in strata defined by self-reported race/ethnicity. Panel b: precents and numbers of individuals with normal, mild, moderate, and severe OSA (defined by cut-points of REI/AHI or 5, 15, and 30), in quintiles of the OSA PGSs by self-reported race/ethnicity (Asian group excluded due to low sample size). Panel c: estimated OSA PGS associations with OSA in TOPMed combined and stratified samples. Analyses were adjusted for age, sex (unless sex-stratified), self-reported race/ethnicity (unless stratified by that), and BMI (both linear and squared terms). AHI: apnea hypopnea index; OSA: obstructive sleep apnea; PGS: polygenic score; REI: respiratory event index.

In TOPMed individuals who had OSA and OSA-related phenotypes measured overnight sleep study available, analyses using both BMIadjOSA- and BMIunadjOSA-PGSs (Figure 3) were generated using participants whose phenotypic characteristics are reported in Supplementary Tables 5 and 6. The PGSs are generally associated with OSA phenotypes (AHI, hypoxic burden, percent sleep time with oxyhemoglobin saturation below 90%, minimum and average oxygen saturation, and respiratory event-related oxygen desaturation during sleep; all log transformed), and are in the expected direction of effect. There are differences, however, in the trait associations between REM and NREM sleep, with stronger associations in phenotypes measured during NREM sleep (e.g., AHI: 0.15, 95% CI [0.12, 0.18]; NREM AHI: 0.16, 95% CI [0.11, 0.21]; REM AHI: 0.06, 95% CI [0.02, 0.11]). In addition, the PGS association with average oxyhemoglobin saturation during sleep (Avg SatO2) is very weak (effect size: −0.0003, 95% CI overlapping with zero, p-value =0.09). To address the hypothesis that this result is because Ave SatO2 potentially reflects lung function better than OSA, we estimated the association of lung function (FEV1 to FVC ratio) PGS with Avg SatO2 as well as with AHI, both log transformed. The lung function PGS was associated with Avg SatO2 (effect size: −0.0002, 95% CI: [−0.003,−0.00007], p-value=9.0 x10^-4^) but only weakly with AHI (effect size: 0.01, 95% CI: [0.002, 0.2], p-value=0.02, Supplementary Table 7).

**Figure 3.**
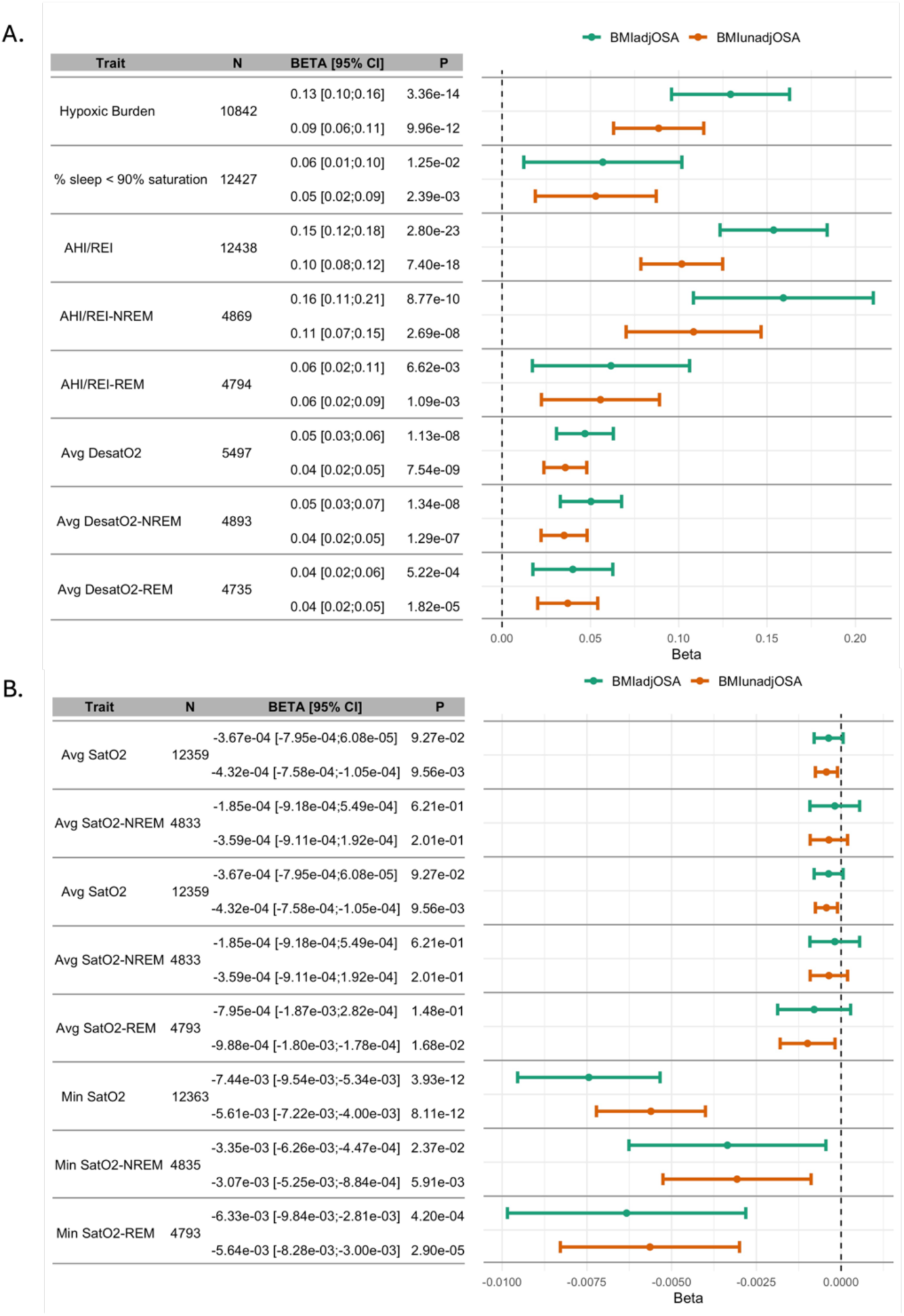
Estimated associations of OSA PGSs with OSA- and sleep-related phenotypes in TOPMed. Results from association analyses of OSA PGSs with OSA-related sleep measures. Panel a: measures that tend to be higher with more severe OSA, panel b: measures that tend to be lower with more severe OSA. AHI: apnea hypopnea index; NREM: non-rapid eye movement sleep; REI: respiratory event index; REM: rapid eye movement sleep; Avg SatO2: average oxyhemoglobin saturation during sleep; Min SatO2: minimum oxyhemoglobin saturation during sleep; Avg DesatO2: average oxyhemoglobin desaturation during respiratory events.

Sex specific PGSs (based on sex-specific GWAS) did not exhibit better performance than PGSs based on sex-combined analysis (Supplementary Figure 7 and 8).

### Associations of OSA PGSs in the Geisinger’s MyCode project

The MyCode project include individuals recruited from the Geinsinger healthcare provider. OSA categorization is based on ICD codes, as described in the Methods section. Figure 4 provides results from association analyses of BMIadjOSA-PGS and BMIunadjOSA-PGS in MyCode in a sex-combined analysis. Results from sex-stratified analysis are provided in Supplementary Figure 9. Characteristics of the study population are provided in Supplementary Table 8. There were 96,869 individuals, of which 19,148 (19.8%) had OSA. The mean age was 48 years, and 60% were female. Majority of individuals were White. The OSA PGSs were highly associated with OSA: BMIadjOSA-PGS overall OR=2.00, 95% CI:[1.89;2.12], in females OR=1.89, 95% CI:[1.75;2.05], and in males OR=2.14, 95% CI:[1.97;2.23].

**Figure 4:**
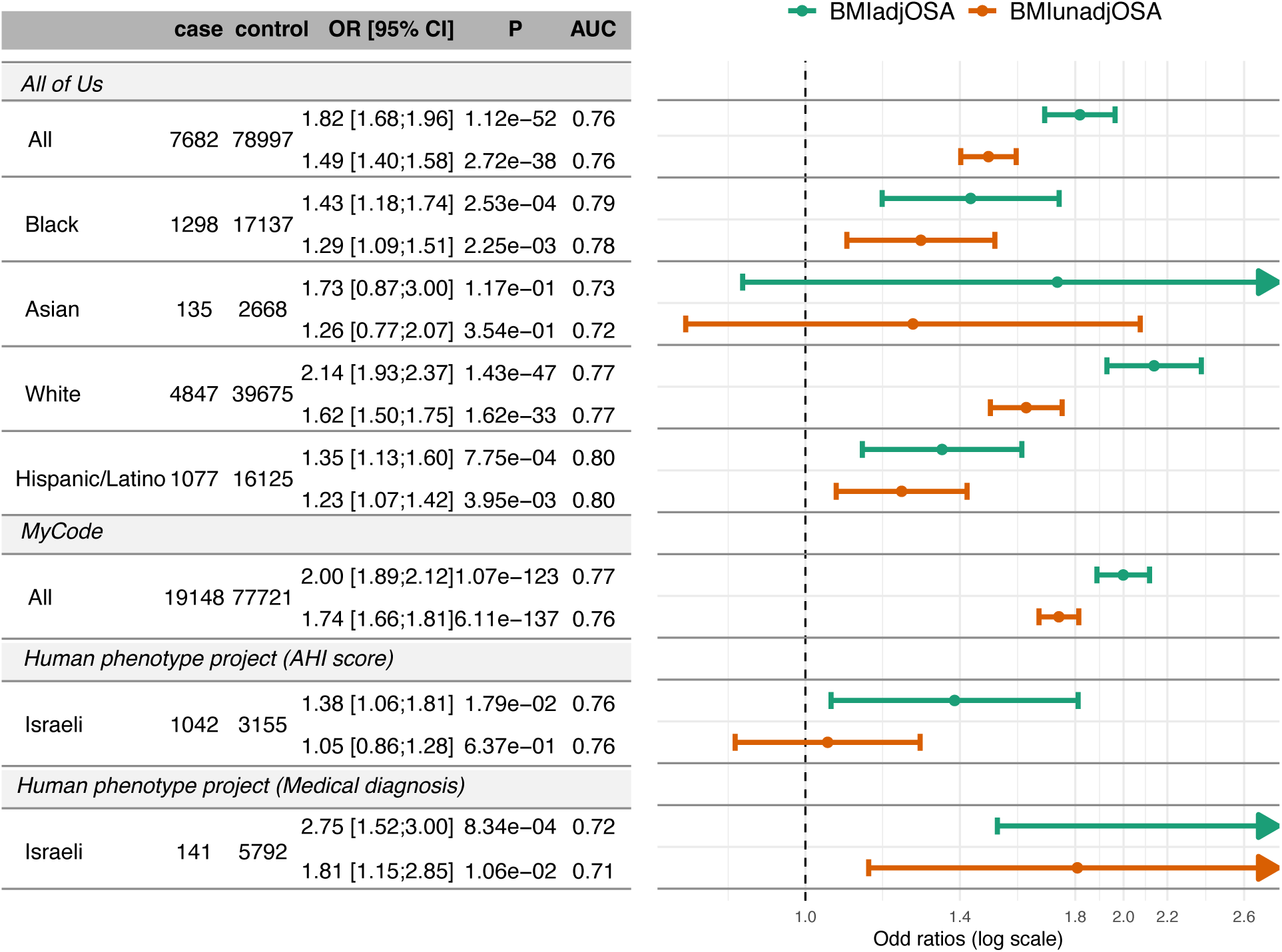
OSA PGS associations with OSA in validation studies. Estimated associations of BMIadjOSA and BMIunadjOSA PGSs with OSA in three validation datasets: All of Us, Geinsinger health system, and the Human Phenotype Project, in combined and stratified analyses.

### Associations of OSA PGSs with OSA phenotypes in the Human Phenotype Project

Supplementary Table 9 characterizes the participants from the HPP. There were 3,070 female and 2,880 male participants, ~52 years old on average, and the average BMI was ~26 kg/m^2^. Very few individuals had OSA according to medical diagnosis: 0.75% of female and 4.1% of male participants. Supplementary Table 10 further characterizes the subset of individuals who had sleep monitoring data measured using the WatchPAT device, and their objective sleep phenotypes (averaged values over three nights). Based on sleep monitoring data, 17.2% of female and 30.7% of male individuals had OSA defined as AHI≥ 15. BMIadjOSA-PGS was strongly associated with OSA in the HPP (Figure 4). The association was stronger when OSA was based on medical diagnosis (141 cases, 5,792 controls; OR=2.75), compared to the association when OSA was defined based on a 3-day average of AHI≥ 15 (1,042 cases, 3,155 controls; OR=1.38). Unlike the analysis in the TOPMed data, associations of quantitative OSA-related phenotypes with the OSA PGSs were similar when stratified by REM and NREM stage (Figure 5). PGS associations were stronger with the oxygen desaturation index (ODI) compared to the AHI or respiratory disturbance index (RDI). We also estimated associations with oxygen saturation phenotypes (Supplementary Figure 10), sleep stage duration (percentages of total sleep duration) (Supplementary Figure 11), and with self-reported sleep phenotypes (Supplementary Figure 12). OSA PGSs were strongly associated with lower minimum and average oxygen saturation during sleep, with stronger associations for BMIadjOSA-PGS compared to BMIunadjOSA-PGS. The PGS associations with other phenotypes were generally weaker. Among the associations that had p-value<0.05, BMIunadjOSA-PGS was associated with less sleep time percentage in light sleep and longer percentage of time in REM sleep. BMIadjOSA-PGS was weakly associate with higher likelihood of daytime sleepiness (p-value=0.04) and BMIunadjOSA-PGS was associated with both shorter sleep duration (beta from association with continuous sleep hours question =-0.07, p-value=0.03), and with lower likelihood of long (>9 hours) sleep duration (OR=0.35 ≤9 hours, p-value=0.04). Characteristics of self-reported sleep phenotypes among the HPP participants are provided in Supplementary Table 11.

**Figure 5:**
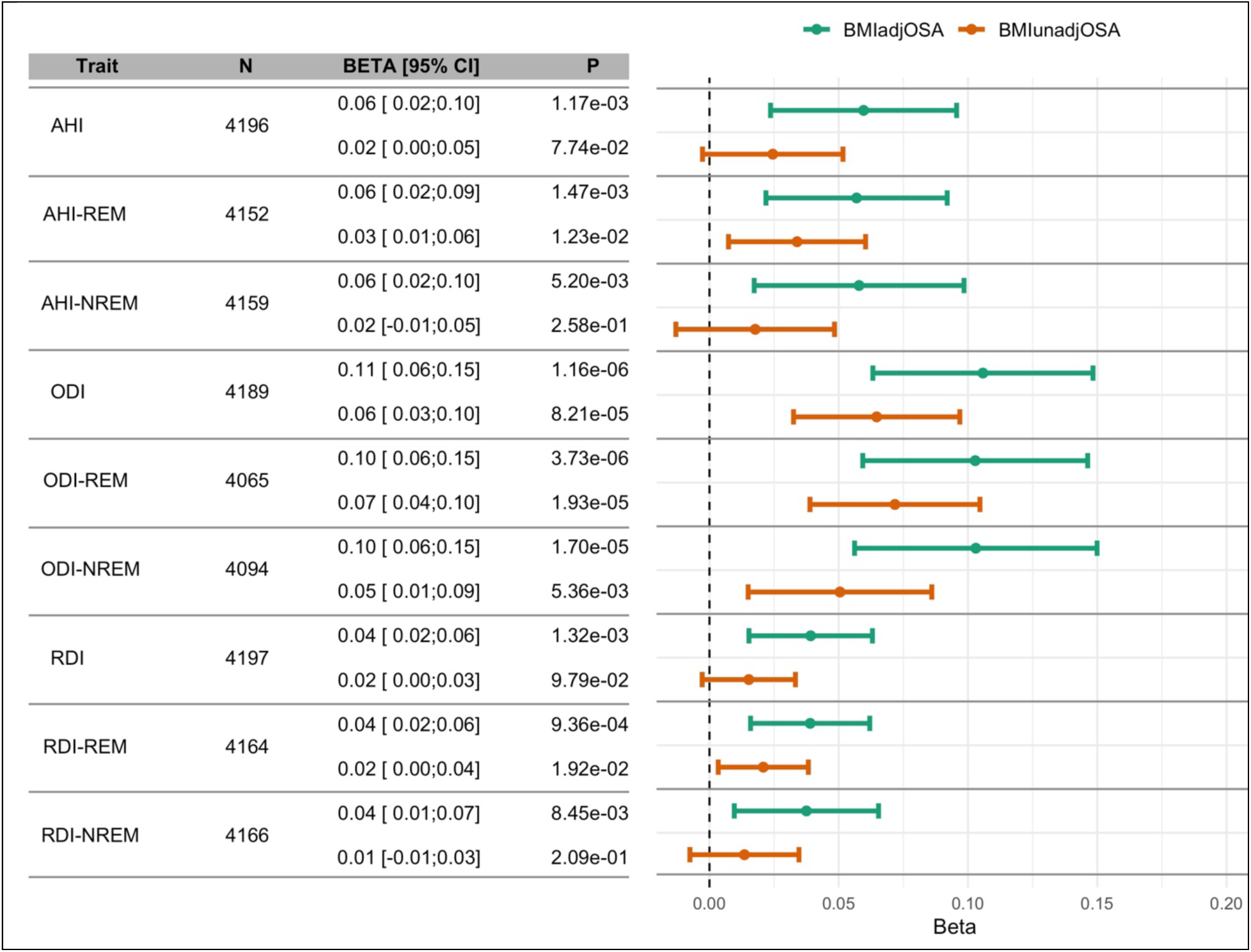
OSA PGS associations with quantitative OSA phenotypes in the Human Phenotype Project. Estimated associations of BMIadjOSA and BMIunadjOSA PGSs with sleep monitoring, log-transformed OSA-related measures in the HPP, stratified by REM and NREM sleep and combined. All measures were averaged over the three nights of sleep monitoring. AHI: apnea hypopnea index; HPP: Human Phenotype Project; ODI: oxygen desaturation index; RDI: respiratory disturbance index; REM: rapid eye movement; NREM: non-REM.

### Associations of OSA PGSs with clinical outcomes in the All of Us study

All of Us participant characteristics are provided in Supplementary Table 12. Between 6%-15% of participants were classified with OSA, with the lowest and highest percentages observed in self-reported Asian self-reported White participants, respectively. All association analyses were adjusted for age, BMI (linear and squared terms), sex at birth, self-reported race/ethnic background, and the first 10 genetic PCs. First, we performed OSA association analysis in the combined cohort with BMIadjOSA PGS, BMIunadjOSA PGS, as well as with other previously reported PGSs that used either only genome-wide significant SNPs (21), or used the clump & threshold method to derive PGS based on BMI-unadjusted analysis using FinnGen-only data (10). The results are provided in Supplementary Table 13, demonstrating that the newly-developed OSA PGSs have stronger associations with OSA compared to previously-developed one. Figure 4 visualizes the associations of the developed OSA PGSs with OSA in All of Us (as well as other cohorts), and Supplementary Figure 9 provides results stratified by sex, age, and obesity groups. Unlike in TOPMed, the OSA PGS association estimates in All of Us differed across self-reported race/ethnicity groups, with the association in self-reported White individuals being the strongest (OR=2.14 of BMIadjOSA-PGS, 95% CI [1.93, 2.37]), and weakest in self-reported Hispanic/Latino individuals (BMIadjOSA-PGS OR= 1.35, 95% CI [1.13, 1.60]).

Next, we performed association analyses of BMIadjOSA-PGS and BMIunadjOSA-PGS with clinical outcomes potentially related to OSA: hypertension, type 2 diabetes, stroke, atrial fibrillation, atrial fibrillation, heart failure, coronary artery disease, asthma, chronic obstructive pulmonary disease (COPD), and chronic kidney disease. The results are visualized in Figure 6. BMIunadjOSA-PGS was associated with all clinical cardiovascular, pulmonary, and kidney outcomes, while BMIadjOSA-PGS had more variable, and usually weaker, associations. Notably, BMIadjOSA-PGS was not associated with type 2 diabetes, heart failure, coronary artery disease, COPD, or chronic kidney disease, but associated with hypertension, stroke, atrial fibrillation, and asthma (raw p-value<0.05).

**Figure 6:**
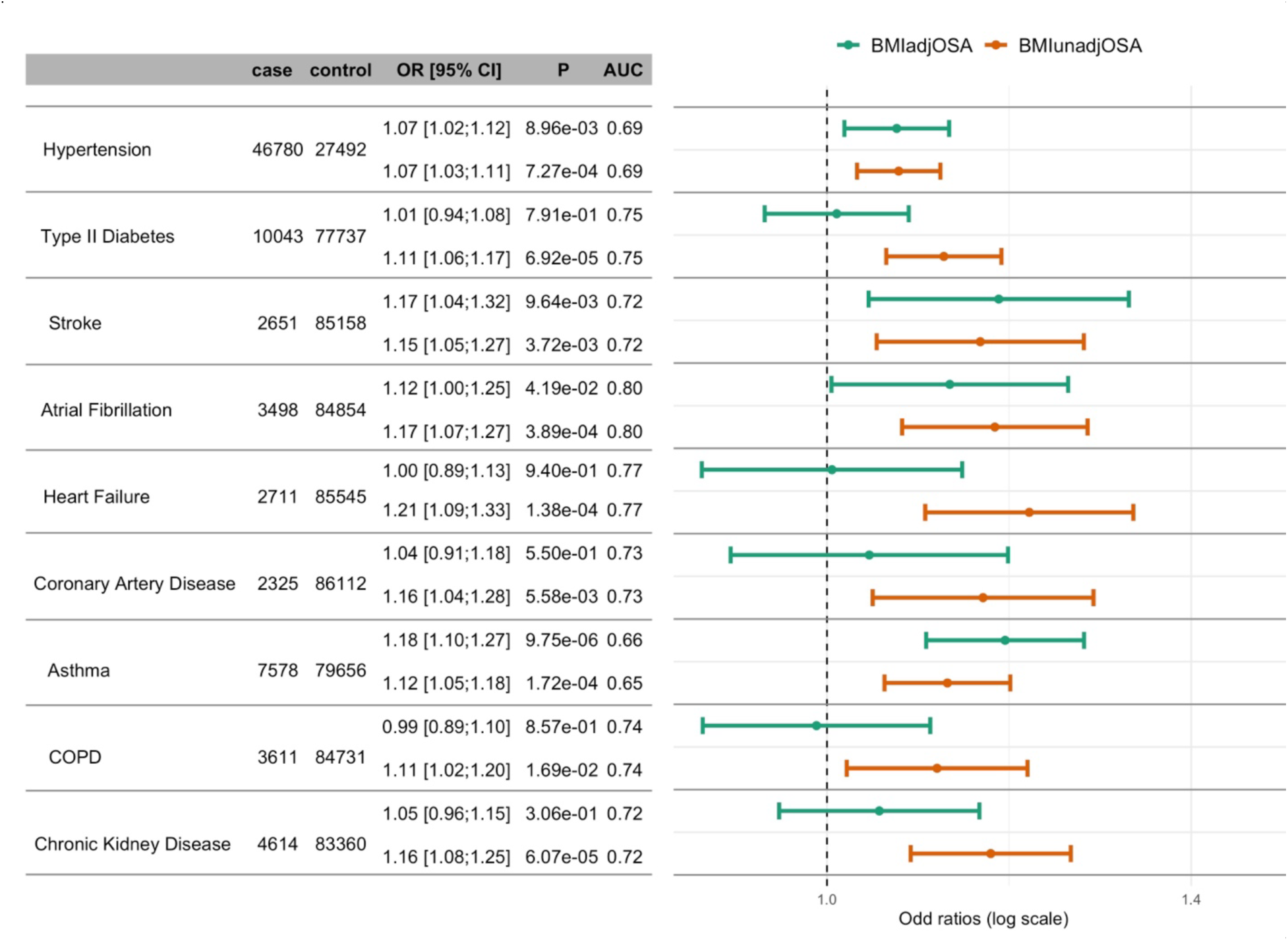
OSA PGS associations with other clinical outcomes in All of Us. Estimated associations of BMIadjOSA and BMIunadjOSA PGSs with clinical outcomes in the All of Us study. Associations were adjusted for age, sex, BMI (linear and squared terms), and 10 PCs of genetic ancestry.

Figure 7 (hypertension and type 2 diabetes) and Supplementary Figure 13 (all other outcomes) provide results from sex-stratified association analyses of OSA PGSs. The associations with hypertension and diabetes suggest sex-differences: Both BMIadjOSA- and BMIunadjOSA-PGS associations with hypertension were driven by the female stratum, while the BMIunadjOSA-PGS association with T2D was driven by the male stratum.

**Figure 7:**
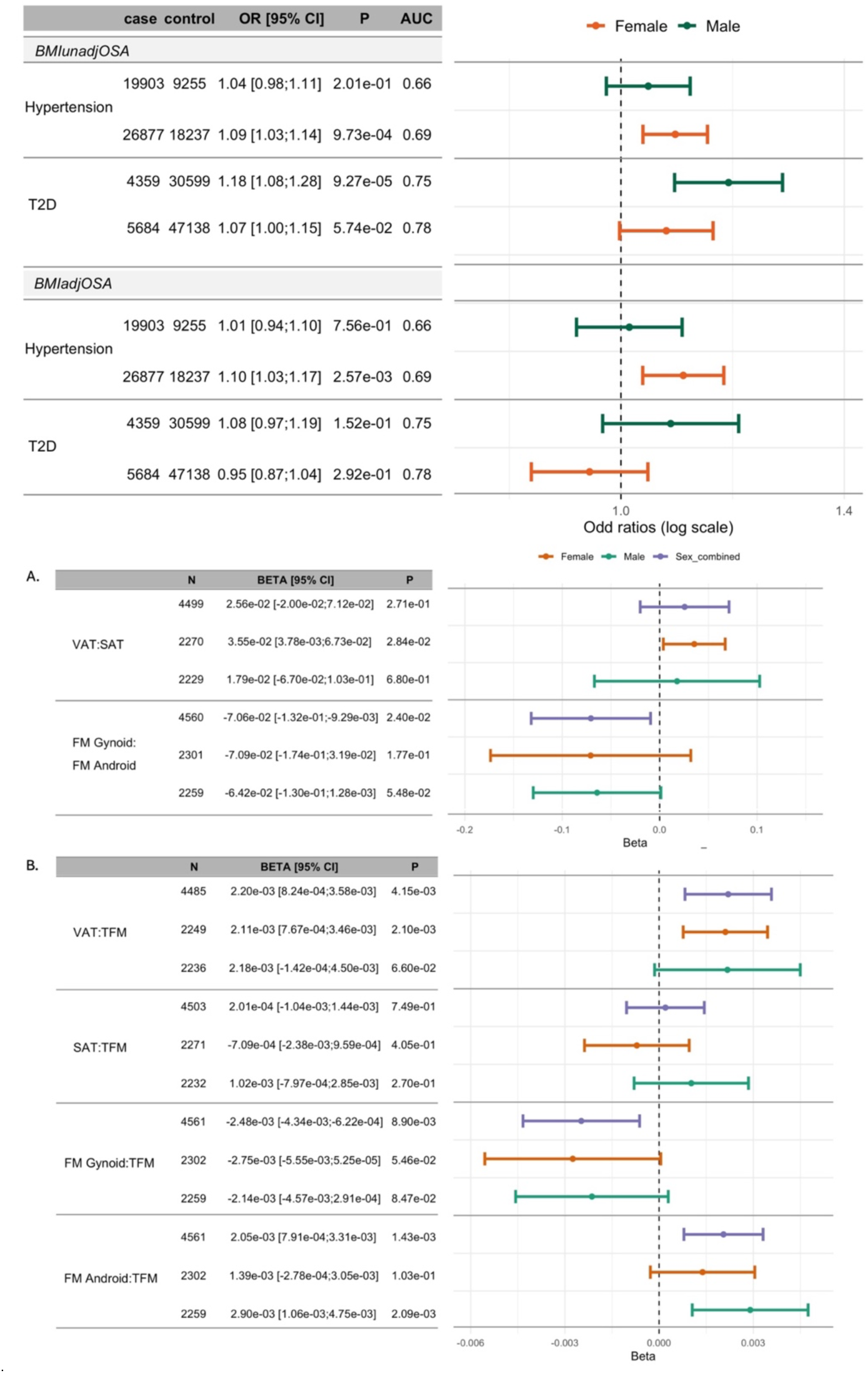
Sex-stratified associations OSA PGSs with cardiometabolic traits and body fat distribution measures from DXA scan. Top: Estimated adjusted odds ratio of OSA PGSs with hypertension and T2D in All of Us. Bottom: estimated associations of BMIadjOSA-PGS with DXA measures. All associations were adjusted for age, sex, BMI (linear and squared terms), and the 10 first genetic principal components. Results from association analyses that used BMIunadjOSA-PGS and was not adjusted for BMI are provided in Supplementary Figure 14. BMI: body mass index; OR: odds ratio; VAT: visceral adipose tissue; SAT: subcutaneous adipose tissue; FM: fat mass; TFM: total scan fat mass; T2D: type 2 diabetes.

### Associations of OSA PGSs with body fat distribution measures from DXA scan

We used data from the HPP to study whether BMIadjOSA and BMIunadjOSA PGSs are associated with body fat distribution measures to potentially explain their different associations with health outcomes. We considered measures based on total body visceral adipose tissue (VAT), subcutaneous adipose tissue (SAT), gynoid fat mass, and android fat masses. Specifically, we used, as outcomes, the proportions of total body VAT and SAT mass out of total body fat mass (TFM), the ratio VAT:SAT mass, the proportions of gynoid and android fat out of TFM, and the ratio gynoid:android fat mass. Characteristics of these DXA phenotypes are provided in Supplementary Table 14. Association analyses results are provided in Figure 7 (BMI adjusted analyses) and Supplementary Figure 14 (BMI unadjusted analyses). In combined sex association analyses that was not adjusted to BMI, BMIunadOSA-PGS was associated with all DXA outcomes. Higher BMIunadjOSA-PGS values were strongly associated with higher proportions of VAT and SAT mass proportions, and with higher android mass proportion but with lower gynoid mass proportion out of TFM. Also, it was associated with increased VAT:SAT ratio, and with lower gynoid:android mass ratio. In BMI-adjusted analyses, results were similar, however, BMIadjOSA-PGS was not associated with SAT ratio out of TFM. Further, BMIadjOSA-PGS was associated with VAT:SAT only in the female stratum (Figure 7).

## Discussion

This is the largest study to date leveraging polygenic scores across global populations to investigate the role of BMI in OSA and related clinical outcomes. Associations of the BMIadjOSA-PGS with OSA, in BMI-adjusted analyses, were strong, and ranged from ORs of 1.38 from device-measured AHI≥ 15 and 2.75 from medical diagnosis-based OSA (HPP), to 1.98 (All of Us), and 2.00 (MyCode), likely reflecting a more severe phenotype that triggered clinical recognition. Importantly, associations differed between PGS developed based on BMI-adjusted and BMI-unadjusted GWAS. Using data available from the All of Us cohort, we further showed that the strength of associations with cardiovascular, metabolic, pulmonary and kidney disorders often differed depending on the BMI adjustment of the source OSA GWAS. For example, type 2 diabetes, coronary artery disease, and heart failure associations were null in association with BMIadjOSA-PGS but were positively associated BMIunadjOSA-PGS, suggesting that much of the co-aggregation of those cardio-metabolic phenotypes may be explained by genetic risk factors associated with obesity. In contrast, both BMIadjOSA- and BMIunadjOSA-PGSs were associated with hypertension, stroke, and asthma, and stroke, suggesting that obesity-independent risk factors for OSA are associated with those outcomes. Interestingly, epidemiological studies that adjusted for BMI and other cardiovascular confounders have generally shown more consistent associations with OSA for hypertension and stroke in comparison to coronary artery disease (27,28). These results point to a unique pathophysiology of OSA captured only by BMI-adjusted GWAS that may not be associated with elevated BMI and may specifically increase risk for vascular and airway diseases.

The association between asthma and BMIadjOSA-PGS is of interest given the reported associations between OSA and asthma phenotypes but uncertainty regarding the causal directionality of the relationship between OSA and asthma (29,30). Common risk factors for both disorders include not only obesity but also inflammation and atopy. Our study findings suggest that additional OSA-related risk factors, beyond obesity, may influence risk of asthma.

Obesity is one of the strongest risk factors for OSA and is independently an important risk factor for many other health outcomes including diabetes and various cardiovascular outcomes. Studies assessing the effect of OSA, and the effect of OSA treatment, on cardiovascular outcomes typically attempt to account for the role of BMI or obesity and evaluate the OSA-specific effect (31–33). Hence it is important to account for BMI in the development of OSA PGSs. The developed PGSs, BMIadjOSA- and BMIunadjOSA-PGSs are highly correlated, yet have different associations with OSA and with other health outcomes. The BMIadjOSA-PGS has stronger association with OSA in BMI-adjusted analysis, while the BMIunadjOSA-PGS has stronger association with other outcomes, likely due to it capturing BMI-related OSA genetic effects. Both PGSs are useful. It would be incorrect to dismiss the BMIunadjOSA-PGS as capturing primarily obesity rather than OSA genetics, because the impact of obesity is being mediated through OSA. BMIadjOSA-PGS captures OSA genetic effects without the contribution of obesity, but it is difficult to assess how effective this adjustment is (i.e., does some of the obesity-related OSA genetics remain captured in BMIadjOSA-PGS?), the observed null association of BMIadjOSA-PGS with diabetes in All of Us suggests that it was largely effective. We studied the associations of OSA-related phenotypes with BMIadjOSA- and BMIunadjOSA-PGSs in both TOPMed and HPP, both while adjusting the associations to BMI. In TOPMed, the associations of oxygen saturation phenotypes were stronger with BMIunadjOSA-PGS compared to BMIadjOSA-PGS (but this was not replicated in HPP). More work is needed to assess whether this points to OSA-specific obesity-related pathophysiology, such as obesity-hypoxemia interactions (30).

We performed association analyses of OSA PGSs with DXA scan measures in the HPP. While it would have been ideal to consider fat measures separately in the neck/upper airway area (34,35) and in the abdomen (36), we had overall VAT and SAT mass measures, and gynoid and android fat masses (not limited to VAT or SAT tissues). OSA PGSs were associated with higher VAT:SAT ratio, but the associations were statistically significant only in females (Figure 7). This is consistent with sex differences in cardiometabolic risk conferred by VAT (37) identified in the Framingham Heart Study. Higher VAT is associated with increased inflammation and insulin resistance (38,39), and increased insulin levels are in turn associated with OSA—even after adjusting for BMI (40). Insulin levels may be linked to OSA via the association of VAT with upper airway size and function (41). The association of OSA PGSs with VAT:SAT ratio, even in BMI-adjusted analysis, is consistent with the role of inflammation and insulin resistance in OSA. The stronger association observed in women also suggests a potential sex-specific mechanism.

Strengths of this study include the use of multiple independent and population-diverse datasets, with populations reflecting the diversity of the U.S. population, as well as a few world-wide populations. Our study is unique in that we employed both BMI-adjusted and -unadjusted GWAS to develop and assess PGSs, assessment of OSA associations in a stratified manner, and estimated the associations of OSA PGSs with multiple clinical measures across available datasets. Our results suggest areas for improvement in future studies. Importantly, there remains the challenge posed by the underdiagnosis of OSA (42), limiting results from GWAS as well as estimations of associations (43). For example, the OSA PGSs effect size estimates demonstrated unprecedented consistency across strata defined by age, sex, and self-reported race/ethnicity (unlike previous results with blood pressure PGS (15)) in TOPMed, where most of the cohorts assessed OSA using objective devices. In contrast, associations differed by self-reported race/ethnicity in All of Us, where OSA status was determined by electronic health record codes. There, the associations were strongest in self-reported White populations. Well-known health disparities between White and minority populations in the U.S. (44), suggest the possibility of referral bias and missed diagnosis of OSA in race/ethnic minority populations in the U.S. Such differences may underlie the observed differences in OSA PGS effect estimates.

In summary, we developed BMIadjOSA- and BMIunadjOSA-PGSs, which were highly associated with OSA across multiple, diverse, populations. Follow up association analyses in All of Us revealed the genetically determined, OSA-specific, obesity independent, association with hypertension, stroke, and asthma. The results support the importance of both obesity related and unrelated genetic risk factors for OSA.

## Online Methods

Figure 1 provides an overview of the study. In brief, we used GWAS summary statistics from multiple published GWAS of OSA using three modern methods, LDPred2 (45), PRS-CS (46), and PRS-CSx (47), that have been shown to be very successful for polygenic, complex, traits. We developed PGS after either meta-analyzing GWAS summary statistics across independent cohorts, or using a PGS combination approach, as a weighted sum. We then evaluated PGS in association with OSA, and performed additional analyses where we studied PGS associations with other phenotypes.

### GWAS summary statistics

We developed PGS based on summary statistics from MVP (21), from the FinnGen study (22), and from a GWAS of OSA in Han Chinese (48). Table 1 provides information about the summary statistics.

### GWAS of OSA in FinnGen

The FinnGen study is a large-scale genomics initiative that has analyzed over 500,000 Finnish biobank samples and correlated genetic variation with health data to understand disease mechanisms and predispositions. The project is a collaboration between research organizations and biobanks within Finland and international industry partners.

The project aims to identify the impact of genetic and environmental factors on various diseases while enhancing understanding of the progression and biological mechanisms underlying these conditions (49). Genome-wide association testing for OSA was performed utilizing Regenie 2.2.4 (50) through the FinnGen Regenie pipelines (https://github.com/FINNGEN/regenie-pipelines). The analysis was adjusted for the current age or age at death, sex, genotyping chip, genetic relatedness, and the first 10 genetic PCs. The analysis was also conducted adjusting for BMI, and it was repeated for both sexes separately. The baseline characteristics of the participants are presented in Supplementary Table 15.

### Creating an MVP LD reference panel for PGS development

We used three popular methods to develop PGSs: LDPred2, PRS-CS, and PRS-CSx, all requiring the use of reference panels – correlation matrices tabulating the linkage disequilibrium (LD; correlation) between single nucleotide polymorphisms (SNPs) used for PGS preparation. We constructed such matrices based on the MVP dataset, using unrelated individuals only (overall N=567,748).

We focused on HapMap SNPs, and further required, for each LD reference panel representing a specific population subset, MAF of at least 0.01, and imputation quality≥0.8. We excluded SNPs from the major histocompatibility complex (MHC) region (chr6: 26–34Mb; grch37),

We first used the R package bigsnpr (V1.12.) to create correlation matrices based on each of MVP HARE groups, and based on the combined sample. For the European HARE group, we subset the dataset to 100K unrelated individuals, to speed up computing time. Because other HARE groups had smaller sample sizes, we used all unrelated individuals associated with the group (N African: 106,445, N Hispanic: 44,714, N Asian: 7,149). To create the multi-population correlation matrix we sampled 100K individuals at random from the unrelated set of MVP individuals. Random sampling ensured that the proportions of individuals from each HARE group was maintained from the complete dataset. LDPred2 requires correlation matrices based on entire chromosomes. Thus, we used function snp_cor from bigsnpr to compute these matrices for each chromosome, HARE group, and the multi-population dataset.

PRS-CS and PRS-CSx required splitting each chromosome to smaller LD blocks, typically of hundreds of SNPs (46) due to computational limitations. Thus, we applied the snp_ldsplit function from the form bigsnpr package to split the correlation matrices from each chromosome, in each HARE group dataset, to LD blocks, varying the r^2^ parameters and minimum and maximum block sizes as needed (following the bigsnpr tutorial). This function was not able to identify independent LD blocks based on the multi-population dataset based on a range of parameters (we considered r^2^ up to 0.3), likely due to admixture, so we did not apply PRS-CS on the multi-population GWAS summary statistics and only used it based on individual HARE group summary statistics.

For PRS-CS and PRS-CSx, we outputted the SNPs in each LD block, used plink v.1.9 to create correlation matrices, and next combined them by chromosome.

### PGS development

We developed PGSs using LDPred2, PRS-CS, and PRS-CSx. The latter method is a multi-PGS combination method that relies on summary statistics from GWAS performed in populations of distinct ancestral make-up and develops PGS matching each of the ancestries, to be later combined as a weighted (or unweighted) sum. We used the reference panels developed in MVP as well as the reference panels provided with the PRS-CS and PRS-CSx software, which are based on subpopulations of the UKB program, and develop PGS for OSA based on BMI-adjusted and BMI-unadjusted analyses. We also developed PGSs using PRS-CS and LDPred2 and combined them as sums of PGSs. We considered the following PGS constructions:

1. **Only based on MVP, HARE group specific GWAS:** using LDPred2 based on MVP HARE group reference panels; using PRS-CS and PRS-CSx based on MVP HARE group LD reference panels; using PRS-CS and PRS-CSx based on UKB LD reference panels. The difference between PRS-CS and PRS-CSx is that PRS-CSx constructs the HARE groups (or ancestry)-specific PGS while borrowing information from other groups while PRS-CS does not.
2. **Only using FinnGen:** using LDPred2 and PRS-CS based on MVP and UKBB European reference panels.
3. **Based on similar ancestry meta-analysis of MVP, FinnGen, and Han Chinese OSA GWAS summary statistics**. Here, we meta-analyzed MVP Asian + Han Chinese OSA GWAS (only BMI unadjusted due to data availability), and meta-analyzed MVP White HARE group with FinnGen Finish Europeans. We then used the same approaches as in bullet 1 above.
4. **PGS based on multi-population meta-analysis:** using the meta-analysis of summary statistics of MVP only, and similarly using meta-analysis of all MVP, FinnGen, and Han Chinse GWAS, with LDPred2 and the multi-population MVP reference panel.

Ancestry/population-specific PGSs developed as described in bullet points 1-3 above were combined together (as described below) as weighted or unweighted sums.

We also developed PGSs based on sex-specific GWAS summary statistics. Because the sample size of female participants is small in MVP, especially of non-White participants, for female sex PGSs we developed MVP-only PGS using the multi-ethnic female GWAS results with LDPred2, and a European population-based PGS using the White MVP female GWAS summary statistics meta-analyzed with the FinnGen female GWAS summary statistics. For male-only PGS, we used the same methods as described above for the sex-combined PGSs.

### MGB Biobank

The Mass General Brigham (MGB) biobank is a biorepository of consented patient samples at MGB (Supplementary Note 2). We extracted data from MGB biobank in November 2021. OSA status was extracted from the field “Obstructive Sleep Apnea OSA”. We used the dataset to derive PGS combination weights. In brief, we used (1) PRS-CSx PGSs as well as, separately, (2) single population PGSs (MVP HARE groups only and combined with FinnGen and Han Chinese OSA GWAS) developed using LDPred2 or (separately) PRS-CS, in logistic regression of OSA over the set of PGSs. The effect estimates of the PGSs are the weights used later to create a sum of PGSs, e.g.: w_1_xPGS_1_ + w_2_xPGS_2_ + w_3_xPGS_3_. Analyses in MGB Biobank were adjusted for current age, sex, self-reported race/ethnicity, genotyping batch, and BMI (linear and squared terms), with BMI being the median BMI in the health records for each person. We performed both BMI-unadjusted and BMI-adjusted analyses, where the adjustment in MGB matched the BMI adjustment used in the original GWAS that the relevant PGSs were constructed by.

### Deriving a single set of PGS variant weights from a multiple PGS combination

To facilitate replication of PGS association analyses as well as general use of PGS in external studies, we here demonstrate how we derive a new PGS, that accounts for the weighted (or unweighted) summation of multiple, standardized, PGSs. The formula for the *k*th PGS is:

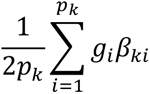

Where note that is standardized according to the number of potential alleles (2 times the number of variants). Suppose that we computed the mean and standard deviation of the PGSs in the evaluation dataset (here, TOPMed), and they are given by *μ_k_*, *σ_k_* for the *k* PGS. Noting that the number of variants can change between PGSs, we denote by *p_k_* the number of variants in the *k*th PGS. When summing *K* PGSs with weights, *w*_1_, …, *w_K_* the expression is:

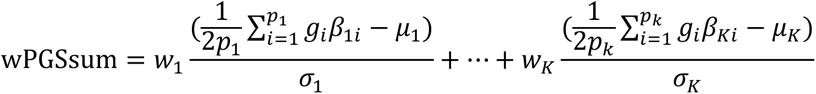

This can alternatively be written as a single PGS formula after some rearrangement of terms, and harmonizing all variants to be considered as being a single set of *p* variants, with a variant potentially having a *β* weight of zero in any specific PGS, then:

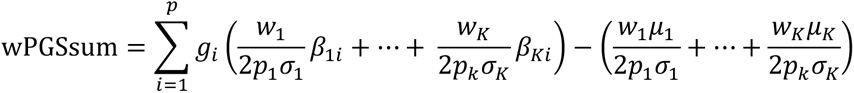

Therefore, we can compute wPGSsum as a single PGS, with the *β* weight for each variant being obtained as a weighted sum of the *β* weights of the individual PGSs. In other words, we can multiply the variant *β* weights in the *k* PGS by *w_k_*/(2*p_k_σ_k_*), with *w_k_* being the weight from the PGS summation, and *σ_k_* being the PGS standard deviation in the TOPMed dataset.

### The TOPMed dataset

The TOPMed dataset provides aggregated whole genome sequencing (WGS) across multiple studies. We used WGS data to construct and assess PGS associations using several studies with available OSA phenotypes. Some studies performed at-home, over-night sleep studies (ARIC, CARDIA, CHS, and FHS via the Sleep Heart Heath Study (51), CFS (52), HCHS/SOL (53), JHS (54), MESA (55)), and only one study used a questionnaire-based OSA status (COPDGene). Detailed descriptions of these studies are provided in Supplementary Note 3.

#### OSA phenotype used in PGS assessment

Supplementary Table 16 describe the OSA assessment across these studies. In brief, we defined OSA in studies that used over-night measurements using the Apnea-Hypopnea Index (AHI) or the Respiratory Event Index (REI) as defined by each study, with AHI/REI≥5 (mild-to-severe OSA) and with AHI/REI≥15 (moderate-to-severe OSA). Because COPDGene used self-reported doctor diagnosed OSA, while other TOPMed studies used a quantitative index, we compared using OSA definition, for studies using an index, as either mild-to-severe OSA versus no OSA, or using moderate-to-severe OSA versus no or mild OSA. To focus the potential analyses, this comparison was performed using the full available TOPMed OSA dataset and the multi-ethnic LDPred2-based MVP PGS only. We next performed other PGS evaluations with the selected OSA definition. The selected OSA definition was the one that had stronger association with the PGS.

#### Using TOPMed to identify an optimal multi-ethnic OSA PGS

Whole genome sequencing and quality control in TOPMed are described in Supplementary Note 4. We constructed the developed PGSs in TOPMed using PRCise 2 (v2.3.1.e; without any clumping nor thresholding). We estimated the association between the PGS and OSA in mixed models implemented in the GENESIS R package (version v2.16.1), with relatedness modeled via a sparse kinship matrix. All models were further adjusted for age, sex, BMI (linear and squared terms), self-reported race/ethnicity, and the first 11 principal components (PCs) of genetic data to prevent population stratification. We note here that we adjusted for self-reported race/ethnicity because health disparities related to the different sociocultural and structured environment experienced by different demographic groups may result in differences in OSA rates between them (56).

We assessed OSA PGS in a multi-step process.

1. **Choosing a reference panel for ancestry-specific PGS development when using PRS-CS.** We compared PGS developed with our MVP LD reference panel to those developed using the existing UKBB LD reference panels. For this comparison we focused on ancestry-specific PGSs developed using the PRS-CS method. If UKBB reference panel performed better or equally well, we proceeded with the UKBB reference panel in PGS using the PRS-CS software, because the UKK reference panel is publicly available and thus more useful to the research community.
2. **Compare ancestry-specific PRS-CS and LDPred2 PGSs**. We compared PRS-CS PGSs developed with the reference panel selected in step 1, with PGSs generated by LDPred2 using the MVP LD reference panel. For LDPred2 we only used the MVP reference panel because the software does not provide a reference panel.
3. **Compare PGS based on multi-ethnic meta-analysis to weighted ancestry-specific PGS combinations.** We compared PGSs constructed based on multi-ethnic meta-analyses and PGSs constructed as PGS combinations: combinations of PRS-CSx-derived PGSs, and combinations of either PRS-CS or LDPred2 PGSs according to step 2 comparison. We used both weighted and unweighted analyses.

As the conclusion of step 3, we selected the optimal multi-ethnic OSA PGSs based on BMI-adjusted and BMI-unadjusted analyses. When comparing PGSs, they were prioritized based on effect size estimates and p-values.

#### Secondary PGS comparisons and analyses in TOPMed

We performed additional comparisons with the multi-ethnic OSA PGS: (a) we compared the selected multi-ethnic PGS to ancestry-specific PGSs within relevant population groups, and (b) we studied the associations of sex-specific PGSs with OSA within sex groups and combined, to assess whether PGS developed based on sex-specific GWAS perform better within their corresponding sex groups, and whether the GWAS sample size is sufficient (especially for female participants) to develop sex-specific PGSs.

#### OSA and lung function PGS associations with OSA phenotypes and with other sleep phenotypes in TOPMed

Some of the TOPMed studies performed over-night sleep studies, and some administered sleep questions, enabling associations of selected OSA PGSs with additional sleep phenotypes.

We estimated the association of the OSA PGS with overnight sleep-evaluated phenotypes that rely on oxyhemoglobin saturation and desaturation, and are typically associated with OSA. These phenotypes included AHI/REI, hypoxic burden (57), minimum and average oxyhemoglobin saturation during sleep, average oxyhemoglobin desaturation during respiratory events (i.e. desaturation compared to the baseline saturation at the beginning of the sleep period), and percent sleep time under 90% oxyhemoglobin saturation. Some of these phenotypes were further evaluated separately during rapid and non-rapid eye movement (REM and NREM) sleep (23). For comparison, we also constructed a PGS for lung function and evaluated its association with some of these phenotypes. The lung function PGS was constructed based on PGS variants and weights downloaded from the PGS catalog (58), PGS ID PGS001180, based on the ratio between force expiratory volume in 1 second to force vital capacity and developed by 59).

We also estimated the association of the OSA PGS with self-reported sleep phenotypes: sleep duration (harmonized in (60)), short and long sleep, defined as sleep duration>9 hours, the Epworth sleepiness scale (ESS), including an excessive daytime sleepiness phenotype defined as ESS>10, and the Women Health Initiative Insomnia rating scale (WHIIRS), including an insomnia phenotype defined as WHIIRS ≥9. TOPMed studies contributing to each analysis are provided in Supplementary Table 16.

### OSA PGS association with OSA in Geisinger’s MyCode

The MyCode Community Health Initiative (MyCode) Study is a hospital-based cohort study recruited from Geisinger, a large healthcare provider in central Pennsylvania. Subjects that have provided biospecimens have been genotyped through a collaboration with Regeneron Genetics Center as part of the DiscovEHR Study (http://www.discovehrshare.com/), including up to 170,765 participants available at the time of this analysis. All participants were genotyped using the Illumina’s HumanOmniExpressExome (~60,000) or Global Screening Array (~110,000). All array-based data were quality controlled (QC’ed) with standard QC procedures before imputation, including gender mismatches, duplicate samples (identical twins were kept), low individual and SNP call rates (<95%), Hardy-Weinberg equilibrium (P ≤1×10^-15^), and heterozygosity (F >0.4). QC’ed array data was imputed to the TOPMed Release 2 reference panel by array. Following imputation, all variants were filtered if they exhibited a minor allele count (MAC) of less than 5, a mean imputation quality score (R^2^) <0.7, or missingness >10%.

The current analysis was restricted to a subset of unrelated adult MyCode participants up to 2^nd^ degree, selected using PRIMUS (61). OSA cases were defined based on a minimum of three OSA-related ICD-9 (ICD-9: 327.20, 327.23, 327.29, 780.51, 780.53, 780.57) or ICD-10 (G4730, G4733 or G4739) on separate dates, controls included those with zero instances of a relevant OSA ICD code, and those with one or two OSA codes were excluded from the analysis. This phenotype was based on a previous chart validation in the Geisinger study participants (62).

BMIadjOSA and BMIunadjOSA PGSs were generated from files with SNP and weight using PRSice2 without clumping and thresholding. PGS associations with OSA were carried out both stratified and combined across sexes using GENESIS and AUC was estimated using pROC. All analyses were carried out using R. All association analyses were adjusted for age, BMI, BMI^2^, sex (in combined analysis), self-reported or EHR-derived race/ethnicity, and the top 20 genetic PCs.

### OSA PGS associations with outcomes in All of Us

We used WGS data from All of Us, version 6. Sequencing and quality control procedures for All of Us, performed by the All of Us team, are described in https://support.researchallofus.org/hc/en-us/categories/4537007565204-Genomics. The data was available in a HAIL matrix tables. We used python version 3 on the All of Us Researcher Workbench. We first filtered genetic files to keep only HapMap SNPs, then converted the file to BED plink format. Next, we filtered out variants that failed All of Us quality control according to the “filter” flag, and variants with missing call rate >1%. We did not further filter by allele frequency. We constructed the PGSs selected by the TOPMed analysis using plink. And we only used variants with MAF ≥ 0.01. In this dataset, we also compared the TOPMed-selected PGSs to previously-reported PGSs, in order to alleviate potential effects of overfitting, where TOPMed was used to select the main PGSs reported here. We estimated the associations of OSA PGSs with OSA and other clinical outcomes: hypertension, type 2 diabetes, stroke, atrial fibrillation, heart failure, coronary artery disease, asthma, chronic obstructive pulmonary disease, and chronic kidney disease. We performed sex-combined and sex-stratified analyses for all phenotypes, and for OSA, also analyses stratified by self-reported race/ethnicity, obesity category (BMI<30 and ≥ 30) and age categories (age≤40, 40<age≤ 60, age>60). Supplementary Note 5 provides the definitions of the phenotypes used (medical codes, etc.).

### OSA PGS associations across OSA and sleep measures in the Human Phenotype Project

We studied the association of OSA PGS with OSA, both from clinical diagnosis and from sleep monitoring, and with self-reported sleep measures in the HPP (63). Home sleep study was performed using the WatchPAT device (Itamar medical) over three nights. To determine OSA status based on sleep monitoring, we averaged the AHI measures from all available nights and then used AHI≥15 as cutoffs for defining OSA. We also estimated PGS associations with NREM and REM AHI, ODI, and RDI, and device-measured sleep duration, as continuous measures in association with the OSA PGS. Analyses were adjusted for age, sex, and BMI (linear and squared terms). Self-reported sleep phenotypes were available via questionnaires mimicking the UKBB project. We estimated the associations of OSA PGS with the responses to the following questions: “Daily sleep hours”, “Tired or little energy fortnight”, “Consider yourself morning evening”, “Easy getting up”, “Nap during day”, and “Trouble falling asleep”. We adjusted for the same variables as before.

### OSA PGS associations with body fat DXA measures in the Human Phenotype Project

We estimated the associations of the developed OSA PGSs with DXA measures available from the HPP. The relevant available measures Included VAT and SAT masses across the entire body (total scan), total mass, and gynoid and android masses, where gynoid correspond to the hips area and android to the waist area. We used 6 measures: ratios of VAT, SAT, gynoid, and android masses out of the total mass, and VAT:SAT and gynoid:android mass ratios. For each of the raw measures, we first checked for and removed any outliers. For distributions that appeared log-normal, we applied log transformation, and if the association results were the same (similar direction and p-value) as in the untransformed phenotype, we used the untransformed phenotype. We used only genetically unrelated individuals, performed BMi-adjusted and -unadjusted analysis using the same covariates as described in the previous section, and performed both sex-combined and sex-stratified analyses.

## Data availability statement

Summary statistics from BMI-adjusted and BMI-unadjusted OSA GWAS, stratified by sex and by HARE group, have been deposited on dbGaP, study accession phs001672.

Individual-level genotypes and register data from FinnGen participants can be accessed by approved researchers via the Fingenious portal (https://site.fingenious.fi/en/) hosted by the Finnish Biobank Cooperative FinBB (https://finbb.fi/en/).

Full summary statistics for the genome-wide association study can be accessed from https://figshare.com/ (DOI: 10.6084/m9.figshare.20033246) and Bio-X institutes website (http://analysis.bio-x.cn/gwas/).

MGB Biobank genotyping and phenotypic data are available to Mass General Brigham investigators with required approval from the Mass General Brigham Institutional Review board (IRB).

BMIadjOSA-PGS and BMIunadjOSA-PGS variant and weights will be deposited to the PGS Catalog. There are also available on the GitHub repository https://github.com/nkurniansyah/OSA_PRS. We also used additional PGSs for performance comparisons. All are available on the PGS catalog (LDPred2 OSA PGS from Zhang et al. 2022: PGS003479, genome-wide significant variants OSA PGS based on BMI-adjusted MVP GWAS: PGS003858, genome-wide significant variants OSA PGS based on BMI-unadjusted MVP GWAS: PGS003857, pulmonary function PGS: PGP000244).

TOPMed freeze 8 WGS data are available by application to dbGaP according to the study specific accessions: ARIC: “phs001211“, CFS: “phs000954”, CHS: “phs001368”, COPD-Gene: “phs000951”, FHS: “phs000974”, HCHS/SOL: “phs001395”, JHS: “phs000964”, MESA: “phs001211”, WHI: “phs001237”. Phenotype data are available from dbGaP according to the study-specific accessions: ARIC: “phs000280“, CFS: “phs000284”, CHS: “phs000287”, COPD-Gene: “phs000179”, FHS: “phs000007”, HCHS/SOL: “phs000810”, JHS: “phs000286”, MESA: “phs000209”, WHI: “phs000200”.

Data from the NIH All of Us study are available via institutional data access for researchers who meet the criteria for access to confidential data. To register as a researcher with All of Us, researchers may use the following URL and complete the laid out steps: https://www.researchallofus.org/register/. Researchers can contact All of Us Researcher Workbench Support at.

Data in this paper is part of the Human Phenotype Project (HPP) and is accessible to researchers from universities and other research institutions at: https://humanphenotypeproject.org/data-access. Interested bona fide researchers should contact to obtain instructions for accessing the data.

MyCode data can be accessed by Geisinger investigators. There are restrictions to the sharing of MyCode DiscovEHR genetic datasets related to agreements between Geisinger and the Regeneron Genetics Center.

## Code availability statement

We provide developed scripts used to perform analyses described in the paper and code to construct the OSA-PGSs in the GitHub repository https://github.com/nkurniansyah/OSA_PRS and the Zenodo repository (will provided upon paper acceptance)

## Ethics statement

MVP received ethical/study protocol approval from the VA Central Institutional Review Board, and written informed consent was obtained for all participants.

Study subjects in FinnGen provided informed consent for biobank research, based on the Finnish Biobank Act. Full ethnics statement is provided in Supplementary Note 6.

The All of Us research program was approved by a single IRB, the “All of Us IRB”, which is charged with reviewing the protocol, informed consent, and other participant-facing materials for the *All of Us* Research Program. The IRB follows the regulations and guidance of the Office for Human Research Protections for all studies, ensuring that the rights and welfare of research participants are overseen and protected uniformly. More information is provided online https://allofus.nih.gov/about/who-we-are/institutional-review-board-irb-of-all-of-us-research-program.

Ethics statement for MGB Biobank is provided in Supplementary Note 2, and for TOPMed studies in Supplementary Note 3.

The Human Phenotype Project’s data used in this work was approved by the Weizmann Institute’s IRB for the 10K study, protocol 578-1.

The MyCode Study was approved by the Geisinger Institutional Review Board and all participants provided informed consent. The current analysis consisted of secondary analysis of existing de-identified data and was deemed to be not “human subjects” research as defined in 45 CFR 46.102(f).

## Acknowledgments

This work is supported by National Heart, Lung, and Blood Institute (NHLBI) grant R01HL161012 and National Aging Institute grant R01AG080598 grant to TS. This research is based on data from the Million Veteran Program, Office of Research and Development, Veterans Health Administration, and was supported by award #MVP001 and #BX004821. TS was also supported by Human Genome Research Institute (NHGRI) grant R56HG013163. HMO was supported by National Cancer Institute grant R01CA283469, National Institute of Allergy and Infectious Diseases grant R01AI170850, and by NHGRI grant R01HG012810. This analysis was approved by the Beth Israel Deaconess Medical Center Committee on Clinical Investigations, protocol #2023P000279, and by the Mass General Brigham IRB, protocol #2021P001928. We are grateful to the Million Veteran Program participants and staff. This publication does not represent the views of the Department of Veteran Affairs or the United States Government. We want to acknowledge the participants and investigators of FinnGen study. See Supplementary Note 6 for additional FinnGen acknowledgements. FinnGen consortium members and their affiliations are provided in Supplementary Note 9. The All of Us Research Program would not be possible without the partnership of its participants. See Supplementary Note 5 for additional All of Us acknowledgements. We thank Mass General Brigham Biobank for providing samples, genomic data, and health information data. This study was performed as a collaboration of the NHLBI Trans-Omics in Precision Medicine (TOPMed) Consortium. We gratefully acknowledge the studies and participants who provided biological samples and data for TOPMed and CCDG. TOPMed and CCDG acknowledgements are provided in Supplementary Note 7, and descriptions, acknowledgements, and ethics statements of contributing studies are provided in Supplementary Note 3. TOPMed consortium researchers and their affiliations are listed in Supplementary Note 7. The views expressed in this manuscript are those of the authors and do not necessarily represent the views of the National Heart, Lung, and Blood Institute; the National Institutes of Health; or the U.S. Department of Health and Human Services. Data used in this paper was kindly provided by the Human Phenotype Project (HPP) global initiative. The Human Phenotype Project Datasets can be accessed using this link: https://humanphenotypeproject.org/home

**Geisinger’s MyCode Community Health Initiative Study (MyCode):** We thank all the participants of the MyCode Study. We thank the members of the Geisinger-Regeneron DiscovEHR Collaboration who have been critical in the generation of the genetic and transcriptomic data used in this study. Anne E. Justice (AEJ), Geetha Chittoor (GC), and Shreyash Gupta (SG) were all funded by the National Institutes of Health (NIH), National Heart Lung and Blood Institute (NHLBI) grant P01 HL160471 – 8274.

## Conflicts of interests

Dr. Redline discloses consulting relationships with Eli Lilly Inc. Additionally, Dr. Redline serves as an unpaid member of the Apnimed Scientific Advisory Board, as an unpaid board member for the Alliance for Sleep Apnoea Partners, and has received loaned equipment for a multi-site study: oxygen concentrators from Philips Respironics and polysomnography equipment from Nox Medical. BP serves on the Steering Committee of the Yale Open Data Access Project funded by Johnson & Johnson. SSR and LMR are consultants to the TOPMed Administrative Coordinating Center (through Westat®).

## Author contributions statement

NK and TS conceptualized the study. NK performed data analysis in MVP, TOPMed, MGB biobank, and All of Us. YH performed data analysis in the HPP. SJS performed OSA GWAS in FinnGen. SG contributed to phenotype extraction and generation in MyCode. AEJ and GC performed PGS association analyses in MyCode. NK finalized all tables and figures. NK, BEC, BS, HW harmonized phenotypes in TOPMed. MM, BH, SYJ, RK, JIR, SSR, SG, TB, MF, AM, HOB, MF, LH, and SR contributed to study design, data collection, and data management, in TOPMed cohorts they represent. JH, PW, KC, and DG contributed to study design and data management in MVP. AEJ, BTK and AP contributed to Geisinger study design and data management. HMO contributed to FinnGen study design and data management. NK, SJS, AEJ, and TS drafted the manuscript. TS supervised the work. NK, SJT, GC, SG, AEJ, YH, BTK, BEC, BWS, HW, JH, MRM, BH, SYJ, LMR, RK, JIR, SSR, SAG, TMB, PYL, HC, MF, LH, DL, AM, HMO-B, BP, PWFW, AIP, HMO, SR, DJG, and TS all critically reviewed the manuscript and approved its final version.

## Notes

### Author Declarations

MVP received ethical/study protocol approval from the VA Central Institutional Review Board, and written informed consent was obtained for all participants. Study subjects in FinnGen provided informed consent for biobank research in accordance with the Finnish Biobank Act. The full ethics statement is as follows: Study subjects in FinnGen provided informed consent for biobank research, based on the Finnish Biobank Act. Alternatively, separate research cohorts, collected prior the Finnish Biobank Act came into effect (in September 2013) and start of FinnGen (August 2017), were collected based on study-specific consents and later transferred to the Finnish biobanks after approval by Fimea (Finnish Medicines Agency), the National Supervisory Authority for Welfare and Health. Recruitment protocols followed the biobank protocols approved by Fimea. The Coordinating Ethics Committee of the Hospital District of Helsinki and Uusimaa (HUS) statement number for the FinnGen study is Nr HUS/990/2017. The FinnGen study Is approved by Finnish Institute for Health and Welfare (permit numbers: THL/2031/6.02.00/2017, THL/1101/5.05.00/2017, THL/341/6.02.00/2018, THL/2222/6.02.00/2018, THL/283/6.02.00/2019, THL/1721/5.05.00/2019 and THL/1524/5.05.00/2020), Digital and population data service agency (permit numbers: VRK43431/2017-3, VRK/6909/2018-3, VRK/4415/2019-3), the Social Insurance Institution (permit numbers: KELA 58/522/2017, KELA 131/522/2018, KELA 70/522/2019, KELA 98/522/2019, KELA 134/522/2019, KELA 138/522/2019, KELA 2/522/2020, KELA 16/522/2020), Findata permit numbers THL/2364/14.02/2020, THL/4055/14.06.00/2020, THL/3433/14.06.00/2020, THL/4432/14.06/2020, THL/5189/14.06/2020, THL/5894/14.06.00/2020, THL/6619/14.06.00/2020, THL/209/14.06.00/2021, THL/688/14.06.00/2021, THL/1284/14.06.00/2021, THL/1965/14.06.00/2021, THL/5546/14.02.00/2020, THL/2658/14.06.00/2021, THL/4235/14.06.00/2021, Statistics Finland (permit numbers: TK-53-1041-17 and TK/143/07.03.00/2020 (earlier TK-53-90-20) TK/1735/07.03.00/2021, TK/3112/07.03.00/2021) and Finnish Registry for Kidney Diseases permission/extract from the meeting minutes on 4th July 2019. The Biobank Access Decisions for FinnGen samples and data utilized in FinnGen Data Freeze 10 include: THL Biobank BB2017_55, BB2017_111, BB2018_19, BB_2018_34, BB_2018_67, BB2018_71, BB2019_7, BB2019_8, BB2019_26, BB2020_1, BB2021_65, Finnish Red Cross Blood Service Biobank 7.12.2017, Helsinki Biobank HUS/359/2017, HUS/248/2020, HUS/430/2021 28, 29, HUS/150/2022 12, 13, 14, 15, 16, 17, 18, 23, 58 and 59, Auria Biobank AB17-5154 and amendment #1 (August 17 2020) and amendments BB_2021-0140, BB_2021- 0156 (August 26 2021, Feb 2 2022), BB_2021-0169, BB_2021-0179, BB_2021-0161, AB20-5926 and amendment #1 (April 23 2020) and it ́s modification (Sep 22 2021), BB_2022-0262, BB_2022-0256, Biobank Borealis of Northern Finland_2017_1013, 2021_5010, 2021_5018, 2021_5015, 2021_5015 Amendment, 2021_5023, 2021_5023 Amendment, 2021_5017, 2022_6001, 2022_6006 Amendment, BB22-0067, 2022_0262, Biobank of Eastern Finland 1186/2018 and amendment 22/2020, 53/2021, 13/2022, 14/2022, 15/2022, 27/2022, 28/2022, 29/2022, 33/2022, 35/2022, 36/2022, 37/2022, 39/2022, 7/2023, Finnish Clinical Biobank Tampere MH0004 and amendments (21.02.2020 & 06.10.2020), 8/2021, 9/2021, 9/2022, 10/2022, 12/2022, 13/2022, 20/2022, 21/2022, 22/2022, 23/2022, 28/2022, 29/2022, 30/2022, 31/2022, 32/2022, 38/2022, 40/2022, 42/2022, 1/2023, Central Finland Biobank 1-2017, BB_2021-0161, BB_2021-0169, BB_2021-0179, BB_2021-0170, BB_2022-0256, and Terveystalo Biobank STB 2018001 and amendment 25th Aug 2020, Finnish Hematological Registry and Clinical Biobank decision 18th June 2021, Arctic biobank P0844: ARC_2021_1001. The All of Us research program was approved by a single IRB, the 'All of Us IRB', which is charged with reviewing the protocol, informed consent, and other participant-facing materials for the All of Us Research Program. The IRB follows the regulations and guidance of the Office for Human Research Protections for all studies, ensuring that the rights and welfare of research participants are overseen and protected uniformly. More information is provided online: https://allofus.nih.gov/about/who-we-are/institutional-review-board-irb-of-all-of-us-research-program. All MGB Biobank subjects have provided their consent to join the MGB Biobank, which includes agreeing to provide a blood sample linked to the electronic medical record. Subjects also agree to be recontacted by the MGB Biobank staff as needed. This manuscript utilized data from 10 different studies within TOPMed. Ethical approval was obtained for each individual study, as follows: ARIC: The ARIC study has been approved by a single Institutional Review Board (sIRB) at Johns Hopkins School of Medicine and Institutional Review Boards (IRB) at all participating institutions: University of North Carolina at Chapel Hill IRB, Johns Hopkins University School of Public Health IRB, University of Minnesota IRB, Wake Forest University Health Sciences IRB, and University of Mississippi Medical Center IRB. Study participants provided written informed consent at all study visits. CARDIA: All CARDIA participants provided informed consent, and the study was approved by the Institutional Review Boards of the University of Alabama at Birmingham and the University of Texas Health Science Center at Houston. CFS: Cleveland Family Study was approved by the Institutional Review Board (IRB) of Case Western Reserve University and Mass General Brigham (formerly Partners HealthCare). Written informed consent was obtained from all participants. CHS: CHS was approved by institutional review committees at each field center and individuals in the present analysis had available DNA and gave informed consent including consent to use of genetic information for the study of cardiovascular disease. COPDGene: All COPDGene participants provided written informed consent, and the study was approved by the Institutional Review Boards of the participating clinical centers. FHS: The Framingham Heart Study was approved by the Institutional Review Board of the Boston University Medical Center. All study participants provided written informed consent. HCHS/SOL: This study was approved by the institutional review boards (IRBs) at each field center, where all participants gave written informed consent, and by the Non-Biomedical IRB at the University of North Carolina at Chapel Hill, to the HCHS/SOL Data Coordinating Center. All IRBs approving the study are: Non-Biomedical IRB at the University of North Carolina at Chapel Hill. Chapel Hill, NC; Einstein IRB at the Albert Einstein College of Medicine of Yeshiva University. Bronx, NY; IRB at Office for the Protection of Research Subjects (OPRS), University of Illinois at Chicago. Chicago, IL; Human Subject Research Office, University of Miami. Miami, FL; Institutional Review Board of San Diego State University. San Diego, CA. JHS: The Institutional Review Boards at Jackson State University, Tougaloo College, and the University of Mississippi Medical Center approved the study, and all participants provided written informed consent. MESA: All MESA participants provided written informed consent, and the study was approved by the Institutional Review Boards at The Lundquist Institute (formerly Los Angeles BioMedical Research Institute) at Harbor-UCLA Medical Center, University of Washington, Wake Forest School of Medicine, Northwestern University, University of Minnesota, Columbia University, and Johns Hopkins University. WHI: All WHI participants provided informed consent and the study was approved by the Institutional Review Board (IRB) of the Fred Hutchinson Cancer Research Center. The Human Phenotype Project's data used in this work was approved by the Weizmann Institute's IRB for the 10K study, protocol 578-1. The MyCode Study was approved by the Geisinger Institutional Review Board and all participants provided informed consent. The current analysis consisted of secondary analysis of existing de-identified data and was deemed to be not human subjects research as defined in 45 CFR 46.102(f).

## References

1. Benjafield AV, Ayas NT, Eastwood PR, Heinzer R, Ip MSM, Morrell MJ, et al. Estimation of the global prevalence and burden of obstructive sleep apnoea: a literature-based analysis. Lancet Respir Med. 2019 Aug;7(8):687–98.

2. Mokhlesi B, Ham SA, Gozal D. The effect of sex and age on the comorbidity burden of OSA: an observational analysis from a large nationwide US health claims database. Eur Respir J. 2016 Apr;47(4):1162–9.

3. Bradley TD, Floras JS. Obstructive sleep apnoea and its cardiovascular consequences. Lancet. 2009 Jan 3;373(9657):82–93.

4. Marin JM, Carrizo SJ, Vicente E, Agusti AGN. Long-term cardiovascular outcomes in men with obstructive sleep apnoea-hypopnoea with or without treatment with continuous positive airway pressure: an observational study. Lancet. 2005 Mar 25;365(9464):1046–53.

5. Kapur V, Strohl KP, Redline S, Iber C, O’Connor G, Nieto J. Underdiagnosis of sleep apnea syndrome in U.S. communities. Sleep Breath. 2002 Jun;6(2):49–54.

6. Stansbury R, Strollo P, Pauly N, Sharma I, Schaaf M, Aaron A, et al. Underrecognition of sleep-disordered breathing and other common health conditions in the West Virginia Medicaid population: a driver of poor health outcomes. J Clin Sleep Med. 2022 Mar 1;18(3):817–24.

7. Kapur VK, Auckley DH, Chowdhuri S, Kuhlmann DC, Mehra R, Ramar K, et al. Clinical practice guideline for diagnostic testing for adult obstructive sleep apnea: an american academy of sleep medicine clinical practice guideline. J Clin Sleep Med. 2017 Mar 15;13(3):479–504.

8. Young T, Skatrud J, Peppard PE. Risk factors for obstructive sleep apnea in adults. JAMA. 2004 Apr 28;291(16):2013–6.

9. Mistry S, Harrison JR, Smith DJ, Escott-Price V, Zammit S. The use of polygenic risk scores to identify phenotypes associated with genetic risk of schizophrenia: Systematic review. Schizophr Res. 2018 Jul;197:2–8.

10. Zhang Y, Elgart M, Kurniansyah N, Spitzer BW, Wang H, Kim D, et al. Genetic determinants of cardiometabolic and pulmonary phenotypes and obstructive sleep apnoea in HCHS/SOL. EBioMedicine. 2022 Oct;84:104288.

11. Richardson TG, Harrison S, Hemani G, Davey Smith G. An atlas of polygenic risk score associations to highlight putative causal relationships across the human phenome. eLife. 2019 Mar 5;8.

12. Ye Y, Chen X, Han J, Jiang W, Natarajan P, Zhao H. Interactions between enhanced polygenic risk scores and lifestyle for cardiovascular disease, diabetes, and lipid levels. Circ Genom Precis Med. 2021 Feb;14(1):e003128.

13. Domingue BW, Trejo S, Armstrong-Carter E, Tucker-Drob EM. Interactions between Polygenic Scores and Environments: Methodological and Conceptual Challenges. Sociol Sci. 2020 Sep 21;7:465–86.

14. Hüls A, Wright MN, Bogl LH, Kaprio J, Lissner L, Molnár D, et al. Polygenic risk for obesity and its interaction with lifestyle and sociodemographic factors in European children and adolescents. Int J Obes (Lond). 2021 Jun;45(6):1321–30.

15. Kurniansyah N, Goodman MO, Khan AT, Wang J, Feofanova E, Bis JC, et al. Evaluating the use of blood pressure polygenic risk scores across race/ethnic background groups. Nat Commun. 2023 Jun 2;14(1):3202.

16. Wray NR, Lin T, Austin J, McGrath JJ, Hickie IB, Murray GK, et al. From basic science to clinical application of polygenic risk scores: A primer. JAMA Psychiatry. 2021 Jan 1;78(1):101–9.

17. Lewis CM, Vassos E. Polygenic risk scores: from research tools to clinical instruments. Genome Med. 2020 May 18;12(1):44.

18. Choi SW, García-González J, Ruan Y, Wu HM, Porras C, Johnson J, et al. PRSet: Pathway-based polygenic risk score analyses and software. PLoS Genet. 2023 Feb 7;19(2):e1010624.

19. Goodman MO, Cade BE, Shah NA, Huang T, Dashti HS, Saxena R, et al. Pathway-Specific Polygenic Risk Scores Identify Obstructive Sleep Apnea-Related Pathways Differentially Moderating Genetic Susceptibility to Coronary Artery Disease. Circ Genom Precis Med. 2022 Oct;15(5):e003535.

20. Redline S, Azarbarzin A, Peker Y. Obstructive sleep apnoea heterogeneity and cardiovascular disease. Nat Rev Cardiol. 2023 Aug;20(8):560–73.

21. Sofer T, Kurniansyah N, Murray M, Ho Y-L, Abner E, Esko T, et al. Genome-wide association study of obstructive sleep apnoea in the Million Veteran Program uncovers genetic heterogeneity by sex. EBioMedicine. 2023 Apr;90:104536.

22. Strausz S, Ruotsalainen S, Ollila HM, Karjalainen J, Kiiskinen T, Reeve M, et al. Genetic analysis of obstructive sleep apnoea discovers a strong association with cardiometabolic health. Eur Respir J. 2021 May 6;57(5).

23. Won CHJ, Reid M, Sofer T, Azarbarzin A, Purcell S, White D, et al. Sex differences in obstructive sleep apnea phenotypes, the multi-ethnic study of atherosclerosis. Sleep. 2020 May 12;43(5).

24. Gami AS, Caples SM, Somers VK. Obesity and obstructive sleep apnea. Endocrinol Metab Clin North Am. 2003 Dec;32(4):869–94.

25. Fang H, Hui Q, Lynch J, Honerlaw J, Assimes TL, Huang J, et al. Harmonizing Genetic Ancestry and Self-identified Race/Ethnicity in Genome-wide Association Studies. Am J Hum Genet. 2019 Oct 3;105(4):763–72.

26. Wendt FR, Pathak GA, Vahey J, Qin X, Koller D, Cabrera-Mendoza B, et al. Modeling the longitudinal changes of ancestry diversity in the Million Veteran Program. Hum Genomics. 2023 Jun 2;17(1):46.

27. Redline S, Yenokyan G, Gottlieb DJ, Shahar E, O’Connor GT, Resnick HE, et al. Obstructive sleep apnea-hypopnea and incident stroke: the sleep heart health study. Am J Respir Crit Care Med. 2010 Jul 15;182(2):269–77.

28. Gottlieb DJ, Yenokyan G, Newman AB, O’Connor GT, Punjabi NM, Quan SF, et al. Prospective study of obstructive sleep apnea and incident coronary heart disease and heart failure: the sleep heart health study. Circulation. 2010 Jul 27;122(4):352–60.

29. Gueye-Ndiaye S, Gunnlaugsson S, Li L, Gaffin JM, Zhang Y, Sofer T, et al. Asthma and Sleep-disordered Breathing Overlap in School-aged Children. Ann Am Thorac Soc. 2024 Jun;21(6):986–9.

30. Prasad B, Nyenhuis SM, Imayama I, Siddiqi A, Teodorescu M. Asthma and obstructive sleep apnea overlap: what has the evidence taught us? Am J Respir Crit Care Med. 2020 Jun 1;201(11):1345–57.

31. Ryan S, Nolan GM, Hannigan E, Cunningham S, Taylor C, McNicholas WT. Cardiovascular risk markers in obstructive sleep apnoea syndrome and correlation with obesity. Thorax. 2007 Jun;62(6):509–14.

32. Gami AS, Hodge DO, Herges RM, Olson EJ, Nykodym J, Kara T, et al. Obstructive sleep apnea, obesity, and the risk of incident atrial fibrillation. J Am Coll Cardiol. 2007 Feb 6;49(5):565–71.

33. Bauters F, Rietzschel ER, Hertegonne KBC, Chirinos JA. The link between obstructive sleep apnea and cardiovascular disease. Curr Atheroscler Rep. 2016 Jan;18(1):1.

34. Zonato AI, Martinho FL, Bittencourt LR, de Oliveira Camponês Brasil O, Gregório LC, Tufik S. Head and neck physical examination: comparison between nonapneic and obstructive sleep apnea patients. Laryngoscope. 2005 Jun;115(6):1030–4.

35. Molnár V, Lakner Z, Molnár A, Tárnoki DL, Tárnoki ÁD, Kunos L, et al. The Predictive Role of the Upper-Airway Adipose Tissue in the Pathogenesis of Obstructive Sleep Apnoea. Life (Basel). 2022 Oct 4;12(10).

36. Oğretmenoğlu O, Süslü AE, Yücel OT, Onerci TM, Sahin A. Body fat composition: a predictive factor for obstructive sleep apnea. Laryngoscope. 2005 Aug;115(8):1493–8.

37. Kammerlander AA, Lyass A, Mahoney TF, Massaro JM, Long MT, Vasan RS, et al. Sex differences in the associations of visceral adipose tissue and cardiometabolic and cardiovascular disease risk: the framingham heart study. J Am Heart Assoc. 2021 Jun;10(11):e019968.

38. Lebovitz HE, Banerji MA. Point: visceral adiposity is causally related to insulin resistance. Diabetes Care. 2005 Sep;28(9):2322–5.

39. Kolb H. Obese visceral fat tissue inflammation: from protective to detrimental? BMC Med. 2022 Dec 27;20(1):494.

40. Huang T, Sands SA, Stampfer MJ, Tworoger SS, Hu FB, Redline S. Insulin resistance, hyperglycemia, and risk of developing obstructive sleep apnea in men and women in the united states. Ann Am Thorac Soc. 2022 Oct;19(10):1740–9.

41. Godoy IRB, Martinez-Salazar EL, Eajazi A, Genta PR, Bredella MA, Torriani M. Fat accumulation in the tongue is associated with male gender, abnormal upper airway patency and whole-body adiposity. Metab Clin Exp. 2016 Nov;65(11):1657–63.

42. Borsoi L, Armeni P, Donin G, Costa F, Ferini-Strambi L. The invisible costs of obstructive sleep apnea (OSA): Systematic review and cost-of-illness analysis. PLoS ONE. 2022 May 20;17(5):e0268677.

43. Sofer T. Overcoming the underdiagnosis of obstructive sleep apnea to empower genetic association analyses. Sleep. 2023 Mar 9;46(3).

44. Kaufmann CN, Spira AP, Wickwire EM, Albrecht JS, Amjad H, Jackson CL, et al. Disparities in the Diagnosis and Treatment of Obstructive Sleep Apnea Among Middle-aged and Older Adults in the United States. Ann Am Thorac Soc. 2023 Jun;20(6):921–6.

45. Privé F, Arbel J, Vilhjálmsson BJ. LDpred2: better, faster, stronger. Bioinformatics. 2021 Apr 1;36(22–23):5424–31.

46. Ge T, Chen C-Y, Ni Y, Feng Y-CA, Smoller JW. Polygenic prediction via Bayesian regression and continuous shrinkage priors. Nat Commun. 2019 Apr 16;10(1):1776.

47. Ruan Y, Lin Y-F, Feng Y-CA, Chen C-Y, Lam M, Guo Z, et al. Improving polygenic prediction in ancestrally diverse populations. Nat Genet. 2022 May 5;54(5):573–80.

48. Xu H, Liu F, Li Z, Li X, Liu Y, Li N, et al. Genome-Wide Association Study of Obstructive Sleep Apnea and Objective Sleep-Related Traits Identifies Novel Risk Loci in Han Chinese Individuals. Am J Respir Crit Care Med. 2022 Jul 12;

49. Kurki MI, Karjalainen J, Palta P, Sipilä TP, Kristiansson K, Donner KM, et al. FinnGen provides genetic insights from a well-phenotyped isolated population. Nature. 2023 Jan 18;613(7944):508–18.

50. Mbatchou J, Barnard L, Backman J, Marcketta A, Kosmicki JA, Ziyatdinov A, et al. Computationally efficient whole-genome regression for quantitative and binary traits. Nat Genet. 2021 Jul;53(7):1097–103.

51. Quan SF, Howard BV, Iber C, Kiley JP, Nieto FJ, O’Connor GT, et al. The Sleep Heart Health Study: design, rationale, and methods. Sleep. 1997 Dec;20(12):1077–85.

52. Redline S, Tishler PV, Tosteson TD, Williamson J, Kump K, Browner I, et al. The familial aggregation of obstructive sleep apnea. Am J Respir Crit Care Med. 1995 Mar;151(3 Pt 1):682–7.

53. Redline S, Sotres-Alvarez D, Loredo J, Hall M, Patel SR, Ramos A, et al. Sleep-disordered breathing in Hispanic/Latino individuals of diverse backgrounds. The Hispanic Community Health Study/Study of Latinos. Am J Respir Crit Care Med. 2014 Feb 1;189(3):335–44.

54. Johnson DA, Guo N, Rueschman M, Wang R, Wilson JG, Redline S. Prevalence and correlates of obstructive sleep apnea among African Americans: the Jackson Heart Sleep Study. Sleep. 2018 Oct 1;41(10).

55. Chen X, Wang R, Zee P, Lutsey PL, Javaheri S, Alcántara C, et al. Racial/Ethnic Differences in Sleep Disturbances: The Multi-Ethnic Study of Atherosclerosis (MESA). Sleep. 2015 Jun 1;38(6):877–88.

56. Khan AT, Gogarten SM, McHugh CP, Stilp AM, Sofer T, Bowers ML, et al. Recommendations on the use and reporting of race, ethnicity, and ancestry in genetic research: Experiences from the NHLBI TOPMed program. Cell Genomics. 2022 Aug 10;2(8).

57. Azarbarzin A, Sands SA, Stone KL, Taranto-Montemurro L, Messineo L, Terrill PI, et al. The hypoxic burden of sleep apnoea predicts cardiovascular disease-related mortality: the Osteoporotic Fractures in Men Study and the Sleep Heart Health Study. Eur Heart J. 2019 Apr 7;40(14):1149–57.

58. Lambert SA, Gil L, Jupp S, Ritchie SC, Xu Y, Buniello A, et al. The Polygenic Score Catalog as an open database for reproducibility and systematic evaluation. Nat Genet. 2021 Apr;53(4):420–5.

59. Tanigawa Y, Qian J, Venkataraman G, Justesen JM, Li R, Tibshirani R, et al. Significant sparse polygenic risk scores across 813 traits in UK Biobank. PLoS Genet. 2022 Mar 24;18(3):e1010105.

60. Sofer T, Lee J, Kurniansyah N, Jain D, Laurie CA, Gogarten SM, et al. BinomiRare: A robust test for association of a rare genetic variant with a binary outcome for mixed models and any case-control proportion. HGG Adv. 2021 Jul 8;2(3).

61. Staples J, Qiao D, Cho MH, Silverman EK, University of Washington Center for Mendelian Genomics, Nickerson DA, et al. PRIMUS: rapid reconstruction of pedigrees from genome-wide estimates of identity by descent. Am J Hum Genet. 2014 Nov 6;95(5):553–64.

62. Keenan BT, Kirchner HL, Veatch OJ, Borthwick KM, Davenport VA, Feemster JC, et al. Multisite validation of a simple electronic health record algorithm for identifying diagnosed obstructive sleep apnea. J Clin Sleep Med. 2020 Feb 15;16(2):175–83.

63. Levine Z, Kalka I, Kolobkov D, Rossman H, Godneva A, Shilo S, et al. Genome-wide association studies and polygenic risk score phenome-wide association studies across complex phenotypes in the human phenotype project. MED. 2024 Jan 12;5(1):90–101.e4.

