## Supplemental material for "Polygenic scores for obstructive sleep apnea based on BMI-adjusted and -unadjusted genetic associations reveal pathways contributing to cardiovascular disease"

#### Supplementary Figures

##### Supplementary Figure 1: Effect estimates from association analysis of PRS-CS based OSA PGSs (BMI-adjusted GWAS) comparing LD reference panel.

| case | control | OR [95% CI] | P | AUC |
|---|---|---|---|---|
| *Black (MVP GWAS)* | | | | |
| 1016 | 3738 | 1.11 [1.01;1.21] | 3.03e−02 | 0.79 |
| 1016 | 3738 | 1.17 [1.05;1.30] | 5.07e−03 | 0.79 |
| *White (MVP GWAS)* | | | | |
| 2270 | 7497 | 2.32 [1.81;2.98] | 4.51e−11 | 0.73 |
| 2270 | 7497 | 2.43 [1.91;3.10] | 7.40e−13 | 0.73 |
| *White (MVP+FINNGEN GWAS)* | | | | |
| 2270 | 7497 | 1.24 [1.15;1.34] | 1.93e−08 | 0.73 |
| 2270 | 7497 | 2.29 [1.89;2.77] | 1.97e−17 | 0.73 |
| *White (FINNGEN GWAS)* | | | | |
| 2270 | 7497 | 1.10 [1.05;1.15] | 6.14e−05 | 0.73 |
| 2270 | 7497 | 1.15 [1.09;1.21] | 7.90e−08 | 0.73 |
| *Hispanic/Latino(MVP GWAS)* | | | | |
| 1051 | 6087 | 1.03 [0.96;1.11] | 4.17e−01 | 0.83 |
| 1051 | 6087 | 1.04 [0.97;1.12] | 2.65e−01 | 0.83 |

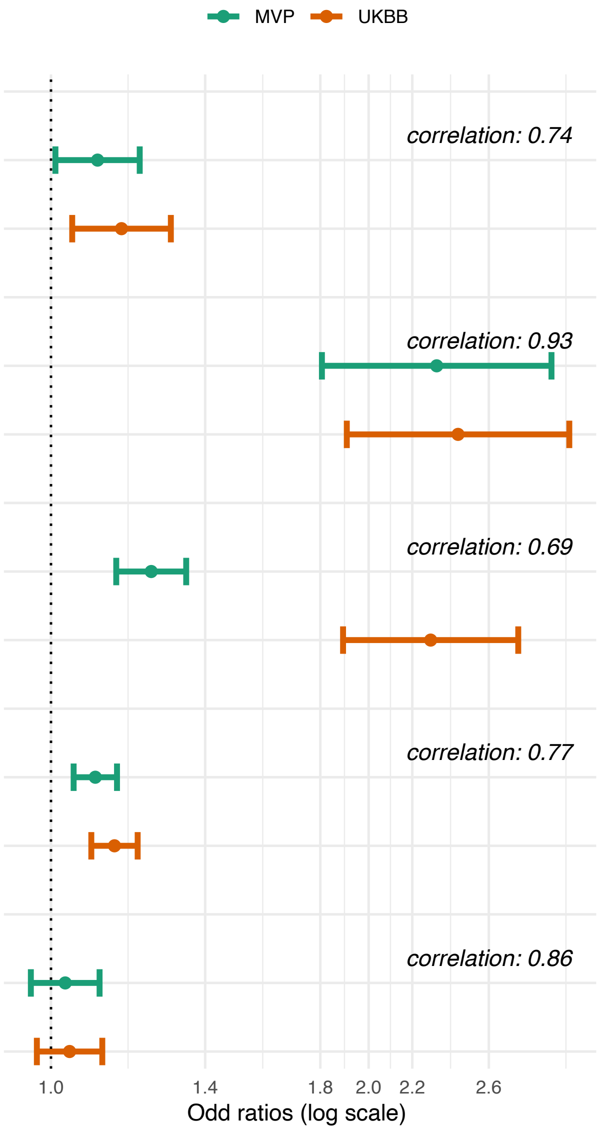

OSA was defined as AHI/REI≥15.
AHI: apnea hypopnea index; OSA: obstructive sleep apnea; REI: respiratory event index.

Supplementary Figure 2: Effect estimates from association analysis of PRS-CS based OSA PGSs (BMI-unadjusted GWAS) comparing LD reference panel.

| case | control | OR [95% CI] | P | AUC |
|---|---|---|---|---|
| *Black (MVP GWAS)* | | | | |
| 1016 | 3738 | 1.09 [0.99;1.19] | 6.59e−02 | 0.78 |
| 1016 | 3738 | 1.14 [1.03;1.25] | 7.73e−03 | 0.79 |
| *White (MVP GWAS)* | | | | |
| 2270 | 7497 | 2.02 [1.64;2.49] | 3.02e−11 | 0.73 |
| 2270 | 7497 | 1.96 [1.58;2.42] | 5.78e−10 | 0.73 |
| *White (MVP+FINNGEN GWAS)* | | | | |
| 2270 | 7497 | 1.16 [1.08;1.24] | 1.47e−05 | 0.73 |
| 2270 | 7497 | 1.68 [1.46;1.93] | 3.74e−13 | 0.73 |
| *White (FINNGEN GWAS)* | | | | |
| 2270 | 7497 | 1.10 [1.04;1.15] | 1.69e−04 | 0.73 |
| 2270 | 7497 | 1.14 [1.08;1.20] | 5.99e−07 | 0.73 |
| *Hispanic/Latino(MVP GWAS)* | | | | |
| 1051 | 6087 | 1.04 [0.96;1.13] | 3.65e−01 | 0.83 |
| 1051 | 6087 | 1.04 [0.95;1.14] | 3.72e−01 | 0.83 |

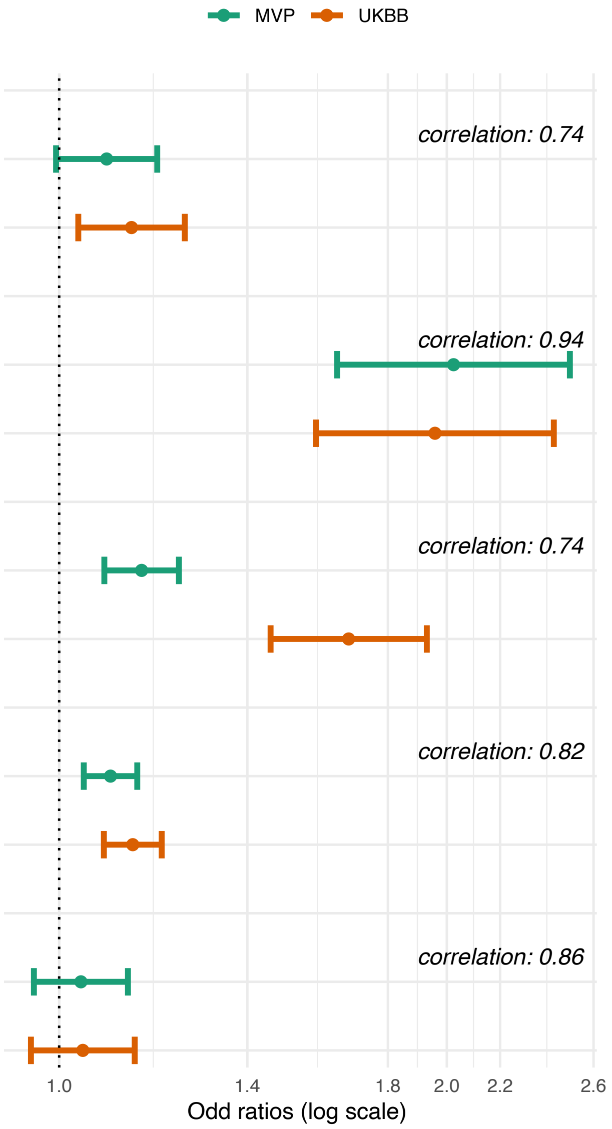

OSA was defined as AHI/REI≥15.
AHI: apnea hypopnea index; OSA: obstructive sleep apnea; REI: respiratory event index.

#### Supplementary Figure 3: Comparison of ancestry-specific OSA PGSs developed by PRS-CS and LDPred2 (BMI-adjusted analysis)

| case | control | OR [95% CI] | P | AUC |
|---|---|---|---|---|
| *Black (MVP GWAS)* | | | | |
| 1016 | 3738 | 1.17 [1.05;1.30] | 5.07e−03 | 0.79 |
| 1016 | 3738 | 1.14 [1.04;1.26] | 6.90e−03 | 0.79 |
| *White (MVP GWAS)* | | | | |
| 2270 | 7497 | 2.43 [1.91;3.10] | 7.40e−13 | 0.73 |
| 2270 | 7497 | 1.61 [1.35;1.92] | 1.10e−07 | 0.73 |
| *White (MVP+FINNGEN GWAS)* | | | | |
| 2270 | 7497 | 2.29 [1.89;2.77] | 1.97e−17 | 0.73 |
| 2270 | 7497 | 1.70 [1.48;1.95] | 1.04e−13 | 0.73 |
| *White (FINNGEN GWAS)* | | | | |
| 2270 | 7497 | 1.15 [1.09;1.21] | 7.90e−08 | 0.73 |
| 2270 | 7497 | 1.14 [1.09;1.20] | 1.06e−07 | 0.73 |
| *Hispanic/Latino(MVP GWAS)* | | | | |
| 1051 | 6087 | 1.04 [0.97;1.12] | 2.65e−01 | 0.83 |
| 1051 | 6087 | 1.04 [0.96;1.12] | 3.34e−01 | 0.83 |

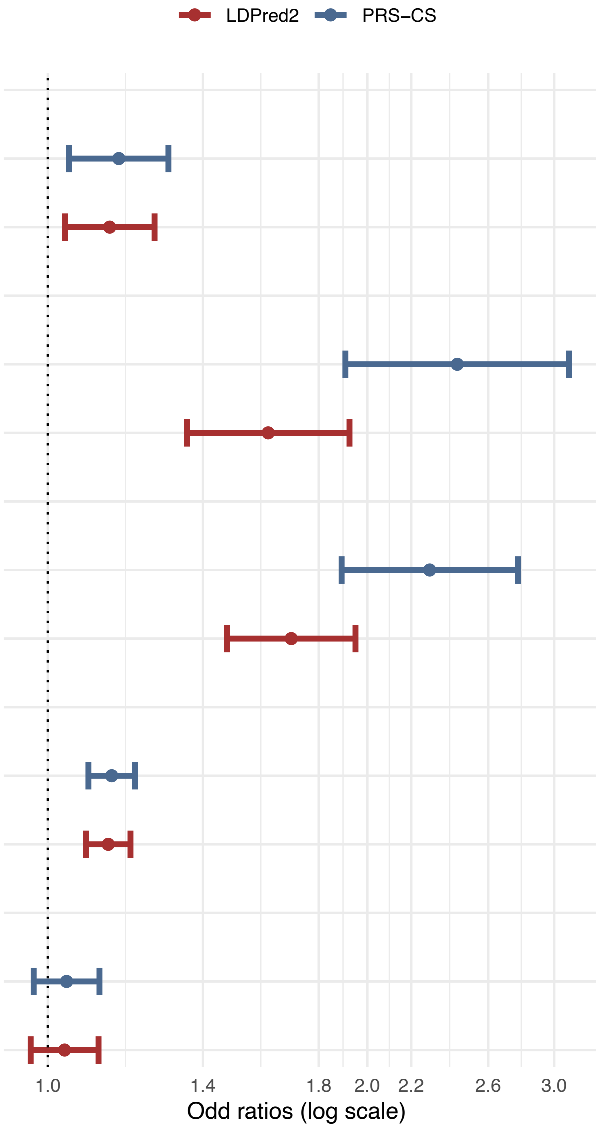

OSA was defined as AHI/REI≥15.
AHI: apnea hypopnea index; OSA: obstructive sleep apnea; REI: respiratory event index.

#### Supplementary Figure 4: Comparison of ancestry-specific OSA PGSs developed by PRS-CS and LDPred2 (BMI-unadjusted analysis)

| case | control | OR [95% CI] | P | AUC |
|---|---|---|---|---|
| *Black (MVP GWAS)* | | | | |
| 1016 | 3738 | 1.14 [1.03;1.25] | 7.73e−03 | 0.79 |
| 1016 | 3738 | 1.13 [1.02;1.25] | 1.83e−02 | 0.79 |
| *White (MVP GWAS)* | | | | |
| 2270 | 7497 | 1.96 [1.58;2.42] | 5.78e−10 | 0.73 |
| 2270 | 7497 | 1.44 [1.24;1.69] | 3.70e−06 | 0.73 |
| *White (MVP+FINNGEN GWAS)* | | | | |
| 2270 | 7497 | 1.68 [1.46;1.93] | 3.74e−13 | 0.73 |
| 2270 | 7497 | 1.28 [1.17;1.41] | 1.45e−07 | 0.73 |
| *White (FINNGEN GWAS)* | | | | |
| 2270 | 7497 | 1.14 [1.08;1.20] | 5.99e−07 | 0.73 |
| 2270 | 7497 | 1.11 [1.06;1.17] | 1.07e−05 | 0.73 |
| *Hispanic/Latino(MVP GWAS)* | | | | |
| 1051 | 6087 | 1.04 [0.95;1.14] | 3.72e−01 | 0.83 |
| 1051 | 6087 | 1.05 [0.97;1.14] | 2.00e−01 | 0.83 |

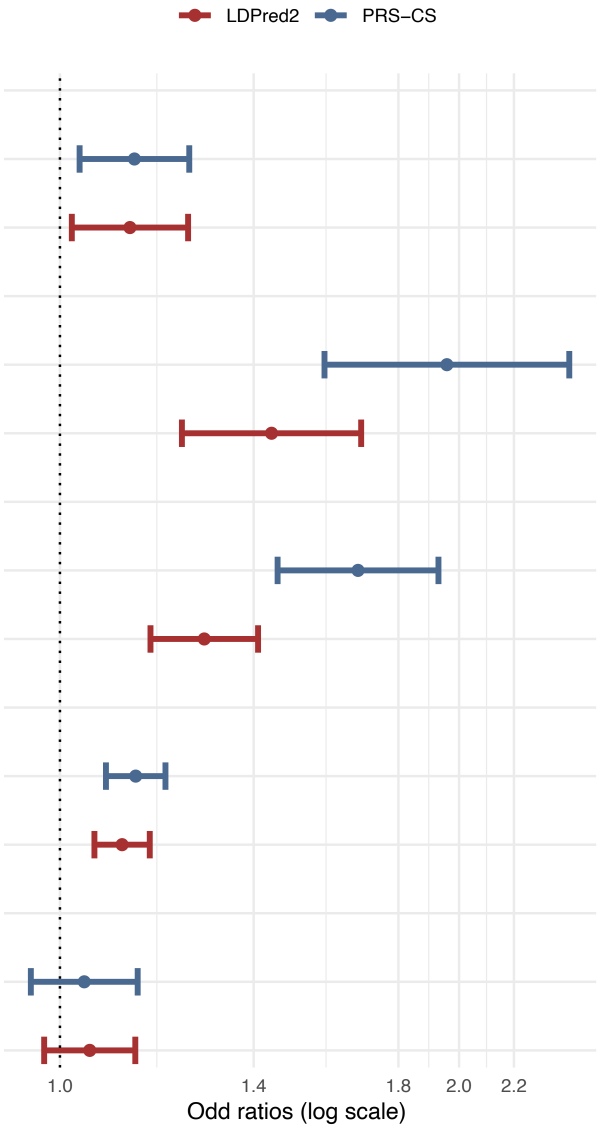

#### Supplementary Figure 5: Comparison of multi-population PGSs (BMI-adjusted GWAS)

| PRS method | OR [95% CI] | P | AUC |
|---|---|---|---|
| *Meta–Analsyis* | | | |
| *LDPred2:MVP* | 1.49 [1.34;1.66] | 2.31e−13 | 0.764 |
| *LDPred2:MVP+ FinnGen* | 1.47 [1.35;1.59] | 7.91e−19 | 0.765 |
| *Unweighted PRSsum* | | | |
| *LDPred2* | 1.19 [1.14;1.24] | 1.03e−15 | 0.788 |
| *PRS–CS* | 1.20 [1.15;1.25] | 3.44e−18 | 0.788 |
| *PRS–CSx* | 1.19 [1.13;1.24] | 1.45e−13 | 0.788 |
| *Weighted PRSsum* | | | |
| *LDPred2* | 1.51 [1.38;1.66] | 1.01e−18 | 0.790 |
| *PRS–CS* | 1.98 [1.74;2.24] | 1.61e−26 | 0.790 |
| *PRS–CSx* | 1.70 [1.54;1.88] | 2.46e−25 | 0.789 |

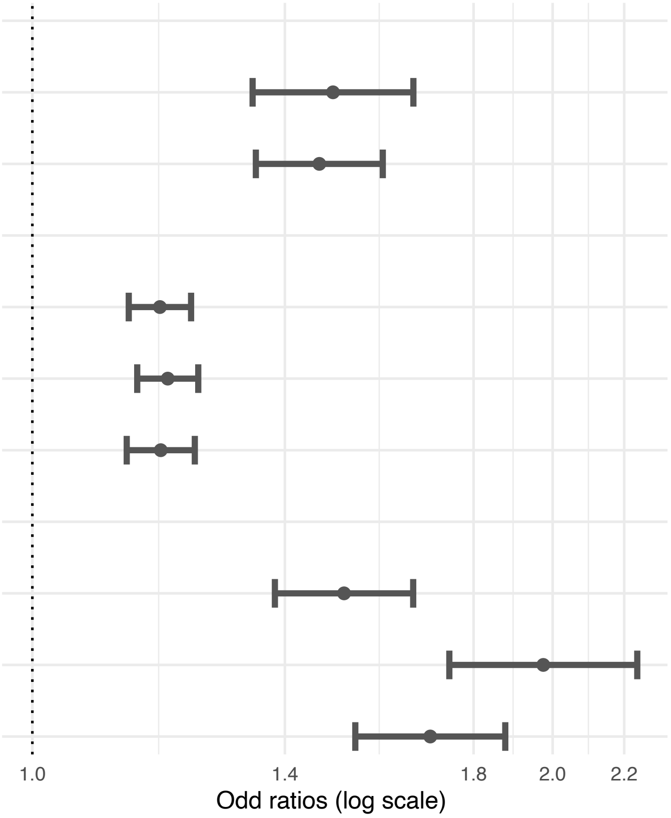

Comparison of PGS associations with OSA in TOPMed. PGS were constructed by different approaches to combine data across ancestry-specific GWAS. In all analyses, N cases=4,458 and N control= 17,466.
OSA was defined using AHI/REI>15, or using self-reported doctor diagnosis (only in participants from the COPD gene study). All analyses used both MVP and FinnGen summary statistics, other than LDPred2:MVP over meta-analysis which used only MVP data.
AHI: apnea hypopnea index; OSA: obstructive sleep apnea; REI: respiratory event index.

#### Supplementary Figure 6: Comparison of multi-population PGSs (BMI-unadjusted GWAS)

| PRS method | OR [95% CI] | P | AUC |
|---|---|---|---|
| *Meta–Analsyis* | | | |
| *LDPred2:MVP* | 1.22 [1.12;1.32] | 2.23e–06 | 0.762 |
| *LDPred2:MVP+HAN Chinese+FinnGen* | 1.17 [1.10;1.24] | 2.21e–07 | 0.762 |
| *Unweighted PRSsum* | | | |
| *LDPred2* | 1.11 [1.07;1.16] | 1.67e–07 | 0.787 |
| *PRS–CS* | 1.17 [1.12;1.23] | 2.63e–12 | 0.785 |
| *PRS–CSx* | 1.12 [1.07;1.18] | 8.04e–07 | 0.785 |
| *Weighted PRSsum* | | | |
| *LDPred2* | 1.25 [1.17;1.34] | 1.19e–10 | 0.787 |
| *PRS–CS* | 1.53 [1.39;1.68] | 2.51e–18 | 0.786 |
| *PRS–CSx* | 1.25 [1.17;1.33] | 2.78e–12 | 0.785 |

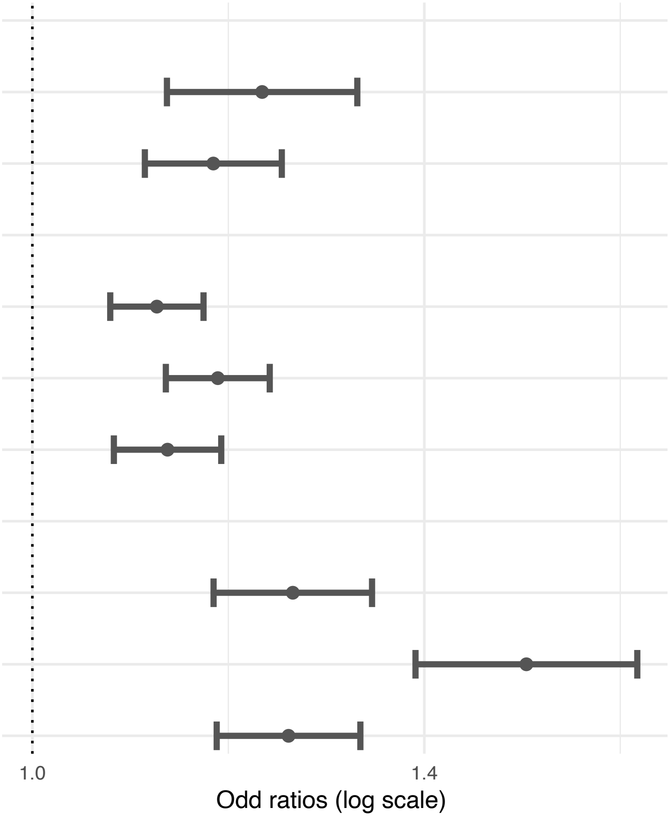

[results here: focusing on BMIunadj GWAS; BMI-adjusted analysis]
OSA was defined using AHI/REI>15, or using self-reported doctor diagnosis (only in participants from the COPD gene study). In all analyses N cases=4,458 and N control= 17,466.
All analyses used both MVP, FinnGen, and Han Chinese summary statistics, other than LDPred2:MVP over meta-analysis which used only MVP data.
AHI: apnea hypopnea index; OSA: obstructive sleep apnea; REI: respiratory event index

Supplementary Figure 7: comparison of estimated OSA associations with sex-specific and sex-combined OSA PGSs (BMI-adjusted).

| | case | control | OR [95% CI] | P | AUC |
|---|---|---|---|---|---|
| *Black* | | | | | |
| Female | 524 | 1852 | 1.52 [0.99;2.34] | 5.63e-02 | 0.77 |
| | | | 1.07 [0.94;1.22] | 3.11e-01 | 0.77 |
| Male | 492 | 1886 | 1.80 [1.15;2.80] | 9.75e-03 | 0.74 |
| | | | 1.70 [1.00;2.90] | 5.20e-02 | 0.74 |
| *White* | | | | | |
| Female | 812 | 4054 | 2.29 [1.77;2.98] | 4.97e-10 | 0.70 |
| | | | 1.15 [1.06;1.24] | 5.86e-04 | 0.70 |
| Male | 1458 | 3443 | 1.90 [1.53;2.36] | 8.34e-09 | 0.69 |
| | | | 1.88 [1.44;2.45] | 3.23e-06 | 0.68 |
| *Hispanic/Latino* | | | | | |
| Female | 438 | 3709 | 1.89 [1.30;2.73] | 7.63e-04 | 0.79 |
| | | | 1.03 [0.92;1.16] | 6.37e-01 | 0.79 |
| Male | 613 | 2378 | 2.33 [1.66;3.00] | 1.03e-06 | 0.81 |
| | | | 2.40 [1.58;3.00] | 4.04e-05 | 0.81 |
| *Multi-ethnic* | | | | | |
| Female | 1812 | 9713 | 2.02 [1.67;2.43] | 3.35e-13 | 0.76 |
| | | | 1.10 [1.04;1.17] | 8.75e-04 | 0.76 |
| Male | 2645 | 7753 | 1.97 [1.67;2.33] | 2.50e-15 | 0.77 |
| | | | 1.95 [1.59;2.39] | 1.49e-10 | 0.76 |

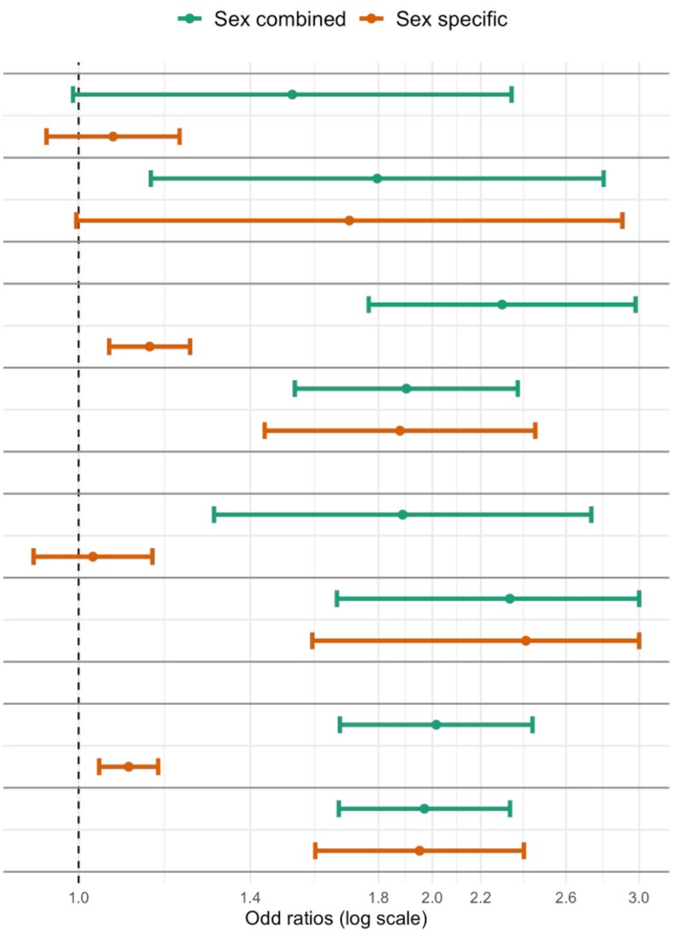

Associations of PGSs constructed using sex-specific and sex-combined analyses within their respective sex groups, in the combined and race/ethnicity-stratified TOPMed dataset. PGSs were constructed based on BMI-adjusted GWAS. Sex-combined PGS is the BMIadjOSA PGS selected as optimal in TOPMed.

**Supplementary Figure 8: comparison of estimated OSA associations with sex-specific and sex-combined OSA PGSs (BMI-unadjusted).**

| | case | control | OR [95% CI] | P | AUC |
|---|---|---|---|---|---|
| *Black* | | | | | |
| Female | 524 | 1852 | 1.46 [1.05;2.02] | 2.46e-02 | 0.77 |
| | | | 1.11 [0.97;1.26] | 1.18e-01 | 0.77 |
| Male | 492 | 1886 | 1.57 [1.10;2.24] | 1.20e-02 | 0.74 |
| | | | 1.64 [1.04;2.58] | 3.18e-02 | 0.74 |
| *White* | | | | | |
| Female | 812 | 4054 | 1.59 [1.31;1.94] | 4.42e-06 | 0.70 |
| | | | 1.16 [1.07;1.26] | 3.83e-04 | 0.70 |
| Male | 1458 | 3443 | 1.53 [1.30;1.81] | 3.32e-07 | 0.69 |
| | | | 1.69 [1.36;2.10] | 2.89e-06 | 0.68 |
| *Hispanic/Latino* | | | | | |
| Female | 438 | 3709 | 1.46 [1.09;1.94] | 1.01e-02 | 0.79 |
| | | | 1.09 [0.97;1.22] | 1.64e-01 | 0.79 |
| Male | 613 | 2378 | 1.53 [1.19;1.98] | 1.07e-03 | 0.80 |
| | | | 1.68 [1.20;2.36] | 2.64e-03 | 0.80 |
| *Multi-ethnic* | | | | | |
| Female | 1812 | 9713 | 1.53 [1.32;1.76] | 8.74e-09 | 0.76 |
| | | | 1.13 [1.07;1.20] | 3.70e-05 | 0.76 |
| Male | 2645 | 7753 | 1.53 [1.35;1.74] | 4.58e-11 | 0.77 |
| | | | 1.68 [1.42;1.99] | 1.76e-09 | 0.77 |

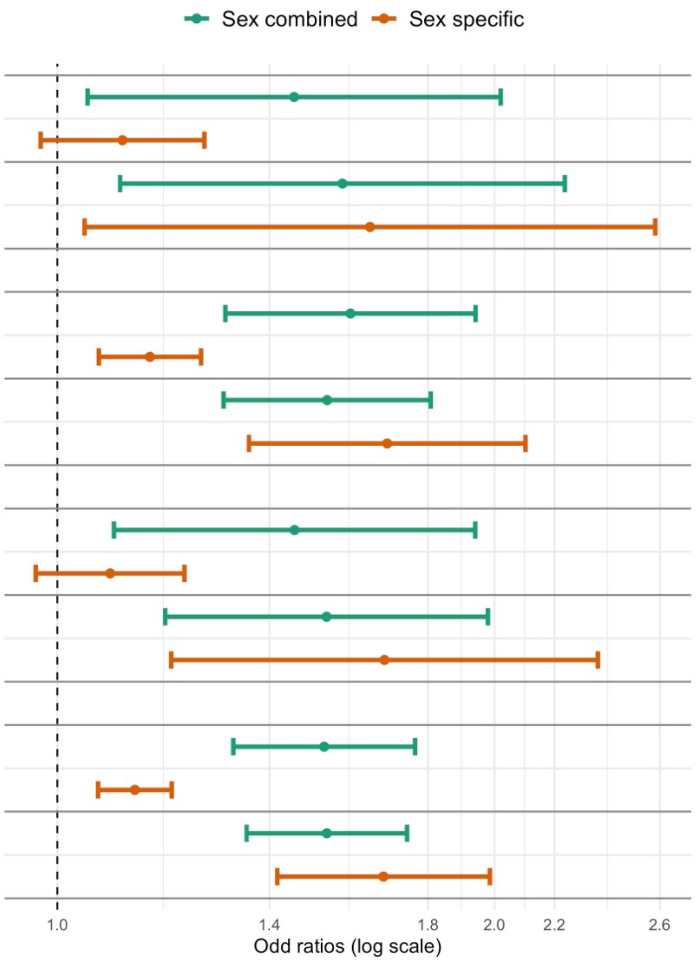

Associations of PGSs constructed using sex-specific and sex-combined analyses within their respective sex groups, in the combined and race/ethnicity-stratified TOPMed dataset. PGSs were constructed based on BMI-unadjusted GWAS. Sex-combined PGS is the BMIunadjOSA PGS selected as optimal in TOPMed.

Supplementary Figure 9: Estimated associations of OSA PGSs OSA in strata defined by sex (MyCode) and by sex, age, and obesity (All of Us).

| | case | control | OR [95% CI] | P | AUC |
|---|---|---|---|---|---|
| ***All of Us*** | | | | | |
| *Age group* | | | | | |
| ≤ 40 | 822 | 24353 | 1.50 [1.20;1.87] | 2.93e-04 | 0.82 |
| | | | 1.32 [1.11;1.57] | 2.00e-03 | 0.82 |
| 41-60 | 3105 | 27517 | 1.86 [1.65;2.10] | 1.26e-23 | 0.76 |
| | | | 1.50 [1.37;1.66] | 7.91e-17 | 0.76 |
| ≥ 61 | 3755 | 27127 | 1.91 [1.71;2.14] | 7.60e-30 | 0.73 |
| | | | 1.55 [1.42;1.69] | 8.89e-23 | 0.73 |
| *Sex* | | | | | |
| Male | 3569 | 30915 | 2.01 [1.79;2.25] | 3.94e-32 | 0.78 |
| | | | 1.57 [1.43;1.71] | 2.26e-22 | 0.78 |
| Female | 4113 | 48082 | 1.67 [1.51;1.85] | 8.76e-23 | 0.77 |
| | | | 1.44 [1.32;1.56] | 3.55e-18 | 0.77 |
| *Obesity* | | | | | |
| Non-Obese | 2513 | 51042 | 2.00 [1.76;2.28] | 3.23e-26 | 0.73 |
| | | | 1.57 [1.42;1.74] | 1.03e-18 | 0.73 |
| Obese | 4797 | 24431 | 1.70 [1.54;1.88] | 1.34e-25 | 0.71 |
| | | | 1.44 [1.33;1.56] | 2.51e-19 | 0.71 |
| ***MyCode*** | | | | | |
| *Sex* | | | | | |
| Male | 28592 | 10014 | 2.14 [1.97;2.33] | 1.88e-71 | 0.74 |
| | | | 1.78 [1.67;1.90] | 3.66e-72 | 0.75 |
| Female | 9134 | 49129 | 1.89 [1.75;2.05] | 1.11e-55 | 0.76 |
| | | | 1.70 [1.60;1.80] | 3.46e-66 | 0.76 |

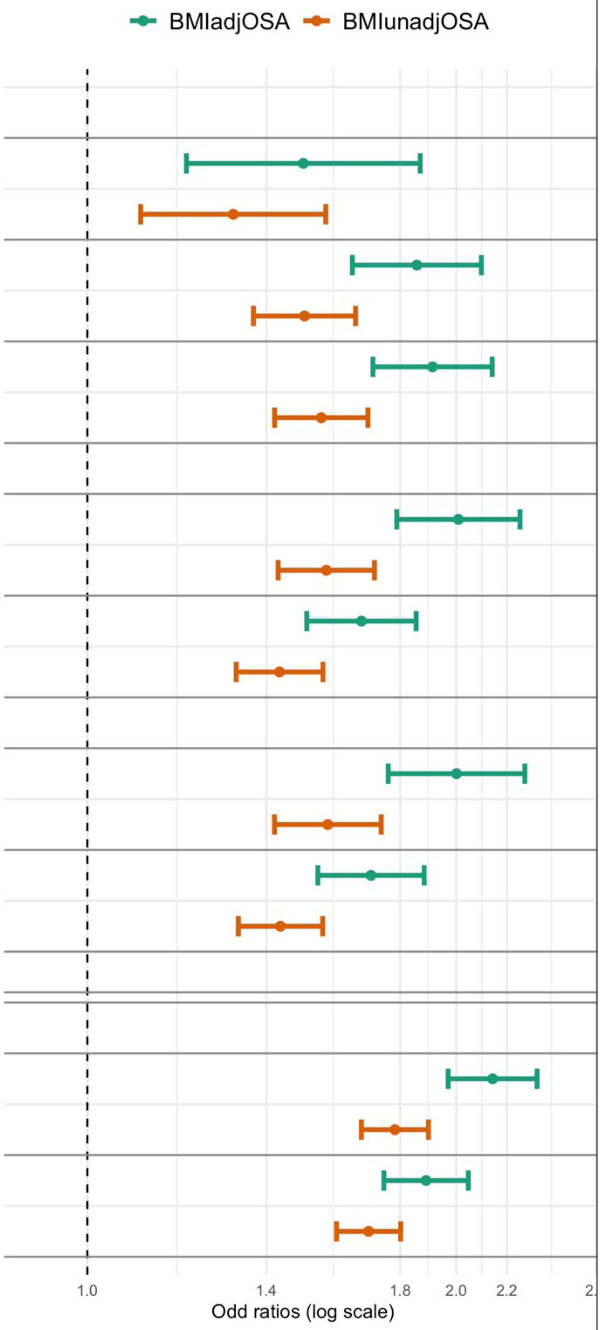

All of Us analyses were adjusted for age, sex, BMI (linear and squared terms) and 10 genetic PCs. MyCode analyses were adjusted for age, BMI (linear and squared terms), sex (in combined analysis), self-reported or EHR-derived race/ethnicity, and the top 20 genetic PCs.

Supplementary Figure 10: Estimated associations of OSA PGSs with oxygen saturation phenotypes in the Human Phenotypes Project.

| Trait | N | BETA [95% CI] | P |
|---|---|---|---|
| Min SatO2 | 4208 | -4.04e-03 [-6.23e-03;-1.84e-03] | 3.15e-04 |
| | | -2.65e-03 [-4.30e-03;-9.89e-04] | 1.76e-03 |
| Avg SatO2 | 4208 | -8.35e-04 [-1.31e-03;-3.58e-04] | 6.15e-04 |
| | | -4.65e-04 [-8.26e-04;-1.04e-04] | 1.15e-02 |

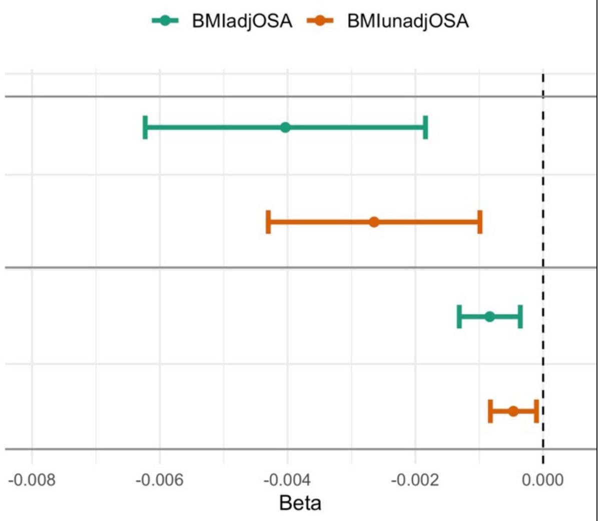

Estimated associations of the two OSA PGSs with log-transformed minimum and average oxyhemoglogin saturation measures during sleep (Min SatO2 and AvgSatO2), where the measures were averaged over the three nights of measurement. All analyses were adjusted for age, sex, BMI (linear and squared terms) and 10 genetic PCs.

Supplementary Figure 11: Estimated associations of OSA PGSs with sleep stage measures in the Human Phenotypes Project.

| Trait | N | BETA [95% CI] | P |
|---|---|---|---|
| % of deep sleep | 4239 | -0.01 [-0.46; 0.44] | 9.65e-01 |
| | | 0.23 [-0.11; 0.57] | 1.94e-01 |
| % of light sleep | 4239 | -0.76 [-1.64; 0.13] | 9.37e-02 |
| | | -0.96 [-1.63;-0.29] | 5.09e-03 |
| % of REM sleep | 4239 | 0.78 [ 0.17; 1.39] | 1.28e-02 |
| | | 0.72 [ 0.26; 1.19] | 2.20e-03 |
| Total deep sleep | 4242 | 0.01 [-0.02; 0.04] | 5.38e-01 |
| | | 0.02 [ 0.00; 0.04] | 1.15e-01 |
| Total light sleep | 4242 | 0.00 [-0.08; 0.08] | 9.34e-01 |
| | | -0.03 [-0.09; 0.03] | 3.23e-01 |
| Total sleep time | 4254 | 0.05 [-0.05; 0.15] | 2.98e-01 |
| | | 0.02 [-0.05; 0.10] | 5.68e-01 |

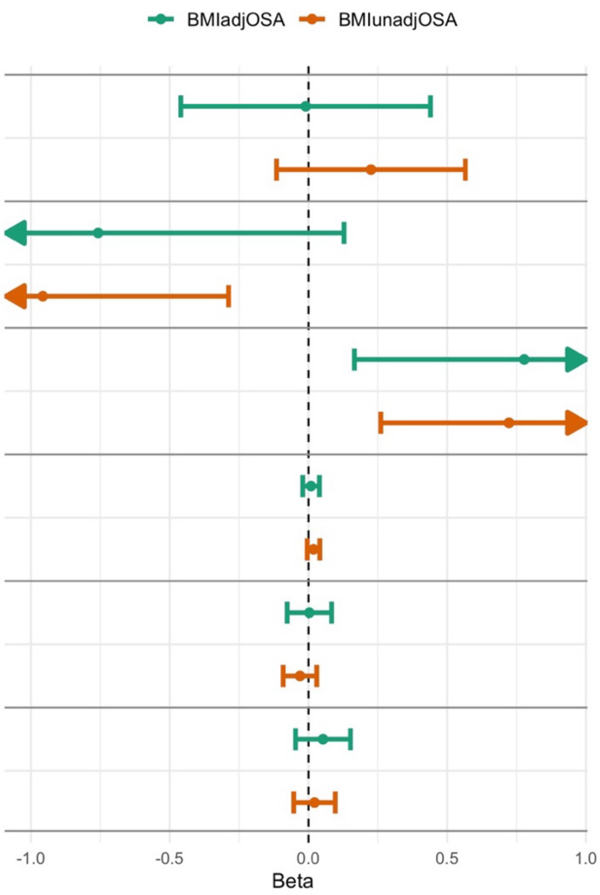

OSA PGS associations with sleep stage-related measures, averaged over the three nights of measurement. All analyses were adjusted for age, sex, BMI (linear and squared terms) and 10 genetic PCs.

#### Supplementary Figure 12: Estimated associations of OSA PGSs with self-reported sleep phenotypes in the Human Phenotypes Project.

| Trait | N | BETA [95% CI] | P |
|---|---|---|---|
| Sleep chronotype | 5845 | 0.05 [-0.07; 0.16] | 4.55e-01 |
| | | -0.01 [-0.10; 0.08] | 8.65e-01 |
| Daytime sleepiness | 5847 | 0.05 [ 0.00; 0.09] | 4.32e-02 |
| | | 0.01 [-0.02; 0.04] | 5.58e-01 |
| Sleep hours daily | 5772 | -0.05 [-0.13; 0.03] | 2.24e-01 |
| | | -0.07 [-0.13;-0.01] | 2.98e-02 |
| Sleep nap | 5840 | -0.01 [-0.07; 0.05] | 7.56e-01 |
| | | -0.04 [-0.08; 0.00] | 6.45e-02 |
| Sleep waking morning easy | 5843 | -0.06 [-0.13; 0.01] | 1.14e-01 |
| | | -0.02 [-0.08; 0.03] | 4.07e-01 |
| Insomnia | 5834 | 0.01 [-0.05; 0.08] | 6.92e-01 |
| | | 0.03 [-0.02; 0.08] | 2.68e-01 |

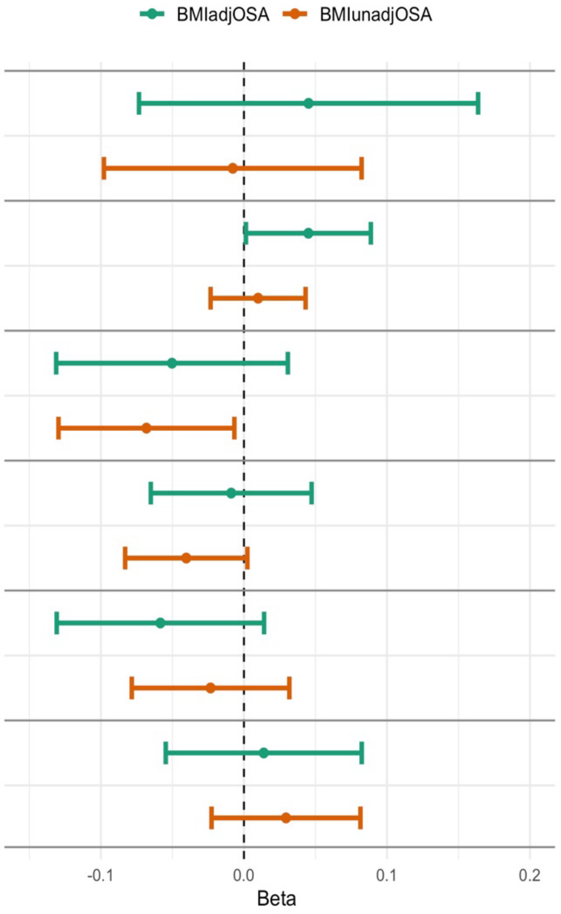

| Trait | case | control | OR [95% CI] | P | AUC |
|---|---|---|---|---|---|
| Insomnia | 85 | 5848 | 1.01 [0.48;2.11] | 9.85e-01 | 0.552 |
| | | | 1.30 [0.74;2.27] | 3.67e-01 | 0.567 |
| Short sleep | 389 | 5383 | 1.12 [0.79;1.60] | 5.20e-01 | 0.594 |
| | | | 1.07 [0.82;1.40] | 6.33e-01 | 0.595 |
| Long sleep | 27 | 5745 | 0.20 [0.05;0.73] | 1.47e-02 | 0.593 |
| | | | 0.35 [0.13;0.93] | 3.58e-02 | 0.636 |

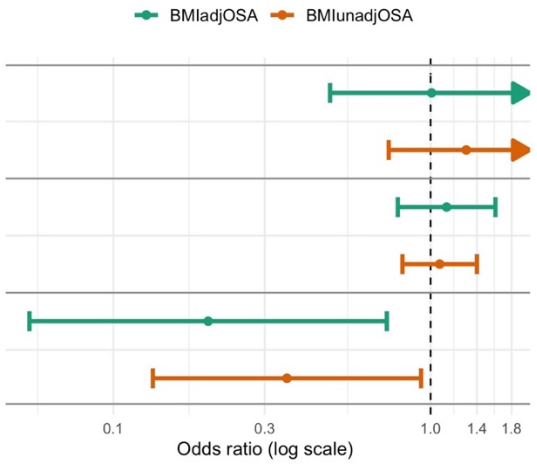

Excessive daytime sleepiness was assessed using the question “How likely are you to doze or fall asleep during day time you don’t mean to?  E.g. when working or driving?”. Insomnia/sleeplessness was assessed using the question “Do you have trouble falling sleep at night or do you wake up in the middle of the night?”. Sleep naps were assessed using the question “Do you have nap during the day?”. Easiness of waking up in the morning was

assessed using the question “on average day, how easy do you find getting up in the morning?”. Long sleep was defined as self-reported sleep duration>9, and short sleep was defined as self-reported sleep duration< 6.
All analyses were adjusted for age, sex, BMI (linear and squared terms) and 10 genetic PCs.

#### Supplementary Figure 13: Sex-stratified associations of OSA PGSs with clinical outcomes in All of Us

| | case | control | OR [95% CI] | P | AUC |
|---|---|---|---|---|---|
| *BMIunadjOSA* | | | | | |
| Stroke | 1159 | 33765 | 1.13 [0.98;1.31] | 8.71e-02 | 0.71 |
| | 1492 | 51393 | 1.17 [1.03;1.33] | 1.81e-02 | 0.69 |
| Atrial Fibrillation | 1957 | 33134 | 1.15 [1.02;1.29] | 1.95e-02 | 0.82 |
| | 1541 | 51720 | 1.18 [1.04;1.34] | 9.08e-03 | 0.79 |
| Heart Failure | 1408 | 33691 | 1.23 [1.08;1.41] | 2.14e-03 | 0.75 |
| | 1303 | 51854 | 1.17 [1.02;1.34] | 2.66e-02 | 0.80 |
| Coronary Artery Disease | 1316 | 33765 | 1.17 [1.02;1.34] | 2.32e-02 | 0.67 |
| | 1009 | 52347 | 1.14 [0.97;1.33] | 1.07e-01 | 0.75 |
| Asthma | 1921 | 33135 | 1.17 [1.05;1.31] | 5.51e-03 | 0.57 |
| | 5657 | 46521 | 1.10 [1.03;1.18] | 5.93e-03 | 0.62 |
| COPD | 1562 | 33608 | 1.15 [1.01;1.30] | 3.45e-02 | 0.71 |
| | 2049 | 51123 | 1.08 [0.97;1.21] | 1.79e-01 | 0.76 |
| Chronic Kidney Disease | 2275 | 32713 | 1.15 [1.03;1.28] | 1.19e-02 | 0.73 |
| | 2339 | 50647 | 1.18 [1.07;1.31] | 1.43e-03 | 0.71 |
| *BMIadjOSA* | | | | | |
| Stroke | 1159 | 33765 | 1.19 [0.99;1.43] | 5.72e-02 | 0.71 |
| | 1492 | 51393 | 1.14 [0.97;1.34] | 1.04e-01 | 0.69 |
| Atrial Fibrillation | 1957 | 33134 | 1.12 [0.97;1.30] | 1.31e-01 | 0.81 |
| | 1541 | 51720 | 1.11 [0.94;1.30] | 2.17e-01 | 0.79 |
| Heart Failure | 1408 | 33691 | 1.02 [0.86;1.20] | 8.36e-01 | 0.74 |
| | 1303 | 51854 | 0.98 [0.83;1.17] | 8.25e-01 | 0.80 |
| Coronary Artery Disease | 1316 | 33765 | 1.06 [0.89;1.26] | 5.25e-01 | 0.67 |
| | 1009 | 52347 | 1.00 [0.83;1.21] | 9.86e-01 | 0.75 |
| Asthma | 1921 | 33135 | 1.22 [1.06;1.41] | 5.54e-03 | 0.58 |
| | 5657 | 46521 | 1.16 [1.07;1.27] | 4.36e-04 | 0.62 |
| COPD | 1562 | 33608 | 0.98 [0.84;1.15] | 8.32e-01 | 0.71 |
| | 2049 | 51123 | 1.00 [0.87;1.15] | 9.71e-01 | 0.76 |
| Chronic Kidney Disease | 2275 | 32713 | 1.03 [0.90;1.18] | 6.40e-01 | 0.73 |
| | 2339 | 50647 | 1.06 [0.93;1.20] | 3.83e-01 | 0.71 |

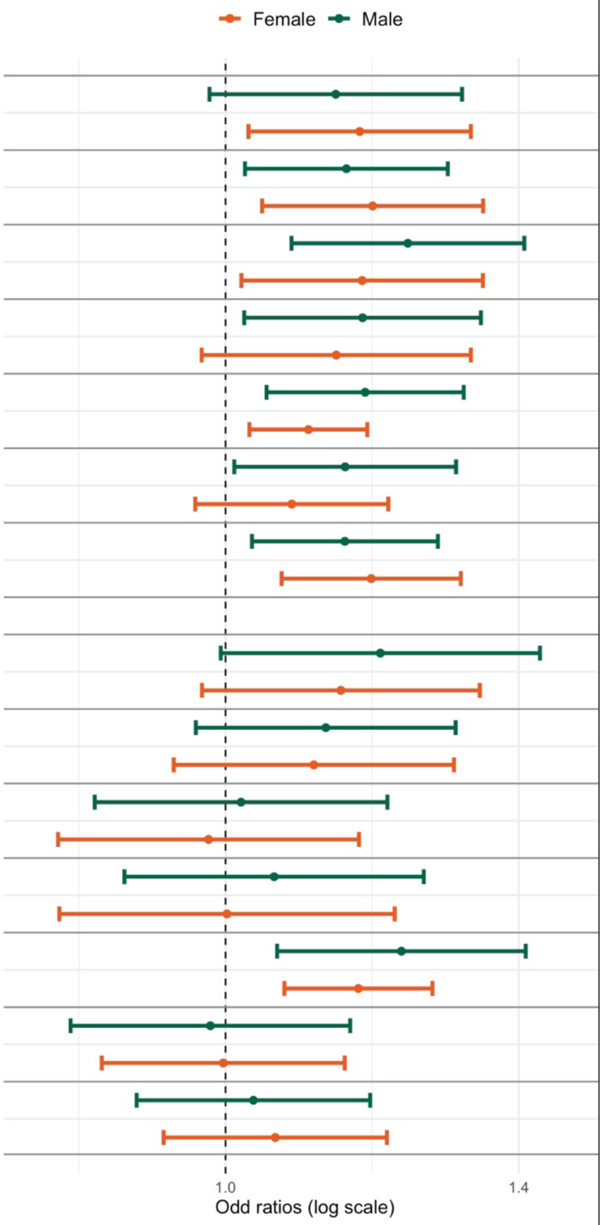

All analyses were adjusted for age, sex assigned at birth, BMI (linear and squared terms), and the first 10 genetic principal components.

#### Supplementary Figure 14: BMIunadjOSA PGS associations with DXA measures in the Human Phenotype Project

A.

| | N | BETA [95% CI] | P |
|---|---|---|---|
| VAT:SAT | 4541 | 3.51e-02 [8.55e-04;6.93e-02] | 4.46e-02 |
| | 2294 | 3.18e-02 [8.06e-03;5.56e-02] | 8.74e-03 |
| | 2247 | 4.10e-02 [-2.34e-02;1.06e-01] | 2.12e-01 |
| FM Gynoid: FM Android | 4603 | -1.26e-01 [-1.82e-01;-7.02e-02] | 9.62e-06 |
| | 2325 | -1.86e-01 [-2.79e-01;-9.25e-02] | 9.87e-05 |
| | 2278 | -5.99e-02 [-1.19e-01;-7.64e-04] | 4.72e-02 |

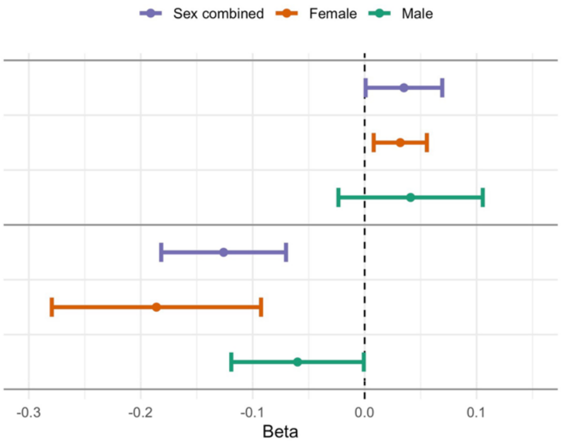

B.

| | N | BETA [95% CI] | P |
|---|---|---|---|
| VAT:TFM | 4527 | 2.54e-03 [1.39e-03;3.69e-03] | 1.51e-05 |
| | 2273 | 2.46e-03 [1.33e-03;3.59e-03] | 2.12e-05 |
| | 2254 | 2.66e-03 [6.66e-04;4.66e-03] | 9.02e-03 |
| SAT:TFM | 4545 | 1.33e-03 [2.62e-04;2.39e-03] | 1.46e-02 |
| | 2295 | 1.39e-03 [-8.51e-05;2.86e-03] | 6.49e-02 |
| | 2250 | 1.19e-03 [-3.37e-04;2.72e-03] | 1.26e-01 |
| FM Gynoid:TFM | 4605 | -3.02e-03 [-4.57e-03;-1.46e-03] | 1.43e-04 |
| | 2326 | -4.11e-03 [-6.40e-03;-1.82e-03] | 4.38e-04 |
| | 2279 | -1.75e-03 [-3.84e-03;3.51e-04] | 1.03e-01 |
| FM Android:TFM | 4605 | 3.18e-03 [1.99e-03;4.37e-03] | 1.68e-07 |
| | 2326 | 2.99e-03 [1.45e-03;4.53e-03] | 1.46e-04 |
| | 2279 | 3.34e-03 [1.52e-03;5.16e-03] | 3.41e-04 |

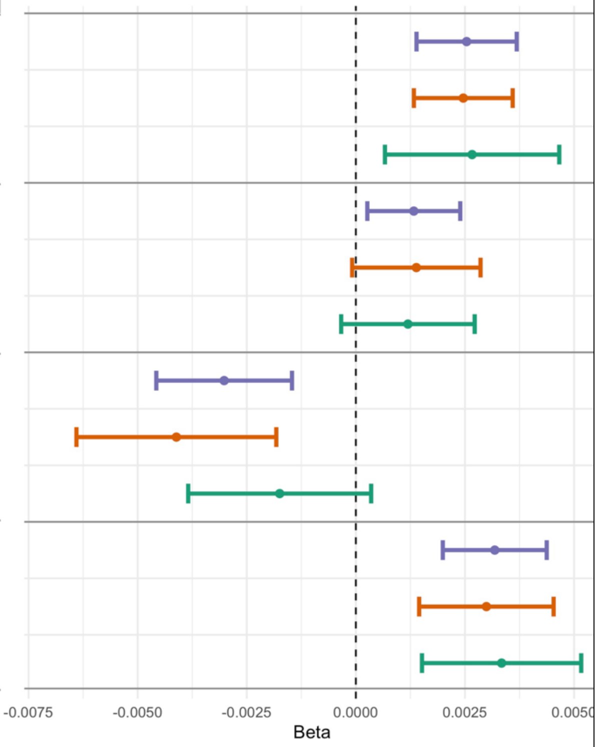

All associations were adjusted for age, sex, and the 10 first genetic principal components. BMI: body mass index; VAT: visceral adipose tissue; SAT: subcutaneous adipose tissue; FM: fat mass; TFM: total scan fat mass.

### Supplementary Tables

#### Supplementary Table 1: Characteristics of individuals participating in OSA PGS association analyses in TOPMed.

| | Black | Asian | White | Hispanic/Latino |
|---|---|---|---|---|
| **N** | 4760 | 230 | 9789 | 7196 |
| **Age (mean (SD))** | 56.38 (12.94) | 67.86 (9.11) | 62.67 (11.05) | 48.05 (14.63) |
| **BMI (mean (SD))** | 29.90 (7.00) | 24.27 (3.30) | 28.63 (5.92) | 30.07 (6.30) |
| **study (%)** | | | | |
| **ARIC** | 0 ( 0.0) | 0 ( 0.0) | 1108 (11.3) | 0 ( 0.0) |
| **CFS** | 506 (10.6) | 0 ( 0.0) | 487 ( 5.0) | 0 ( 0.0) |
| **CHS** | 200 ( 4.2) | 0 ( 0.0) | 687 ( 7.0) | 0 ( 0.0) |
| **COPDGene** | 2983 (62.7) | 0 ( 0.0) | 6322 (64.6) | 0 ( 0.0) |
| **FHS** | 0 ( 0.0) | 0 ( 0.0) | 485 ( 5.0) | 0( 0.0) |
| **HCHS/SOL** | 0 ( 0.0) | 0 ( 0.0) | 0 ( 0.0) | 6734 (93.6) |
| **JHS** | 579 (12.2) | 0 ( 0.0) | 0 ( 0.0) | 0 ( 0.0) |
| **MESA** | 492 (10.3) | 230 (100.0) | 700 ( 7.2) | 461 ( 6.4) |
| **Female sex (%)** | 2379 (50.0) | 115 ( 50.0) | 4876 (49.8) | 4183 (58.1) |
| **Moderate-to-severe OSA (%)** | 1020 (21.4) | 114 ( 49.6) | 2271 (23.2) | 1058 (14.7) |

Moderate-to-severe OSA was defined based on AHI or REI≥15.
Abbreviations: AHI: Apnea Hypopnea Index; BMI: Body Mass Index; OSA: Obstructive Sleep Apnea; REI: Respiratory Event Index.

Supplementary Table 2: Characteristics of individuals participating in OSA analysis in the MGB Biobank.

| | Black | Asian | White | Hispanic/Latino | Unknown |
|---|---|---|---|---|---|
| **N** | 2023 | 932 | 34764 | 1182 | 1292 |
| **Age (mean (SD))** | 53.16 (16.23) | 47.00 (15.67) | 60.62 (16.83) | 52.29 (16.56) | 52.08 (18.42) |
| **Sex F (%)** | 1267 (62.6) | 542 (58.2) | 18535 (53.3) | 812 (68.7) | 687 (53.2) |
| **BMI (mean (SD))** | 31.38 (7.64) | 25.15 (4.69) | 28.32 (6.31) | 29.80 (6.12) | 28.79 (6.33) |
| **OSA Yes (%)** | 345 (17.1) | 66 ( 7.1) | 4856 (14.0) | 117 ( 9.9) | 130 (10.1) |

MGB: Mass General Brigham; BMI: body mass index; OSA: obstructive sleep apnea.

#### Supplementary Table 3: TOPMed-evaluated mean, SDs, and combination weights, of PRS-CS based PGSs used in the final BMIadjOSA PGS and BMIunadjOSA PGS.

| Study | Mean | SD | PGS summation weight |
|---|---|---|---|
| **PGSs based on BMI-adjusted GWAS** | | | |
| **MVP-AFR** | 2.66e-07 | 1.46e-07 | 0.042 |
| **MVP-EUR** | 6.21e-07 | 1.95e-07 | 0.729 |
| **FinnGen** | -2.41E-08 | 4.95e-08 | 0.122 |
| **MVP-HIS** | -5.62E-07 | 1.27e-07 | 0.040 |
| **Final PGS sum** | 1.27e-11 | 7.32E-01 | |
| **PGSs based on BMI-unadjusted GWAS** | | | |
| MVP-AFR | 3.32e-07 | 1.59e-07 | 0.024 |
| **MVP-ASN + Han Chinese meta** | -1.94E-06 | 2.03e-07 | 0.043 |
| **MVP-EUR** | 3.96e-07 | 1.9e-07 | 0.566 |
| **FinnGen** | -1.10E-07 | 5.26e-08 | 0.120 |
| **MVP-HIS** | -1.14E-06 | 1.82e-07 | 0.037 |
| **Final PGS sum** | 9.14e-11 | 5.29E-01 | |

Means and SDs of the PGSs were computed based on the TOPMed dataset used in this study. PGS summation weights of the ancestry-specific PGSs were computed in a logistic regression of OSA over the PGSs, adjusted to age, sex, BMI, in the MGB biobank.
MPV: Million Veteran Program; AFR: MVP African HARE group; EUR: MVP European HARE group; HIS: MVP European HARE; ASN: MVP Asian HARE group; HARE: Harmonizing Genetic Ancestry and Self-identified Race/Ethnicity; PGS: polygenic score; MGB: Mass General Brigham.

Supplementary Table 4: OSA assessment methods in participating studies.

| Study | OSA assessment | Study role |
|---|---|---|
| **MVP** | Electronic health records based on ICD codes | GWAS to guide PGS development |
| **FinnGen** | Electronic health records-based extraction; subjective symptoms, clinical examination and sleep registration applying AHI ≥5 or REI ≥5. | |
| **MGB Biobank** | Electronic health records (OSA code) | Develop PGS combination weights |
| **ARIC** | Part of the Sleep Heart Health Study. The first visit took place between November 1995 and January 1998. At-home polysomnography using a Compumedics P Series system, Abbotsville, Victoria, Australia. OSA defined based on AHI. | TOPMed PGS evaluation |
| **CHS** | | |
| **FHS** | | |
| **CFS** | In primary analysis: type 3 home sleep apnea test (Edentrace; Eden Prairie, MN). OSA defined based on REI. In REM/NREM stratified analysis, data was used from a different examination (between 2000 and 2006), where sleep apnea was assessed using 14-channel overnight polysomnography (Compumedics E series, Abottsford, AU) obtained in a clinical research unit. | |
| **COPDGene** | Self-reported doctor-diagnosed OSA. | |
| **HCHS/SOL** | At-home overnight recording using an ARES Unicorder 5.2 (B-Alert, Carlsbad, CA). OSA defined based on REI. | |
| **JHS** | Type 3 home sleep apnea device (Embletta-Gold device, Embla, Broomfield, CO). OSA defined based on REI. | |
| **MESA** | At home polysomnography, Compumedics Somte Systems, Abbotsville, Australia, AU. OSA defined based on AHI. | |
| **All of Us** | Electronic health records (ICD codes) | Independent study validation or OSA PGSs; association analysis with other clinical outcomes |
| **Geisinger's MyCode Study** | Electronic health records (ICD codes) with a minimum of three encounter dates with OSA code. Controls had zero encounters. Patients with one or two OSA codes were excluded from analysis. | Independent study validation or OSA PGSs |
| **Human phenotype project** | WatchPAT device (itamar medical, Israel) applied over 3 nights, AHI averaged across the nights. OSA defined based on AHI. | Independent study validation or OSA PGSs; association analysis with other sleep measures |

Supplementary Table 5. Participants characteristics in analyses of OSA-related phenotypes (not REM/NREM stratified).

| | Black | Asian | White | Hispanic/Latino |
|---|---|---|---|---|
| **N** | 1777 | 230 | 3467 | 7195 |
| **Age (mean (SD))** | 59.24 (18.65) | 67.86 (9.11) | 63.60 (14.17) | 48.05 (14.64) |
| **Sex F (%)** | 1049 (59.0) | 115 ( 50.0) | 1832 (52.9) | 4183 (58.1) |
| **BMI(mean (SD))** | 31.18 (7.34) | 24.27 (3.30) | 28.56 (5.76) | 30.07 (6.30) |
| **study (%)** | | | | |
| **ARIC** | 0 ( 0.0) | 0 ( 0.0) | 1108 (32.0) | 0 ( 0.0) |
| **CFS** | 506 (28.5) | 0 ( 0.0) | 487 (14.0) | 0 ( 0.0) |
| **CHS** | 200 (11.3) | 0 ( 0.0) | 687 (19.8) | 0 ( 0.0) |
| **FHS** | 0 ( 0.0) | 0 ( 0.0) | 485 (14.0) | 0 ( 0.0) |
| **HCHS/SOL** | 0 ( 0.0) | 0 ( 0.0) | 0 ( 0.0) | 6734 (93.6) |
| **JHS** | 579 (32.6) | 0 ( 0.0) | 0 ( 0.0) | 0 ( 0.0) |
| **MESA** | 492 (27.7) | 230 (100.0) | 700 (20.2) | 461 ( 6.4) |
| **AHI/REI(mean (SD))** | 17.60 (20.55) | 20.37 (18.54) | 15.39 (16.85) | 7.71 (13.67) |
| **Avg SatO2 (mean (SD))** | 94.64 (2.70) | 94.92 (1.22) | 94.23 (2.23) | 96.28 (1.15) |
| **Min SatO2 (mean (SD))** | 84.29 (7.97) | 83.23 (7.56) | 85.17 (6.90) | 86.58 (6.48) |
| **Percent sleep < 90 saturation (mean (SD))** | 3.75 (10.44) | 2.24 (4.45) | 3.81 (10.65) | 1.09 (3.80) |

Supplementary Table 6. Participants characteristics in analyses of REM/NREM stratified OSA-related phenotypes.

| | Black | Asian | White | Hispanic/Latino |
|---|---|---|---|---|
| N | 1605 | 228 | 3213 | 460 |
| Age (mean (SD)) | 61.48 (17.13) | 67.82 (9.13) | 65.30 (12.15) | 68.45 (9.24) |
| Sex F (%) | 955 (59.5) | 113 ( 49.6) | 1716 (53.5) | 245 ( 53.3) |
| BMI (mean (SD)) | 31.00 (6.91) | 24.27 (3.29) | 28.39 (5.34) | 30.11 (5.46) |
| Study (%) | | | | |
| ARIC | 0 ( 0.0) | 0 ( 0.0) | 1108 (34.5) | 0 ( 0.0) |
| CFS | 337 (21.0) | 0 ( 0.0) | 235 ( 7.3) | 0 ( 0.0) |
| CHS | 200 (12.5) | 0 ( 0.0) | 687 (21.4) | 0 ( 0.0) |
| FHS | 0 ( 0.0) | 0 ( 0.0) | 485 (15.1) | 0 ( 0.0) |
| JHS | 578 (36.0) | 0 ( 0.0) | 0 ( 0.0) | 0 ( 0.0) |
| MESA | 490 (30.5) | 228 (100.0) | 698 (21.7) | 460 (100.0) |
| Avg DesatO2 (mean (SD)) | 3.42 (1.93) | 3.72 (1.78) | 2.63 (1.41) | 3.62 (2.13) |
| Avg DesatO2 NREM (mean (SD)) | 3.22 (1.98) | 3.50 (1.63) | 2.53 (1.35) | 3.33 (1.92) |
| Avg DesatO2 REM (mean (SD)) | 4.14 (3.19) | 5.05 (3.79) | 3.12 (2.39) | 5.04 (4.34) |
| Avg SatO2 NREM (mean (SD)) | 94.99 (2.07) | 95.02 (1.17) | 94.38 (1.85) | 94.49 (1.42) |
| Avg SatO2 REM (mean (SD)) | 94.36 (3.07) | 94.43 (1.76) | 94.07 (2.56) | 93.61 (2.63) |
| Min SatO2 NREM (mean (SD)) | 86.58 (6.26) | 85.40 (6.33) | 86.81 (5.51) | 84.77 (7.14) |
| Min SatO2 REM (mean (SD)) | 85.57 (7.69) | 84.79 (7.68) | 86.51 (6.45) | 82.52 (9.60) |
| AHI/REI NREM (mean (SD)) | 14.23 (19.27) | 18.38 (18.36) | 12.98 (15.87) | 18.91 (19.40) |
| AHI/REI REM (mean (SD)) | 27.74 (24.46) | 26.60 (21.01) | 22.43 (19.86) | 33.18 (23.11) |

##### Supplementary Table 7: Association of FEV1/FVC PGS with OSA continuous phenotypes

| Outcome | N | Estimated association | SE | P-value |
|---|---|---|---|---|
| **AHI/REI** | 12438 | 0.01 | 0.005 | 0.02 |
| **Hypoxic burden** | 10842 | 0.003 | 0.005 | 0.54 |
| **Min SatO2** | 12363 | -0.0006 | 0.0003 | 0.06 |
| **Avg SatO2** | 12359 | -0.0002 | 6.53E-05 | 0.0009 |

FEV1/FVC PGS weights were downloaded from the PGS catalog (PGP000244). Associations were adjusted for age, sex, BMI (both linear and squared terms), study, self-reported race/ethnicity, and 11 genetic PCs.

Supplementary Table 8: Geisinger's MyCode Community Health Initiative (MyCode) Study health system participating patient characteristics.

| | Black/African American | Asian | White | Hispanic/Latino | Other/Mixed |
|---|---|---|---|---|---|
| **N** | 2,557 | 663 | 89,684 | 3,289 | 676 |
| **Age (mean (SD))** | 47.95 (15.43) | 46.59 (15.89) | 59.26 (17.13) | 45.47 | 54.64 (15.88) |
| **Sex F (%)** | 1,555 (60.81) | 459 (69.23) | 53,528 (59.69) | 2,307 (70.14) | 414 (61.24) |
| **BMI (mean (SD))** | 32.55 (8.55) | 26.50 (5.23) | 30.59 (7.78) | 31.39 (7.61) | 30.47 (7.62) |
| **OSA Yes (%)** | 573 (22.41) | 65 (9.80) | 17,721 (19.76) | 635 (19.31) | 154 (22.78) |

#### Supplementary Table 9: Human Phenotypes Project participant characteristics.

| | Female | Male |
|---|---|---|
| **N** | 3,070 | 2,880 |
| **Age (mean(SD))** | 52.1(7.7) | 51.3(7.7) |
| **BMI (mean(SD))** | 25.6(4.3) | 26.5(3.7) |
| **OSA based on medical diagnosis Yes (%)** | 23(0.75) | 119 (4.1) |
| **Insomnia based on medical diagnosis Yes (%)** | 53(1.72) | 32(1.11) |

Supplementary Table 10: Human Phenotypes Project sleep monitoring participant characteristics.

| | **Female** | **Male** |
|---|---|---|
| **N** | 2609 | 2600 |
| **Age (mean(SD))** | 52.4(7.6) | 51.6(7.7) |
| **BMI (mean(SD))** | 25.7(4.4) | 26.8(3.8) |
| **AHI (mean(SD))** | 8.7(8.3) | 13.0(10.8) |
| **AHI-REM (mean(SD))** | 13.7(12.1) | 18.7(13.3) |
| **AHI-NREM (mean(SD)** | 7.1(7.6) | 11.2(10.6) |
| **ODI (mean(SD))** | 3.3(4.6) | 5.7(7.2) |
| **ODI-REM (mean(SD))** | 5.7(7.7) | 8.6(9.5) |
| **ODI-NREM (mean(SD))** | 2.5(4.0) | 4.8(6.9) |
| **RDI (mean(SD))** | 13.5(8.3) | 18.8(10.5) |
| **RDI-REM (mean(SD))** | 19.3(11.3) | 25.4(12.1) |
| **RDI-NREM (mean(SD))** | 11.5(8.0) | 16.7(10.8) |
| **MinSat O2 (mean(SD))** | 87.8(4.2) | 87.1(4.6) |
| **MaxSat O2 (mean(SD))** | 99.0(0.7) | 99.0(0.7) |
| **AvgSat O2 (mean(SD))** | 95.2(1.2) | 94.9(1.1) |
| **% of deep sleep time (mean(SD))** | 17.8(4.5) | 17.4(4.5) |
| **% of light sleep time (mean(SD))** | 57.8(8.8) | 59.3(8.7) |
| **% of rem sleep time (mean(SD))** | 24.4(5.9) | 23.3(6.1) |
| **Total deep sleep time (mean(SD))** | 1.1(0.3) | 1.0(0.3) |
| **Total light sleep time (mean(SD))** | 3.6(0.8) | 3.5(0.8) |
| **Total sleep time (mean(SD))** | 6.2(1.0) | 5.9(1.0) |
| **OSA (AHI > = 15) Yes (%)** | 449(17.2) | 797(30.7) |

Supplementary Table 11: Characteristics of Human Phenotype Projects participants and their self-reported sleep phenotypes.

| | Female | Male |
|---|---|---|
| **N** | 3262 | 2950 |
| **Age (mean(SD))** | 52.1(7.8) | 51.3(7.8) |
| **BMI (mean(SD))** | 25.7(4.4) | 26.5(3.8) |
| **Total sleep duration (mean(SD))** | 6.9(0.9) | 6.6(0.9) |
| **Long sleep Yes(%)** | 19(0.58) | 8(0.28) |
| **Short sleep Yes (%)** | 182(5.58) | 241(8.17) |
| **Insomnia/sleeplessness (%)** | | |
| **Never / rarely** | 1263(20,44) | 1484(24.01) |
| **Sometime** | 1193(19.3) | 964(15.60) |
| **Usually** | 781 (12.63) | 487(7.89) |
| **Prefer not to answer** | 6 (0.1) | 2(0.03) |
| **Easy to wake up in the morning (%)** | | |
| **Not at all easy** | 209(3.38) | 85(1.37) |
| **Not very easy** | 714(11.54) | 503(8.12) |
| **Fairly easy** | 1468(23.72) | 1439(23.25) |
| **Very easy** | 844(13.64) | 907(14.66) |
| **Prefer not to answer** | 3(0.05) | 2(0.03) |
| **Do not know** | 11(0.18) | 4(0.06) |
| **Sleep chronotype %** | | |
| **Definitely morning person** | 975(15.75) | 718(11.60) |
| **More a morning than evening person** | 1024(16.54) | 885(14.30) |
| **More an evening than a morning person** | 632(10.21) | 660(10.66) |
| **Definitely evening person** | 325(5.25) | 336(5.42) |
| **Prefer not to answer** | 5(0.08) | 3(0.05) |
| **Do not know** | 289(4.67) | 338(5.42) |
| **Sleep nap (%)** | | |
| **Never / rarely** | 2103(34.00) | 1956(31.62) |
| **Sometime** | 851(13.76) | 783(12.65) |
| **Usually** | 289((4.67) | 200(3.23) |
| **Prefer not to answer** | 3(0.04) | 1(0.01) |
| **Excessive daytime sleepiness (%)** | | |
| **Never/rarely** | 2673(43.16) | 2300(37.14) |
| **Sometime** | 461(7.44) | 524(8.46) |
| **Often** | 98(1.58) | 104(1.68) |
| **Prefer not to answer** | 1(0.01) | 1(0.01) |
| **Do not know** | 18(0.3) | 12(0.21) |

Excessive daytime sleepiness was assessed using the question “How likely are you to doze or fall asleep during day time you don’t mean to? E.g. when working or driving?”. Insomnia/sleeplessness was assessed using the question “Do you have trouble falling sleep at night or do you wake up in the middle of the night?”. Sleep naps were assessed using the question “Do you have nap during the day?”. Easiness of waking up in the morning was assessed using the question “on average day, how easy do you find getting up in the morning?”. Long sleep was defined as self-reported sleep duration>9, and short sleep was defined as self-reported sleep duration< 6.

#### Supplementary Table 12: Characteristics of individuals participating in All of Us association analyses.

| | Black | Asian | White | Hispanic/Latino | Other | Unknown |
|---|---|---|---|---|---|---|
| **N** | 18894 | 2843 | 46903 | 17555 | 552 | 3858 |
| **Age (mean (SD))** | 49.60 (14.72) | 44.45 (17.03) | 56.33 (16.89) | 45.86 (15.84) | 45.95 (17.04) | 50.38 (17.36) |
| **BMI (mean (SD))** | 30.32 (7.35) | 25.04 (4.63) | 28.53 (6.29) | 30.08 (6.29) | 28.21 (6.12) | 28.88 (6.58) |
| **Sex at birth F(%)** | 10366 (54.9) | 1694 (59.6) | 27855 (59.4) | 11961 (68.1) | 284 (51.4) | 2236 (58.0) |
| **OSA Yes(%)** | 1757 ( 9.3) | 175 ( 6.2) | 7228 (15.4) | 1430 ( 8.1) | 53 ( 9.6) | 466 (12.1) |
| **T2D Yes(%)** | 2856 (15.1) | 260 ( 9.1) | 5791 (12.3) | 2675 (15.2) | 57 (10.3) | 532 (13.8) |
| **AFib Yes(%)** | 525 ( 2.8) | 53 ( 1.9) | 3493 ( 7.4) | 548 ( 3.1) | 25 ( 4.5) | 206 ( 5.3) |
| **CAD Yes(%)** | 751 ( 4.0) | 53 ( 1.9) | 2032 ( 4.3) | 510 ( 2.9) | 22 ( 4.0) | 169 ( 4.4) |
| **Stroke Yes(%)** | 818 ( 4.3) | 66 ( 2.3) | 2684 ( 5.7) | 699 ( 4.0) | 20 ( 2.7) | 200 ( 5.2) |
| **Asthma Yes(%)** | 1963 (10.4) | 170 ( 6.0) | 5530 (11.8) | 1927 (11.0) | 44 ( 8.0) | 515 (13.3) |
| **CKD Yes(%)** | 1415 ( 7.5) | 80 ( 2.8) | 3469 ( 7.4) | 1033 ( 5.9) | 22 ( 4.0) | 329 ( 8.5) |
| **HF Yes(%)** | 1043 ( 5.5) | 51 ( 1.8) | 2288 ( 4.9) | 591 ( 3.4) | 17 ( 3.1) | 205 ( 5.3) |
| **HTN Yes(%)** | 13044 (69.5) | 1358 (48.2) | 30174 (64.9) | 10112 (57.9) | 0 | 0 |
| **COPD Yes(%)** | 1126 ( 6.0) | 30 ( 1.1) | 3018 ( 6.4) | 548 ( 3.1) | 19 ( 3.4) | 249 ( 6.5) |

#### Supplementary Table 13: Comparison of newly developed and existing OSA PGS associations with OSA in All of Us.

| Study | GWAS adjustment | Method | PMID | OR and 95% CI | P-value | AUC |
|---|---|---|---|---|---|---|
| **BMIunadjOSA** | BMI unadjusted | Weighted PGS summation of PRS-CS generated PGS | Current manuscript | 1.49 [1.40;1.58] | 2.72e-38 | 0.76 |
| **BMIadjOSA** | BMI adjusted | Weighted PGS summation of PRS-CS generated PGS | Current manuscript | 1.82 [1.68;1.96] | 1.12e-52 | 0. 76 |
| **Sofer & Kurniansyah et al.** | BMI unadjusted | Genome-wide significant SNPs | 36989840 | 1.61 [1.27; 2.06] | 9.03e-05 | 0.76 |
| **Sofer & Kurniansyah et al.** | BMI adjusted | Genome-wide significant SNPs | 36989840 | 1.21 [0.76; 1.90] | 0.41 | 0.76 |
| **Zhang et al** | BMI unadjusted | LDPred2 | 36174398 | 1.26 [1.18;1.36] | 1.78e-11 | 0.76 |

N cases: 7,682, N controls: 78,997.

All association models were adjusted for sex assigned at birth, age, BMI (linear and squared terms), the first 10 PCs of genetic ancestry.

Abbreviations. AIC: area under the (receiver operating) curve; BMI: body mass index; CI: confidence interval; OR: odds ratio; OSA: obstructive sleep apnea; PGS: polygenic score.

AUC was the same for all PGSs when rounding to the second digit.

#### Supplementary Table 14: Characteristics of DXA measures in the Human Phenotype Project

| | Female | Male |
|---|---|---|
| **Age (mean(SD))** | 52.2 (7.5) | 51.7 (7.7) |
| **BMI (mean(SD))** | 25.6 (4.3) | 26.8 (3.8) |
| **VAT fat mass ((mean(SD)))** | 0.63 (0.44) | 1.22 (0.74) |
| **SAT fat mass (mean(SD))** | 1.59 (0.75) | 1.41 (0.70) |
| **Android fat mass (mean(SD))** | 2.05 (0.94) | 2.34 (1.11) |
| **Gynoid fat mass (mean(SD))** | 4.73 (1.42) | 3.49 (1.20) |
| **Total fat mass (mean(SD))** | 25.83 (8.47) | 23.32 (8.13) |
| **VAT: Total fat mass (mean(SD))** | 0.02 (0.01) | 0.05 (0.02) |
| **SAT: Total fat mass (mean(SD))** | 0.06 (0.01) | 0.06 (0.01) |
| **FM Gynoid: Total fat mass (mean(SD))** | 0.19 (0.02) | 0.15 (0.02) |
| **FM Android: Total fat mass (mean(SD))** | 0.08 (0.02) | 0.10(0.02) |
| **VAT: SAT (mean(SD))** | 0.40 (0.25) | 0.93 (0.62) |
| **FM Gynoid: FM Android (mean(SD))** | 2.62 (0.93) | 1.67 (0.59) |

Fat masses are in kilograms.
BMI: body mass index; VAT: visceral adipose tissue; SAT: subcutaneous adipose tissue; FM: fat mass.

##### Supplementary Table 15: FinnGen participant characteristics.

| | All N=412,181 | OSA N=43,901 | Non-OSA N=368,280 |
|---|---|---|---|
| **Sex (male)** | 181,871 | 26,199 | 155,672 |
| **Age (SD)** | 60.3 (18.9) | 64.4 (12.6) | 59.9 (18.4) |
| **BMI (SD)** | 27.3 (5.5) | 31.7 (6.6) | 26.8 (5.1) |

Baseline characteristics of the individuals with and without obstructive sleep apnea (OSA) diagnosis in FinnGen.

Supplementary Table 16: TOPMed studies contributing to OSA PGS association with each of the non-OSA sleep phenotypes

| Sleep phenotype | Participating studies |
|---|---|
| **Phenotypes measured using sleep apnea device** | |
| **AHI/REI*** | ARIC, CHS, FHS, CFS, HCHS/SOL, JHS, MESA<br>During REM and NREM: ARIC, CHS, FHS, MESA, CFS (subset) |
| **Hypoxic burden** | ARIC, CHS, FHS, CFS, HCHS/SOL, JHS, MESA |
| **Minimum, average oxyhemoglobin saturation during sleep** | ARIC, CHS, FHS, CFS, HCHS/SOL, JHS, MESA |
| **Average oxyhemoglobin desaturation during respiratory events** | ARIC, CHS, FHS, CFS, HCHS/SOL, JHS, MESA |
| **Percent sleep time with oxyhemoglobin saturation<90%** | ARIC, CHS, FHS, CFS, HCHS/SOL, JHS, MESA |
| **Self-reported phenotypes** | |
| **Sleep duration, long, short sleep** | ARIC, CHS, FHS, CFS, HCHS/SOL, JHS, MESA, WHI |
| **Epworth sleepiness scale, excessive daytime sleepiness** | ARIC, CHS, FHS, CFS, HCHS/SOL, JHS, MESA |
| **Women Health Initiative insomnia rating scale, insomnia** | CHS, HCHS/SOL, JHS, MESA, WHI |

*HCHS/SOL, measured REI while other studies measured AHI.
AHI: Apnea Hypopnea Index; REI: Respiratory Event Index.

### Supplementary Note 1: Choosing optimal OSA PGSs in analysis in TOPMed.

#### Using UK Biobank as a reference panel is sufficient for developing PGS based on MVP summary statistics

We studied whether using UK Biobank reference panel to develop PGSs based on summary statistics from MVP (and potentially other studies, via meta-analysis) is sufficient. Supplementary Figures 1 and 2 visualizes the associations of ancestry-specific PGS developed using PRS-CS with OSA in TOPMed individuals of similar ancestry as the GWAS dataset used for PGS derivation. It compares the estimated associations when using MVP and when using UKBB -matched reference panels, i.e., EUR HARE group (MVP) or EUR participants (UKBB) for European ancestries PGS, AFR HARE group (MVP) or AFR participants (UKBB) for African ancestries PGS, and HIS HARE group (MVP) or AMR participants (UKBB) for Hispanic/Latino PGS. Note that Asian-specific PGS was not derived due to the low sample size of participants of East Asian ancestries. However, it was later derived as part of PRS-CSx multi-ancestry derivation as explained later. One can see that PGS that were derived using UKBB participants for LD reference panels performed well and sometimes better than PGSs that used MVP participants for LD refence panels. This is likely related to the imputation quality of the SNPs. Thus, when using the PRS-CS and the PRS-CSx software in multi-PGS methods next, we only focus on PGSs developed using the UKBB reference panels.

#### Identifying best performing single-ancestry PGSs

Supplementary Figures 3 and 4 compares single-ancestry PGS constructed using PRS-CS (UKBB reference panel) and using LDPred2 (MVP reference panel) in association with OSA in their respective ancestry group in the TOPMed dataset. The results are similar between LDPred2 and PRS-CS. We therefore move forward with PRS-CS for multi-PGS methods.

##### Multi-ethnic GWAS PGS versus a multi-PGS combining ancestry-specific PGSs

Supplementary Figure 5 compares the OSA associations of a PGS constructed based on a multi-ethnic GWAS, where we meta-analyzed MVP and FinnGen, to multi-PGS, i.e. PGS summations combining PGSs developed using PRS-CSx (in a joint model of a few ancestries), using LDPred2 (each genetic ancestry group PGS was developed separately), and using PRS-CS (each genetic ancestry group PGS was developed separately). Associations are provided in the combined and in separate race/ethnic groups. The weighted sum of PRS-CS constructed PGSs had the strongest associations with OSA. This is true also when constructing PGS based on BMI-unadjusted analysis (Supplementary Figure 6). We proceed with these PGSs as the best performing OSA PGSs, and we refer to them as BMIadjOSA-PGS and BMIunadjOSA-PGS.

#### Supplementary Note 2: MGB biobank

Samples, genomic data, and health information were obtained from the Mass General Brigham Biobank, a biorepository of consented patient samples at Mass General Brigham. Phenotypic data was extracted from the MGB Biobank on November 14, 2021. OSA was status was extracted as “Obstructive sleep apnea OSA existence” yes/no, and BMI was the median BMI value available in the records.

**DNA samples**

DNA samples are processed from whole blood that was collected as a dedicated research draw or as a clinical discard. Dedicated research samples are aimed to be processed within four hours of collection. Clinical discards are processed 24+ hours after collection. Whole blood is spun to buffy coat with a centrifuge and the buffy coat is stored in a freezer up to several months. The buffy coat is then extracted to DNA. The DNA is then placed in an ultralow freezer (-80ºC).Each DNA aliquot contains a minimum of 2 ug of DNA. The concentration varies.

**Genotyping**

Samples have been genotyped using three versions of the biobank SNP array offered by Illumina that is designed to capture the diversity of genetic backgrounds across the globe. The first batch of data was generated on the Multi-Ethnic Genotyping Array (MEGA) array, the first release of this SNP array. The second, third, and fourth batches were generated on the Expanded Multi-Ethnic Genotyping Array (MEGA Ex) array. All remaining data were generated on the Multi-Ethnic Global (MEG) BeadChip.

**Imputation**

Prior to performing imputation, files were converted to VCF format, separated by chromosomes. When multiple probes measured the same genotypes, they were checked for concordance and were set to a missing value if the genotypes did not match. Files were uploaded to the Michigan Imputation Server, and Genotypes were imputed using TOPMed reference panel. Genomic coordinates are provided in GRCh38.

**Quality control**

We performed quality control using PLINK (v2.0. We filtered SNPs with low-quality imputation (r < 0.5), with missing call rates > 0.1, HWE p-value less than $1x10^{-6}$ and MAF <1%.

We computed principal component (PC) using PLINK: we pruned the genotype data using a window size of 1000 variants, sliding across the genome with a step size of 250 variants at a time, filtering out any SNPs with LD $R^2$>0.1. We used unrelated individuals (3rd degree, identified using PLINK) to compute the loadings for the first 10 PCs.

**PGS construction**

We constructed all PGS using PRSice 2 (1), using the same SNPs as those selected in the various developed PGS, i.e. without further clumping. PGSs were scaled using the same mean and SD as the TOPMed dataset, so that PGS summation weights will be appropriate for TOPMed.

**Ethics statement**

All Biobank subjects have provided their consent to join the Partners Biobank, which includes agreeing to provide a blood sample linked to the electronic medical record. Subjects also agree to be recontacted by the Partners Biobank staff as needed.

**Acknowledgements**

We thank Mass General Brigham Biobank for providing samples, genomic data, and health information data.

#### Supplementary Note 3: TOPMed parent study descriptions

##### ARIC

The Atherosclerosis Risk in Communities study (dbGaP accession phs000090) is a population-based prospective cohort study of cardiovascular disease sponsored by the National Heart, Lung, and Blood Institute (NHLBI). ARIC included 15,792 individuals, predominantly European American and African American, aged 45-64 years at baseline (1987-89), chosen by probability sampling from four US communities. Cohort members completed three additional triennial follow-up examinations, a fifth exam in 2011-2013, a sixth exam in 2016-2017, a seventh exam in 2018-2019, an eighth exam in 2020, a ninth exam in 2021-2022, a tenth exam in 2023, and the 11th exam started in 2024. The ARIC study has been described in detail previously (2,3).

**Ethics statement:**

The ARIC study has been approved by a single Institutional Review Board (sIRB) at Johns Hopkins School of Medicine and Institutional Review Boards (IRB) at all participating institutions: University of North Carolina at Chapel Hill IRB, Johns Hopkins University School of Public Health IRB, University of Minnesota IRB, Wake Forest University Health Sciences IRB, and University of Mississippi Medical Center IRB. Study participants provided written informed consent at all study visits.

**ARIC acknowledgements:**

The Atherosclerosis Risk in Communities study has been funded in whole or in part with Federal funds from the National Heart, Lung, and Blood Institute, National Institutes of Health, Department of Health and Human Services (contract numbers 75N92022D00001,

75N92022D00002, 75N92022D00003, 75N92022D00004, 75N92022D00005). The authors thank the staff and participants of the ARIC study for their important contributions.
WGS for “NHLBI TOPMed: Atherosclerosis Risk in Communities (ARIC)” (phs001211) was performed at the Baylor College of Medicine Human Genome Sequencing Center (HHSN268201500015C and 3U54HG003273-12S2) and the Broad Institute for MIT and Harvard (3R01HL092577- 06S1). The Genome Sequencing Program (GSP) was funded by the National Human Genome Research Institute (NHGRI), the National Heart, Lung, and Blood Institute (NHLBI), and the National Eye Institute (NEI). The GSP Coordinating Center (U24 HG008956) contributed to cross program scientific initiatives and provided logistical and general study coordination. The Centers for Common Disease Genomics (CCDG) program was supported by NHGRI and NHLBI, and whole genome sequencing was performed at the Baylor College of Medicine Human Genome Sequencing Center (UM1 HG008898).

##### CARDIA

The Coronary Artery Risk Development in Young Adults study (dbGaP accession phs000285) is a prospective multicenter study with 5,115 adults Caucasian and African American participants of the age group 18-30 years at baseline, recruited from four centers at the baseline examination in 1985-1986 (4). The recruitment was done from the total community in Birmingham, AL, from selected census tracts in Chicago, IL and Minneapolis, MN; and from the Kaiser Permanente health plan membership in Oakland, CA. Nine examinations have been completed in the years 0, 2, 5, 7, 10, 15, 20, 25 and 30, with high retention rates (91%, 86%, 81%, 79%, 74%, 72%, 72%, and 71%, respectively) and written informed consent was obtained in each visit.

**Ethics statement:**

All CARDIA participants provided informed consent, and the study was approved by the Institutional Review Boards of the University of Alabama at Birmingham and the University of Texas Health Science Center at Houston.

**CARDIA acknowledgements:**

The Coronary Artery Risk Development in Young Adults Study (CARDIA) is conducted and supported by the National Heart, Lung, and Blood Institute (NHLBI) in collaboration with the University of Alabama at Birmingham (75N92023D00002 & 75N92023D00005), Northwestern University (75N92023D00004), University of Minnesota (75N92023D00006), and Kaiser Foundation Research Institute (75N92023D00003). CARDIA was also partially supported by the Intramural Research Program of the National Institute on Aging (NIA) and an intra-agency agreement between NIA and NHLBI (AG0005).

#### CFS

The Cleveland Family Study (CFS) was designed to examine the genetic basis of sleep apnea in 2,534 African-American and European-American individuals from 356 families. Index probands with confirmed sleep apnea were recruited from sleep centers in northern Ohio, supplemented with additional family members and neighborhood control families (5). Four visits occurred between 1990 and 2006; in the first 3, data were collected in participants' homes while the last occurred in a clinical research center (2000 - 2006). Measurements included sleep apnea monitoring, blood pressure, anthropometry, spirometry and other related phenotypes. Blood samples (overnight fasting, before bed and following an oral glucose tolerance test), nasal and oral ultrasound, and ECG were also obtained during the 4th exam. Institutional Review Board approval and signed informed consent was obtained for all participants.

**Ethics statement:**

Cleveland Family Study was approved by the Institutional Review Board (IRB) of Case Western Reserve University and Mass General Brigham (formerly Partners HealthCare). Written informed consent was obtained from all participants.

#### CHS

The Cardiovascular Health Study (CHS**)** is a population-based cohort study of risk factors for coronary heart disease and stroke in adults ≥65 years conducted across four field centers (6). The original predominantly European ancestry cohort of 5,201 persons was recruited in 1989-1990 from random samples of the Medicare eligibility lists; subsequently, an additional predominantly African-American cohort of 687 persons was enrolled for a total sample of 5,888. Participants underwent annual extensive clinical examinations through 1999 with additional phone contact at six month intervals between clinic visits and every six months after clinic visits ended to ascertain hospitalizations and health status. Blood samples were drawn from all participants at their baseline examination and DNA was subsequently extracted from available samples.

**Ethics statement:**

CHS was approved by institutional review committees at each field center and individuals in the present analysis had available DNA and gave informed consent including consent to use of genetic information for the study of cardiovascular disease.

**CHS acknowledgements:**

Cardiovascular Health Study: This research was supported by contracts HHSN268201200036C, HHSN268200800007C, HHSN268201800001C, N01HC55222, N01HC85079, N01HC85080, N01HC85081, N01HC85082, N01HC85083, N01HC85086, 75N92021D00006, and grants R01HL120393, U01HL080295, U01HL130114, and HL105756 from the National Heart, Lung, and Blood Institute (NHLBI), with additional contribution from the National Institute of Neurological Disorders and Stroke (NINDS). Additional support was provided by R01AG023629 from the National Institute on Aging (NIA). A full list of principal CHS investigators and institutions can be found at CHS-NHLBI.org. The content is solely the responsibility of the authors and does not necessarily represent the official views of the National Institutes of Health.

#### COPDGene

COPDGene (7) is a cohort study for respiratory disease research, recruiting more than 10,000 subjects between the ages of 45 and 80 who had at least 10 pack-years of smoking during January 2008 - June 2011 at 21 clinical centers. Participants were characterized using spirometry, six-minute walk, inspiratory and expiratory chest CT scans, respiratory symptoms, medical history, medication history and 36-Item short form health survey. In the current analysis, we only used COPDGene control participants (meaning, individuals without COPD).

**OSA phenotype in COPDGene:**

In their respiratory disease questionnaire, participants were asked: “Have you ever had sleep apnea?”, and if the response was “yes”, they were asked: “Was it diagnosed by a doctor or other health professional?”. We used OSA definition based on a positive (“yes”) response to diagonisis by a doctor or other health professional.

**Ethics statement:**

All COPDGene participants provided written informed consent, and the study was approved by the Institutional Review Boards of the participating clinical centers.

**COPDGene acknowledgements:**

The COPDGene project described was supported by Award Number U01 HL089897 and Award Number U01 HL089856 from the National Heart, Lung, and Blood Institute. The content is solely the responsibility of the authors and does not necessarily represent the official views of the National Heart, Lung, and Blood Institute or the National Institutes of Health. The COPDGene project is also supported by the COPD Foundation through contributions made to an Industry Advisory Board comprised of AstraZeneca, Boehringer Ingelheim, GlaxoSmithKline, Novartis, Pfizer, Siemens and Sunovion. A full listing of COPDGene investigators can be found at: http://www.copdgene.org/directory

#### FHS

The Framingham Heart Study (dbGaP accession phs000007) began in 1948 with the recruitment of an original cohort of 5,209 men and women (mean age 44 years; 55 percent women). In 1971 a second generation of study participants was enrolled; this cohort (mean age 37 years; 52% women) consisted of 5,124 children and spouses of children of the original cohort. A third-generation cohort of 4,095 children of offspring cohort participants (mean age 40 years; 53 percent women) was enrolled in 2002-2005 and are seen every 4 to 8 years. Details of study designs for the three cohorts are summarized elsewhere (8–10). At each clinic visit, a medical history was obtained, and participants underwent a physical examination. Only study participants consented for genetic and non-genetic data are included. FHS has been approved by the Boston University IRB.

**Ethics statement:**

The Framingham Heart Study was approved by the Institutional Review Board of the Boston University Medical Center. All study participants provided written informed consent.

**FHS acknowledgements:**

The Framingham Heart Study (FHS) acknowledges the support of contracts NO1-HC-25195, HHSN268201500001I and 75N92019D00031 from the National Heart, Lung and Blood Institute and grant supplement R01 HL092577-06S1 for this research. We also acknowledge the dedication of the FHS study participants without whom this research would not be possible.

#### HCHS/SOL

The Hispanic Community Health Study/Study of Latinos (dbGaP accession phs000810) is a community-based longitudinal cohort study of 16,415 self-identified Hispanic/Latino persons aged 18–74 years and selected from households in predefined census-block groups across four US field centers (in Chicago, Miami, the Bronx, and San Diego). The census-block groups were chosen to provide diversity among cohort participants with regard to socioeconomic status and national origin or background (11–13). The HCHS/SOL cohort includes participants who self-identified as having a Hispanic/Latino background; the largest groups are Central American (n = 1,730), Cuban (n = 2,348), Dominican (n = 1,460), Mexican (n = 6,471), Puerto Rican (n = 2,728), and South American (n = 1,068). The HCHS/SOL baseline clinical examination occurred between 2008 and 2011 and included comprehensive biological, behavioral, and sociodemographic assessments. Visit 2 took place between 2014 and 2017, which re-examined 11,623 participants from the baseline sample. Visit 3 has started in 2020 and will last 4 years, ending on January 2024. In addition to clinic visit, participants are contacted annually to assess clinical outcomes. The study was approved by the Institutional Review Boards at each participating institution and written informed consent was obtained from all participants.

**Ethics statement:**

This study was approved by the institutional review boards (IRBs) at each field center, where all participants gave written informed consent, and by the Non-Biomedical IRB at the University of North Carolina at Chapel Hill, to the HCHS/SOL Data Coordinating Center. All IRBs approving the study are: Non-Biomedical IRB at the University of North Carolina at Chapel Hill. Chapel Hill, NC; Einstein IRB at the Albert Einstein College of Medicine of Yeshiva University. Bronx, NY; IRB at Office for the Protection of Research Subjects (OPRS), University of Illinois at Chicago. Chicago, IL; Human Subject Research Office, University of Miami. Miami, FL; Institutional Review Board of San Diego State University. San Diego, CA.

**HCHS/SOL acknowledgements:**

The Hispanic Community Health Study/Study of Latinos is a collaborative study supported by contracts from the National Heart, Lung, and Blood Institute (NHLBI) to the University of North Carolina (HHSN268201300001I / N01-HC-65233), University of Miami (HHSN268201300004I / N01-HC- 65234), Albert Einstein College of Medicine (HHSN268201300002I / N01-HC-65235), University of Illinois at Chicago – HHSN268201300003I / N01- HC-65236 Northwestern Univ), and San Diego State University (HHSN268201300005I / N01-HC-65237). The following

Institutes/Centers/Offices have contributed to the HCHS/SOL through a transfer of funds to the NHLBI: National Institute on Minority Health and Health Disparities, National Institute on Deafness and Other Communication Disorders, National Institute of Dental and Craniofacial Research, National Institute of Diabetes and Digestive and Kidney Diseases, National Institute of Neurological Disorders and Stroke, NIH Institution-Office of Dietary Supplements.

#### JHS

The Jackson Heart Study (dbGaP accession phs000286) is a longitudinal investigation of genetic and environmental risk factors associated with the disproportionate burden of cardiovascular disease in African Americans (14,15). At baseline, the JHS recruited 5306 African American residents of the Jackson, Mississippi Metropolitan Statistical Area, which included approximately 6.6% of all African American adults aged 35-84 residing in the area. Participants were recruited via random sampling (17% of participants), volunteers (30%), prior participants in the Atherosclerosis Risk in Communities (ARIC) study (31%), and secondary family members (22%). Among these participants, approximately 3400 gave consent that allows genetic research. JHS participants received three back-to-back clinical examinations (Exam 1, 2000-2004; Exam 2, 2005-2008; and Exam 3, 2009-2013), and a fourth clinical examination started in 2020. Participants are also contacted annually by telephone to update personal and health information including vital status, interim medical events, hospitalizations, functional status and sociocultural information.

**Ethics statement:**

The Institutional Review Boards at Jackson State University, Tougaloo College, and the University of Mississippi Medical Center approved the study, and all participants provided written informed consent.

**JHS acknowledgements:**

The Jackson Heart Study (JHS) is supported and conducted in collaboration with Jackson State University (HHSN268201800013I), Tougaloo College (HHSN268201800014I), the Mississippi State Department of Health (HHSN268201800015I) and the University of Mississippi Medical Center (HHSN268201800010I, HHSN268201800011I and HHSN268201800012I) contracts from the National Heart, Lung, and Blood Institute (NHLBI) and the National Institute for Minority Health and Health Disparities (NIMHD). The authors also wish to thank the staffs and participants of the JHS.

Genome sequencing (dbGap accession phs000964) was performed at the Northwest Genomics Center (HHSN268201100037C). Core support including centralized genomic read mapping and genotype calling, along with variant quality metrics and filtering were provided by the TOPMed Informatics Research Center (R01HL117626; contract HHSN268201800002I). Core support including phenotype harmonization, data management, sample-identity QC, and general program coordination were provided by the TOPMed Data Coordinating Center (R01HL120393; U01HL120393; contract HHSN268201800001I).

#### MESA

The Multi-Ethnic Study of Atherosclerosis (dbGaP accession phs000209) is a study of the characteristics of subclinical cardiovascular disease (disease detected non-invasively before it has produced clinical signs and symptoms) and the risk factors that predict progression to clinically overt cardiovascular disease or progression of the subclinical disease (16). MESA consisted of a diverse, population-based sample of an initial 6,814 asymptomatic men and women aged 45-84. 38 percent of the recruited participants were White, 28 percent African American, 22 percent Hispanic, and 12 percent Asian, predominantly of Chinese descent. Participants were recruited from six field centers across the United States: Wake Forest University, Columbia University, Johns Hopkins University, University of Minnesota, Northwestern University and University of California - Los Angeles. Participants are being followed for identification and characterization of cardiovascular disease events, including acute myocardial infarction and other forms of coronary heart disease (CHD), stroke, and congestive heart failure; for cardiovascular disease interventions; and for mortality. The first examination took place over two years, from July 2000 - July 2002. It was followed by five examination periods that were 17-20 months in length. Participants have been contacted every 9 to 12 months throughout the study to assess clinical morbidity and mortality.

**Ethics statements:**

All MESA participants provided written informed consent, and the study was approved by the Institutional Review Boards at The Lundquist Institute (formerly Los Angeles BioMedical Research Institute) at Harbor-UCLA Medical Center, University of Washington, Wake Forest School of Medicine, Northwestern University, University of Minnesota, Columbia University, and Johns Hopkins University.

**MESA acknowledgements:**

The MESA projects are conducted and supported by the National Heart, Lung, and Blood Institute (NHLBI) in collaboration with MESA investigators. Support for the Multi-Ethnic Study of Atherosclerosis (MESA) projects are conducted and supported by the National Heart, Lung, and Blood Institute (NHLBI) in collaboration with MESA investigators. Support for MESA is provided by contracts 75N92020D00001, HHSN268201500003I, N01-HC-95159, 75N92020D00005, N01-HC-95160, 75N92020D00002, N01-HC-95161, 75N92020D00003, N01-HC-95162, 75N92020D00006, N01-HC-95163, 75N92020D00004, N01-HC-95164, 75N92020D00007, N01-HC-95165, N01-HC-95166, N01-HC-95167, N01-HC-95168, N01-HC-95169, UL1-TR-000040, UL1-TR-001079, UL1-TR-001420, UL1TR001881, DK063491, R01AG08598, and R01HL105756. Funding for MESA SHARe genotyping was provided by NHLBI Contract N02-HL-64278. Genotyping was performed at Affymetrix (Santa Clara, California, USA) and the Broad Institute of Harvard and MIT (Boston, Massachusetts, USA) using the Affymetrix Genome-Wide Human SNP Array 6.0. The authors thank the other investigators, the staff, and the participants of the MESA study for their valuable contributions. A full list of participating MESA investigators and institutes can be found at http://www.mesa-nhlbi.org.

##### WHI

The Women's Health Initiative (WHI) is a prospective national health study focused on identifying optimal strategies for preventing chronic diseases that are the major causes of death and disability in postmenopausal women. The WHI initially recruited 161,808 women between 1993 and 1997 with the goal of including a socio-demographically diverse population with diversity background groups proportionate to the total minority population of US women aged 50-79 years. The WHI consists of two major parts: a set of randomized Clinical Trials and an Observational Study. The WHI Clinical Trials (CT; N=68,132) includes three overlapping components, each a randomized controlled comparison: the Hormone Therapy Trials (HT), Dietary Modification Trial, and Calcium and Vitamin D Trial. A parallel prospective observational study (OS; N = 93,676) examined biomarkers and risk factors associated with various chronic diseases. While the HT trials ended in the mid-2000s, active follow-up of the WHI-CT and WHI-OS cohorts has continued for over 25 years, with the accumulation of large numbers of diverse clinical outcomes, risk factor measurements, medication use, and many other types of data.

**Ethics statement:**
All WHI participants provided informed consent and the study was approved by the Institutional Review Board (IRB) of the Fred Hutchinson Cancer Research Center.

**WHI acknowledgements:**
The WHI program is funded by the National Heart, Lung, and Blood Institute, National Institutes of Health, U.S. Department of Health and Human Services through contracts 75N92021D00001, 75N92021D00002, 75N92021D00003, 75N92021D00004, 75N92021D00005.

#### Supplementary Note 4: Whole genome sequencing and quality control in TOPMed

We used genetic data from whole genome sequencing via the TOPMed program

(freeze 8 release) (17). The TOPMed Data Coordinating Center (DCC) created a kinship matrix to estimate recent genetic relatedness and provided genetic principal components (PCs) using the PC-Relate algorithm. Detailed information on genome sequencing, allele calling, and quality control in TOPMed can be found at [https://www.nhlbiwgs.org/topmed-whole-genome-sequencing-methods-freeze-8](https://www.nhlbiwgs.org/topmed-whole-genome-sequencing-methods-freeze-8). Only variants passing TOPMed QC with a minor allele frequency (MAF) > 0.01 were included. We further identified unrelated individuals based on the kinship matrix, ensuring they were degree-3 unrelated using the pcairPartition function from the GENESIS R package version v2.16.1

#### Supplementary Note 5: All of Us phenotypes and acknowledgements

**Clinical outcomes in All of Us:**

We identified previously-published manuscript that reported use of ICD codes to identify patients with a specific disease outcomes, with the exception of hypertension, for which we used blood pressure measures from the baseline All of Us exam. Details and references are below.

To define the condition of OSA, we used the following ICD-9 and ICD-10 codes: 780.53, 327.20, G47.39, G47.3, 780.51, 327.29, 780.57, 327.23, and G47.33. Snoring (ICD-10: R06.83) was excluded from the control group (18).

Hypertension was defined according to AHA guidelines as SBP ≥ 130, DBP ≥ 80, or use of antihypertensive medication. We extracted SBP and DBP measurements from physical exam data for all participants. The earliest measurements were used. Antihypertensive medication usage was obtained from either electronic health records (EHR) or physical exam data, using the closest date to the BP measurement. Antihypertensive medications included peripheral vasodilators, agents acting on the renin-angiotensin system, beta-blocking agents, antihypertensives, calcium channel blockers, and diuretics. SBP and DBP values were increased by 15 mmHg and 10 mmHg, respectively (19).

For stroke, we used the following codes: hemorrhagic stroke (ICD-9: 431; ICD-10: I61.XX), ischemic stroke (ICD-9: 433.XX-434.XX, 436; ICD-10: I63.XXX), and transient ischemic attack (TIA) (ICD-9: 435.X; ICD-10: G45.9) (20).

Type 2 diabetes was identified using the ICD-9 code E11.X (21).

Atrial fibrillation was identified using ICD-9 code 427.3 and ICD-10 code I48.X (21).

For asthma, we used the following ICD-10 codes: J45.2, J45.3, J45.4, J45.5, J45.9, J45.990, J45.991, and J45.998 (22).

For chronic obstructive pulmonary disease (COPD), we used ICD-9 codes 491.X, 492.X, 496, and ICD-10 codes J41.X, J42.X, J43.X, and J44.X (23).

Chronic kidney disease (CKD) was identified using the following codes: chronic renal failure (ICD-9: 403.11, 404.12, 404.13, 404.92, 585, 585.1, 585.2, 585.3, 585.4, 585.5, 585.6, 585.9, 586, 587), systemic disease causing CKD (ICD-9: 203.0, 277.3, 287.0, 446.0, 446.2, 446.4, 446.6, 710.0, 710.1), and ICD-10 codes for CKD (N18.1, N18.2, N18.3, N18.4, N18.5, N18.6, N18.9) (24).

For coronary artery disease (CAD), we used ICD-9 codes 410.X, 411.0, 412.X, 429.79, and ICD-10 codes I21.X, I22.X, I23.X, I24.1, I25.2 (21).

Heart failure was identified using ICD-10 codes I50.2, I50.3, and ICD-9 code 428.X (25,26).

**All of Us acknowledgements:**

The All of Us Research Program is supported by the National Institutes of Health, Office of the Director: Regional Medical Centers: 1 OT2 OD026549; 1 OT2 OD026554; 1 OT2 OD026557; 1 OT2 OD026556; 1 OT2 OD026550; 1 OT2 OD 026552; 1 OT2 OD026553; 1 OT2 OD026548; 1 OT2 OD026551; 1 OT2 OD026555; IAA #: AOD 16037; Federally Quali- fied Health Centers: HHSN 263201600085U; Data and Research Center: 5 U2C OD023196; Biobank: 1 U24 OD023121; The Participant Center: U24 OD023176; Participant Technology Systems Center: 1 U24 OD023163; Communications and Engagement: 3 OT2 OD023205; 3 OT2 OD023206; and Community Partners: 1 OT2 OD025277; 3 OT2 OD025315; 1 OT2 OD025337; 1 OT2 OD025276. The All of Us Research Program would not be possible without the partnership of its participants.

#### Supplementary Note 6: FinnGen full ethics statement and acknowledgement

**Ethics statement:**

Study subjects in FinnGen provided informed consent for biobank research, based on the Finnish Biobank Act. Alternatively, separate research cohorts, collected prior the Finnish Biobank Act came into effect (in September 2013) and start of FinnGen (August 2017), were collected based on study-specific consents and later transferred to the Finnish biobanks after approval by Fimea (Finnish Medicines Agency), the National Supervisory Authority for Welfare and Health. Recruitment protocols followed the biobank protocols approved by Fimea. The

Coordinating Ethics Committee of the Hospital District of Helsinki and Uusimaa (HUS) statement number for the FinnGen study is Nr HUS/990/2017.

The FinnGen study Is approved by Finnish Institute for Health and Welfare (permit numbers: THL/2031/6.02.00/2017, THL/1101/5.05.00/2017, THL/341/6.02.00/2018, THL/2222/6.02.00/2018, THL/283/6.02.00/2019, THL/1721/5.05.00/2019 and THL/1524/5.05.00/2020), Digital and population data service agency (permit numbers: VRK43431/2017-3, VRK/6909/2018-3, VRK/4415/2019-3), the Social Insurance Institution (permit numbers: KELA 58/522/2017, KELA 131/522/2018, KELA 70/522/2019, KELA 98/522/2019, KELA 134/522/2019, KELA 138/522/2019, KELA 2/522/2020, KELA 16/522/2020), Findata permit numbers THL/2364/14.02/2020, THL/4055/14.06.00/2020, THL/3433/14.06.00/2020, THL/4432/14.06/2020, THL/5189/14.06/2020, THL/5894/14.06.00/2020, THL/6619/14.06.00/2020, THL/209/14.06.00/2021, THL/688/14.06.00/2021, THL/1284/14.06.00/2021, THL/1965/14.06.00/2021, THL/5546/14.02.00/2020, THL/2658/14.06.00/2021, THL/4235/14.06.00/2021, Statistics Finland (permit numbers: TK-53-1041-17 and TK/143/07.03.00/2020 (earlier TK-53-90-20) TK/1735/07.03.00/2021, TK/3112/07.03.00/2021) and Finnish Registry for Kidney Diseases permission/extract from the meeting minutes on $^{4t}$h July 2019.

The Biobank Access Decisions for FinnGen samples and data utilized in FinnGen Data Freeze 10 include: THL Biobank BB2017_55, BB2017_111, BB2018_19, BB_2018_34, BB_2018_67, BB2018_71, BB2019_7, BB2019_8, BB2019_26, BB2020_1, BB2021_65, Finnish Red Cross Blood Service Biobank 7.12.2017, Helsinki Biobank HUS/359/2017, HUS/248/2020, HUS/430/2021 §28, §29, HUS/150/2022 §12, §13, §14, §15, §16, §17, §18, §23, §58 and §59, Auria Biobank AB17-5154 and amendment #1 (August 17 2020) and amendments BB_2021-0140, BB_2021-0156 (August 26 2021, Feb 2 2022), BB_2021-0169, BB_2021-0179, BB_2021-0161, AB20-5926 and amendment #1 (April 23 2020) and it´s modification (Sep 22 2021), BB_2022-0262, BB_2022-0256, Biobank Borealis of Northern Finland_2017_1013, 2021_5010, 2021_5018, 2021_5015, 2021_5015 Amendment, 2021_5023, 2021_5023 Amendment, 2021_5017,

2022_6001, 2022_6006 Amendment, BB22-0067, 2022_0262, Biobank of Eastern Finland 1186/2018 and amendment 22§/2020, 53§/2021, 13§/2022, 14§/2022, 15§/2022, 27§/2022, 28§/2022, 29§/2022, 33§/2022, 35§/2022, 36§/2022, 37§/2022, 39§/2022, 7§/2023, Finnish Clinical Biobank Tampere MH0004 and amendments (21.02.2020 & 06.10.2020), 8§/2021, 9§/2021, §9/2022, §10/2022, §12/2022, 13§/2022, §20/2022, §21/2022, §22/2022, §23/2022, 28§/2022, 29§/2022, 30§/2022, 31§/2022, 32§/2022, 38§/2022, 40§/2022, 42§/2022, 1§/2023, Central Finland Biobank 1-2017, BB_2021-0161, BB_2021-0169, BB_2021-0179, BB_2021-0170, BB_2022-0256, and Terveystalo Biobank STB 2018001 and amendment 2$^{5t}$h Aug 2020, Finnish Hematological Registry and Clinical Biobank decision 1$^{8t}$h June 2021, Arctic biobank P0844: ARC_2021_1001.

**FinnGen acknowledgements:**

We want to acknowledge the participants and investigators of FinnGen study. The FinnGen project is funded by two grants from Business Finland (HUS 4685/31/2016 and UH 4386/31/2016) and the following industry partners: AbbVie Inc., AstraZeneca UK Ltd, Biogen MA Inc., Bristol Myers Squibb (and Celgene Corporation & Celgene International II Sàrl), Genentech Inc., Merck Sharp & Dohme LCC, Pfizer Inc., GlaxoSmithKline Intellectual Property Development Ltd., Sanofi US Services Inc., Maze Therapeutics Inc., Janssen Biotech Inc, Novartis Pharma AG, and Boehringer Ingelheim International GmbH. Following biobanks are acknowledged for delivering biobank samples to FinnGen: Auria Biobank (www.auria.fi/biopankki), THL Biobank (www.thl.fi/biobank), Helsinki Biobank (www.helsinginbiopankki.fi), Biobank Borealis of Northern Finland (https://www.ppshp.fi/Tutkimus-ja-opetus/Biopankki/Pages/Biobank-Borealis-briefly-in-English.aspx), Finnish Clinical Biobank Tampere (www.tays.fi/en-

US/Research_and_development/Finnish_Clinical_Biobank_Tampere), Biobank of Eastern Finland (www.ita-suomenbiopankki.fi/en), Central Finland Biobank (www.ksshp.fi/fi-FI/Potilaalle/Biopankki), Finnish Red Cross Blood Service Biobank (www.veripalvelu.fi/verenluovutus/biopankkitoiminta), Terveystalo Biobank (www.terveystalo.com/fi/Yritystietoa/Terveystalo-Biopankki/Biopankki/) and Arctic Biobank (https://www.oulu.fi/en/university/faculties-and-units/faculty-medicine/northern-finland-birth-cohorts-and-arctic-biobank). All Finnish Biobanks are members of BBMRI.fi infrastructure (www.bbmri.fi). Finnish Biobank Cooperative -FINBB (https://finbb.fi/) is the coordinator of BBMRI-ERIC operations in Finland. The Finnish biobank data can be accessed through the Fingenious® services (https://site.fingenious.fi/en/) managed by FINBB.

#### Supplementary Note 7: TOPMed and CCDG acknowledgments

Molecular data for the Trans-Omics in Precision Medicine (TOPMed) program was supported by the National Heart, Lung and Blood Institute (NHLBI). Genome sequencing for "NHLBI TOPMed: Genome sequencing for "NHLBI TOPMed: Whole Genome Sequencing and Related Phenotypes in the Framingham Heart Study" (phs000974.v4.p3) was performed at the Broad Institute Genomics Platform (3R01HL092577-06S1, 3U54HG003067-12S2). Genome sequencing for "NHLBI TOPMed: The Jackson Heart Study" (phs000964.v1.p1) was performed at the Northwest Genomics Center (HHSN268201100037C). Genome sequencing for the "NHLBI TOPMed: The Atherosclerosis Risk in Communities Study" (phs001211.v3.p2) was performed at the Broad Institute Genomics Platform (3R01HL092577-06S1) and the Baylor College of Medicine Human

Genome Sequencing Center (HHSN268201500015C, 3U54HG003273-12S2). Genome sequencing for “NHLBI TOPMed: Cleveland Family Study” (phs000954.v3.p2) was performed at the Northwest Genomics Center (3R01HL098433-05S1, HHSN268201600032I). Genome sequencing for “NHLBI TOPMed: Genetic Epidemiology of COPD (COPDGene) in the TOPMed Program” (phs000951) was performed at the University of Washington Northwest Genomics Center (3R01 HL089856-08S1) and the Broad Institute of MIT and Harvard (HHSN268201500014C). Genomics sequencing for “NHLBI TOPMed: Cardiovascular Health Study” (phs001368.v2.p1) was performed at the Baylor College of Medicine Human Genome Sequencing Center (3U54HG003273-12S2, HHSN268201500015C, HHSN268201600033I). Genome sequencing for “NHLBI TOPMed: Hispanic Community Health Study/Study of Latinos” (phs001395.v1.p1) was performed at the Baylor College of Medicine Human Genome Sequencing Center (HHSN268201600033I). Genome sequencing for “NHLBI TOPMed: Women’s Health Initiative (WHI)” (phs001237.v2.p1) was performed at the Broad Institute of MIT and Harvard (HHSN268201500014C). Genome sequencing for “NHLBI TOPMed: Multi-Ethnic Study of Atherosclerosis” (phs001416.v2.p1) was performed at Broad Institute Genomics Platform (HHSN268201500014C, 3U54HG003067-13S1. Core support including centralized genomic read mapping and genotype calling, along with variant quality metrics and filtering were provided by the TOPMed Informatics Research Center (3R01HL-117626-02S1; contract HHSN268201800002I). Core support including phenotype harmonization, data management, sample-identity QC, and general program coordination were provided by the TOPMed Data Coordinating Center (R01HL- 120393; U01HL-120393; contract HHSN268201800001I). We gratefully acknowledge the studies and participants who provided biological samples and data for TOPMed. The Genome Sequencing Program (GSP) was funded by the National Human Genome Research Institute (NHGRI), the National Heart, Lung, and Blood Institute (NHLBI), and the National Eye Institute (NEI). The GSP Coordinating Center (U24 HG008956) contributed to cross-program scientific initiatives and provided logistical and general study coordination. The Centers for Common Disease Genomics (CCDG) program was supported by NHGRI and NHLBI, and whole genome sequencing was performed at the Baylor College of Medicine Human Genome Sequencing Center (UM1 HG008898 and R01HL059367).

#### Supplementary Note 8: TOPMed consortium members

Namiko Abe[1], Gonçalo Abecasis[2], Francois Aguet[3], Christine Albert[4], Laura Almasy[5], Alvaro Alonso[6], Seth Ament[7], Peter Anderson[8], Pramod Anugu[9], Deborah Applebaum-Bowden[10], Kristin Ardlie[3], Dan Arking[11], Donna K Arnett[12], Allison Ashley-Koch[13], Stella Aslibekyan[14], Tim Assimes[15], Paul Auer[16], Dimitrios Avramopoulos[11], Najib Ayas[17], Adithya Balasubramanian[18], John Barnard[19], Kathleen Barnes[20], R. Graham Barr[21], Emily Barron-Casella[11], Lucas Barwick[22], Terri Beaty[11], Gerald Beck[23], Diane Becker[24], Lewis Becker[11], Rebecca Beer[25], Amber Beitelshees[7], Emelia Benjamin[26], Takis Benos[27], Marcos Bezerra[28], Larry Bielak[2], Joshua Bis[29], Thomas Blackwell[2], John Blangero[30], Eric Boerwinkle[31], Donald W. Bowden[32], Russell Bowler[33], Jennifer Brody[8], Ulrich Broeckel[34], Jai Broome[8], Deborah Brown[35], Karen Bunting[1], Esteban Burchard[36], Carlos Bustamante[37], Erin Buth[38], Brian Cade[39], Jonathan Cardwell[40], Vincent Carey[41], Julie Carrier[42], April Carson[43], Cara Carty[44], Richard Casaburi[45], Juan P Casas Romero[46], James Casella[11], Peter Castaldi[47], Mark Chaffin[3], Christy Chang[7], Yi-Cheng Chang[48], Daniel

Chasman[49], Sameer Chavan[40], Bo-Juen Chen[1], Wei-Min Chen[50], Yii-Der Ida Chen[51], Michael Cho[41], Seung Hoan Choi[3], Lee-Ming Chuang[52], Mina Chung[53], Ren-Hua Chung[54], Clary Clish[55], Suzy Comhair[56], Matthew Conomos[38], Elaine Cornell[57], Adolfo Correa[58], Carolyn Crandall[45], James Crapo[59], L. Adrienne Cupples[60], Joanne Curran[61], Jeffrey Curtis[62], Brian Custer[63], Coleen Damcott[7], Dawood Darbar[64], Sean David[65], Colleen Davis[8], Michelle Daya[40], Mariza de Andrade[66], Lisa de las Fuentes[67], Paul de Vries[68], Michael DeBaun[69], Ranjan Deka[70], Dawn DeMeo[41], Scott Devine[7], Huyen Dinh[18], Harsha Doddapaneni[18], Qing Duan[71], Shannon Dugan-Perez[18], Ravi Duggirala[72], Jon Peter Durda[57], Susan K. Dutcher[73], Charles Eaton[74], Lynette Ekunwe[9], Adel El Boueiz[75], Patrick Ellinor[76], Leslie Emery[8], Serpil Erzurum[19], Charles Farber[50], Jesse Farek[18], Tasha Fingerlin[77], Matthew Flickinger[2], Myriam Fornage[31], Nora Franceschini[78], Chris Frazar[8], Mao Fu[7], Stephanie M. Fullerton[8], Lucinda Fulton[79], Stacey Gabriel[3], Weiniu Gan[25], Shanshan Gao[40], Yan Gao[9], Margery Gass[80], Heather Geiger[81], Bruce Gelb[82], Mark Geraci[27], Soren Germer[1], Robert Gerszten[83], Auyon Ghosh[41], Richard Gibbs[18], Chris Gignoux[15], Mark Gladwin[27], David Glahn[84], Stephanie Gogarten[8], Da-Wei Gong[7], Harald Goring[85], Sharon Graw[86], Kathryn J. Gray[87], Daniel Grine[40], Colin Gross[2], C. Charles Gu[79], Yue Guan[7], Xiuqing Guo[51], Namrata Gupta[3], David M. Haas[88], Jeff Haessler[80], Michael Hall[89], Yi Han[18], Patrick Hanly[90], Daniel Harris[91], Nicola L. Hawley[92], Jiang He[93], Ben Heavner[38], Susan Heckbert[94], Ryan Hernandez[36], David Herrington[95], Craig Hersh[96], Bertha Hidalgo[14], James Hixson[31], Brian Hobbs[41], John Hokanson[40], Elliott Hong[7], Karin Hoth[97], Chao (Agnes) Hsiung[98], Jianhong Hu[18], Yi-Jen Hung[99], Haley Huston[100], Chii Min Hwu[101], Marguerite Ryan Irvin[14], Rebecca Jackson[102], Deepti Jain[8], Cashell Jaquish[103], Jill Johnsen[104], Andrew Johnson[25], Craig Johnson[8], Rich Johnston[6], Kimberly Jones[11], Hyun Min Kang[105], Robert Kaplan[106], Sharon Kardia[2], Shannon

Kelly[36], Eimear Kenny[82], Michael Kessler[7], Alyna Khan[8], Ziad Khan[18], Wonji Kim[107], John Kimoff[108], Greg Kinney[109], Barbara Konkle[110], Charles Kooperberg[80], Holly Kramer[111], Christoph Lange[112], Ethan Lange[40], Leslie Lange[113], Cathy Laurie[8], Cecelia Laurie[8], Meryl LeBoff[41], Jiwon Lee[41], Sandra Lee[18], Wen-Jane Lee[101], Jonathon LeFaive[2], David Levine[8], Dan Levy[25], Joshua Lewis[7], Xiaohui Li[51], Yun Li[71], Henry Lin[51], Honghuang Lin[114], Xihong Lin[115], Simin Liu[116], Yongmei Liu[117], Yu Liu[118], Ruth J.F. Loos[119], Steven Lubitz[76], Kathryn Lunetta[114], James Luo[25], Ulysses Magalang[120], Michael Mahaney[61], Barry Make[11], Ani Manichaikul[50], Alisa Manning[121], JoAnn Manson[41], Lisa Martin[122], Melissa Marton[81], Susan Mathai[40], Rasika Mathias[11], Susanne May[38], Patrick McArdle[7], Merry-Lynn McDonald[123], Sean McFarland[107], Stephen McGarvey[124], Daniel McGoldrick[125], Caitlin McHugh[38], Becky McNeil[126], Hao Mei[9], James Meigs[127], Vipin Menon[18], Luisa Mestroni[86], Ginger Metcalf[18], Deborah A Meyers[128], Emmanuel Mignot[129], Julie Mikulla[25], Nancy Min[9], Mollie Minear[130], Ryan L Minster[27], Braxton D. Mitchell[7], Matt Moll[47], Zeineen Momin[18], May E. Montasser[7], Courtney Montgomery[131], Donna Muzny[18], Josyf C Mychaleckyj[50], Girish Nadkarni[82], Rakhi Naik[11], Take Naseri[132], Pradeep Natarajan[3], Sergei Nekhai[133], Sarah C. Nelson[38], Bonnie Neltner[40], Caitlin Nessner[18], Deborah Nickerson[134], Osuji Nkechinyere[18], Kari North[71], Jeff O'Connell[135], Tim O'Connor[7], Heather Ochs-Balcom[136], Geoffrey Okwuonu[18], Allan Pack[137], David T. Paik[138], Nicholette Palmer[139], James Pankow[140], George Papanicolaou[25], Cora Parker[141], Gina Peloso[142], Juan Manuel Peralta[72], Marco Perez[15], James Perry[7], Ulrike Peters[143], Patricia Peyser[2], Lawrence S Phillips[6], Jacob Pleiness[2], Toni Pollin[7], Wendy Post[144], Julia Powers Becker[145], Meher Preethi Boorgula[40], Michael Preuss[82], Bruce Psaty[8], Pankaj Qasba[25], Dandi Qiao[41], Zhaohui Qin[6], Nicholas Rafaels[146], Laura Raffield[147], Mahitha Rajendran[18], Vasan S. Ramachandran[114], D.C. Rao[79], Laura Rasmussen-Torvik[148], Aakrosh Ratan[50], Susan Redline[47],

Robert Reed[7], Catherine Reeves[149], Elizabeth Regan[59], Alex Reiner[150], Muagututi'a Sefuiva Reupena[151], Ken Rice[8], Stephen Rich[50], Rebecca Robillard[152], Nicolas Robine[81], Dan Roden[153], Carolina Roselli[3], Jerome Rotter[154], Ingo Ruczinski[11], Alexi Runnels[81], Pamela Russell[40], Sarah Ruuska[100], Kathleen Ryan[7], Ester Cerdeira Sabino[155], Danish Saleheen[21], Shabnam Salimi[156], Sejal Salvi[18], Steven Salzberg[11], Kevin Sandow[157], Vijay G. Sankaran[158], Jireh Santibanez[18], Karen Schwander[79], David Schwartz[40], Frank Sciurba[27], Christine Seidman[159], Jonathan Seidman[160], Frédéric Sériès[161], Vivien Sheehan[162], Stephanie L. Sherman[163], Amol Shetty[7], Aniket Shetty[40], Wayne Hui- Heng Sheu[101], M. Benjamin Shoemaker[164], Brian Silver[165], Edwin Silverman[41], Robert Skomro[166], Albert Vernon Smith[167], Jennifer Smith[2], Josh Smith[8], Nicholas Smith[94], Tanja Smith[1], Sylvia Smoller[106], Beverly Snively[168], Michael Snyder[15], Tamar Sofer[41], Nona Sotoodehnia[8], Adrienne M. Stilp[8], Garrett Storm[169], Elizabeth Streeten[7], Jessica Lasky Su[170], Yun Ju Sung[79], Jody Sylvia[41], Adam Szpiro[8], Daniel Taliun[2], Hua Tang[171], Margaret Taub[11], Kent D. Taylor[172], Matthew Taylor[86], Simeon Taylor[7], Marilyn Telen[13], Timothy A. Thornton[8], Machiko Threlkeld[173], Lesley Tinker[174], David Tirschwell[8], Sarah Tishkoff[175], Hemant Tiwari[176], Catherine Tong[177], Russell Tracy[178], Michael Tsai[140], Dhananjay Vaidya[11], David Van Den Berg[179], Peter VandeHaar[2], Scott Vrieze[140], Tarik Walker[40], Robert Wallace[97], Avram Walts[40], Fei Fei Wang[8], Heming Wang[180], Jiongming Wang[167], Karol Watson[45], Jennifer Watt[18], Daniel E. Weeks[27], Joshua Weinstock[105], Bruce Weir[8], Scott T Weiss[181], Lu-Chen Weng[76], Jennifer Wessel[182], Cristen Willer[62], Kayleen Williams[38], L. Keoki Williams[183], Carla Wilson[41], James Wilson[184], Lara Winterkorn[81], Quenna Wong[8], Joseph Wu[138], Huichun Xu[7], Lisa Yanek[11], Ivana Yang[40], Ketian Yu[2], Seyedeh Maryam Zekavat[3], Yingze Zhang[185], Snow Xueyan Zhao[59], Wei Zhao[186], Xiaofeng Zhu[187], Michael Zody[1], Sebastian Zoellner[2]

1 - New York Genome Center, New York, New York; 2 - University of Michigan, Ann Arbor, Michigan; 3 - Broad Institute, Cambridge, Massachusetts; 4 - Cedars Sinai, Boston, Massachusetts; 5 - Children's Hospital of Philadelphia, University of Pennsylvania, Philadelphia, Pennsylvania; 6 - Emory University, Atlanta, Georgia; 7 - University of Maryland, Baltimore, Maryland; 8 - University of Washington, Seattle, Washington; 9 - University of Mississippi, Jackson, Mississippi; 10 - National Institutes of Health, Bethesda, Maryland; 11 - Johns Hopkins University, Baltimore, Maryland; 12 - University of Kentucky, Lexington, Kentucky; 13 - Duke University, Durham, North Carolina; 14 - University of Alabama, Birmingham, Alabama; 15 - Stanford University, Stanford, California; 16 - Medical College of Wisconsin, Milwaukee, Wisconsin; 17 - Medicine, Providence Health Care, Vancouver; 18 - Baylor College of Medicine Human Genome Sequencing Center, Houston, Texas; 19 - Cleveland Clinic, Cleveland, Ohio; 20 - Tempus, University of Colorado Anschutz Medical Campus, Aurora, Colorado; 21 - Columbia University, New York, New York; 22 - LTRC, The Emmes Corporation, Rockville, Maryland; 23 - Quantitative Health Sciences, Cleveland Clinic, Cleveland, Ohio; 24 - Medicine, Johns Hopkins University, Baltimore, Maryland; 25 - National Heart, Lung, and Blood Institute, National Institutes of Health, Bethesda, Maryland; 26 - Boston University School of Medicine, Boston University, Massachusetts General Hospital, Boston, Massachusetts; 27 - University of Pittsburgh, Pittsburgh, Pennsylvania; 28 - Fundação de Hematologia e Hemoterapia de Pernambuco - Hemope, Recife; 29 - Cardiovascular Health Research Unit, Department of Medicine, University of Washington, Seattle, Washington; 30 - Human Genetics, University of Texas Rio Grande Valley School of Medicine, Brownsville, Texas; 31 - University of Texas Health at Houston, Houston, Texas; 32 - Department of Biochemistry, Wake Forest Baptist Health, Winston-Salem, North Carolina; 33 - National Jewish Health, National Jewish Health, Denver, Colorado; 34 - Pediatrics, Medical College of Wisconsin, Milwaukee, Wisconsin; 35 - Pediatrics, University of Texas Health at Houston, Houston, Texas; 36 - University of California, San Francisco, San Francisco, California; 37 – Biomedical Data Science, Stanford University, Stanford, California; 38 - Biostatistics, University of Washington, Seattle, Washington; 39 - Brigham and Women's Hospital, Brigham & Women's Hospital, Boston, Massachusetts; 40 -

University of Colorado at Denver, Denver, Colorado; 41 - Brigham & Women's Hospital, Boston, Massachusetts; 42 - University of Montreal; 43 - Medicine, University of Mississippi, Jackson, Mississippi; 44 - Washington State University, Pullman, Washington; 45 - University of California, Los Angeles, Los Angeles, California; 46 - Brigham & Women's Hospital; 47 - Medicine, Brigham & Women's Hospital, Boston, Massachusetts; 48 - National Taiwan University, Taipei; 49 - Division of Preventive Medicine, Brigham & Women's Hospital, Boston, Massachusetts; 50 - University of Virginia, Charlottesville, Virginia; 51 - Lundquist Institute, Torrance, California; 52 - National Taiwan University Hospital, National Taiwan University, Taipei; 53 - Cleveland Clinic, Cleveland Clinic, Cleveland, Ohio; 54 - National Health Research Institute Taiwan, Miaoli County; 55 - Metabolomics Platform, Broad Institute, Cambridge, Massachusetts; 56 - Immunity and Immunology, Cleveland Clinic, Cleveland, Ohio; 57 - University of Vermont, Burlington, Vermont; 58 - Population Health Science, University of Mississippi, Jackson, Mississippi; 59 - National Jewish Health, Denver, Colorado; 60 - Biostatistics, Boston University, Boston, Massachusetts; 61 - University of Texas Rio Grande Valley School of Medicine, Brownsville, Texas; 62 - Internal Medicine, University of Michigan, Ann Arbor, Michigan; 63 - Vitalant Research Institute, San Francisco, California; 64 - University of Illinois at Chicago, Chicago, Illinois; 65 - University of Chicago, Chicago, Illinois; 66 - Health Quantitative Sciences Research, Mayo Clinic, Rochester, Minnesota; 67 - Department of Medicine, Cardiovascular Division, Washington University in St Louis, St. Louis, Missouri; 68 - Human Genetics Center, Department of Epidemiology, Human Genetics, and Environmental Sciences, University of Texas Health at Houston, Houston, Texas; 69 - Vanderbilt University, Nashville, Tennessee; 70 - University of Cincinnati, Cincinnati, Ohio; 71 - University of North Carolina, Chapel Hill, North Carolina; 72 - University of Texas Rio Grande Valley School of Medicine, Edinburg, Texas; 73 - Genetics, Washington University in St Louis, St Louis, Missouri; 74 - Brown University, Providence, Rhode Island; 75 - Channing Division of Network Medicine, Harvard University, Cambridge, Massachusetts; 76 - Massachusetts General Hospital, Boston, Massachusetts; 77 - Center for Genes, Environment and Health, National Jewish Health, Denver, Colorado; 78 - Epidemiology, University of North Carolina, Chapel Hill, North Carolina; 79 - Washington University in St Louis, St Louis, Missouri; 80 - Fred Hutchinson Cancer Research

Center, Seattle, Washington; 81 - New York Genome Center, New York City, New York; 82 - Icahn School of Medicine at Mount Sinai, New York, New York; 83 - Beth Israel Deaconess Medical Center, Boston, Massachusetts; 84 - Department of Psychiatry, Boston Children's Hospital, Harvard Medical School, Boston, Massachusetts; 85 - University of Texas Rio Grande Valley School of Medicine, San Antonio, Texas; 86 - University of Colorado Anschutz Medical Campus, Aurora, Colorado; 87 - Obstetrics and Gynecology, Mass General Brigham, Boston, Massachusetts; 88 - OB/GYN, Indiana University, Indianapolis, Indiana; 89 - Cardiology, University of Mississippi, Jackson, Mississippi; 90 - Medicine, University of Calgary, Calgary; 91 - Genetics, University of Maryland, Philadelphia, Pennsylvania; 92 - Department of Chronic Disease Epidemiology, Yale University, New Haven, Connecticut; 93 - Tulane University, New Orleans, Louisiana; 94 - Epidemiology, University of Washington, Seattle, Washington; 95 - Wake Forest Baptist Health, Winston-Salem, North Carolina; 96 - Channing Division of Network Medicine, Brigham & Women's Hospital, Boston, Massachusetts; 97 - University of Iowa, Iowa City, Iowa; 98 - Institute of Population Health Sciences, NHRI, National Health Research Institute Taiwan, Miaoli County; 99 - Tri-Service General Hospital National Defense Medical Center; 100 - Blood Works Northwest, Seattle, Washington; 101 - Taichung Veterans General Hospital Taiwan, Taichung City; 102 - Internal Medicine, Division of Endocrinology, Diabetes and Metabolism, Oklahoma State University Medical Center, Columbus, Ohio; 103 - NHLBI, National Heart, Lung, and Blood Institute, National Institutes of Health, Bethesda, Maryland; 104 - Research Institute, Blood Works Northwest, Seattle, Washington; 105 - Biostatistics, University of Michigan, Ann Arbor, Michigan; 106 - Albert Einstein College of Medicine, New York, New York; 107 - Harvard University, Cambridge, Massachusetts; 108 - McGill University, Montreal; 109 - Epidemiology, University of Colorado at Denver, Aurora, Colorado; 110 - Medicine, Blood Works Northwest, Seattle, Washington; 111 - Public Health Sciences, Loyola University, Maywood, Illinois; 112 - Biostats, Harvard School of Public Health, Boston, Massachusetts; 113 - Medicine, University of Colorado at Denver, Aurora, Colorado; 114 - Boston University, Boston, Massachusetts; 115 - Harvard School of Public Health, Boston, Massachusetts; 116 - Epidemiology and Medicine, Brown University, Providence, Rhode Island; 117 - Cardiology, Duke University, Durham, North Carolina; 118 - Cardiovascular Institute, Stanford University,

Stanford, California; 119 - The Charles Bronfman Institute for Personalized Medicine, Icahn School of Medicine at Mount Sinai, New York, New York; 120 - Division of Pulmonary, Critical Care and Sleep Medicine, Ohio State University, Columbus, Ohio; 121 - Broad Institute, Harvard University, Massachusetts General Hospital; 122 - cardiology, George Washington University, Washington, District of Columbia; 123 - University of Alabama at Birmingham, University of Alabama, Birmingham, Alabama; 124 - Epidemiology, Brown University, Providence, Rhode Island; 125 - Genome Sciences, University of Washington, Seattle, Washington; 126 - RTI International; 127 - Medicine, Massachusetts General Hospital, Boston, Massachusetts; 128 - University of Arizona, Tucson, Arizona; 129 - Center For Sleep Sciences and Medicine, Stanford University, Palo Alto, California; 130 - National Institute of Child Health and Human Development, National Institutes of Health, Bethesda, Maryland; 131 - Genes and Human Disease, Oklahoma Medical Research Foundation, Oklahoma City, Oklahoma; 132 - Ministry of Health, Government of Samoa, Apia; 133 - Howard University, Washington, District of Columbia; 134 - Department of Genome Sciences, University of Washington, Seattle, Washington; 135 - University of Maryland, Baltimore, Maryland; 136 - University at Buffalo, Buffalo, New York; 137 - Division of Sleep Medicine/Department of Medicine, University of Pennsylvania, Philadelphia, Pennsylvania; 138 - Stanford Cardiovascular Institute, Stanford University, Stanford, California; 139 - Biochemistry, Wake Forest Baptist Health, Winston-Salem, North Carolina; 140 - University of Minnesota, Minneapolis, Minnesota; 141 - Biostatistics and Epidemiology Division, RTI International, Research Triangle Park, North Carolina; 142 - Department of Biostatistics, Boston University, Boston, Massachusetts; 143 - Fred Hutch and UW, Fred Hutchinson Cancer Research Center, Seattle, Washington; 144 - Cardiology/Medicine, Johns Hopkins University, Baltimore, Maryland; 145 - Medicine, University of Colorado at Denver, Denver, Colorado; 146 - CCPM, University of Colorado at Denver, Denver, Colorado; 147 - Genetics, University of North Carolina, Chapel Hill, North Carolina; 148 - Northwestern University, Chicago, Illinois; 149 - New York Genome Center, New York Genome Center, New York City, New York; 150 - Fred Hutchinson Cancer Research Center, University of Washington, Seattle, Washington; 151 - Lutia I Puava Ae Mapu I Fagalele, Apia; 152 - Sleep Research Unit, University of Ottawa Institute for Mental Health Research, University of Ottawa, Ottawa; 153 - Medicine, Pharmacology, Biomedical Informatics, Vanderbilt University, Nashville, Tennessee; 154 - Pediatrics, Lundquist Institute, Torrance, California; 155 - Faculdade de Medicina, Universidade de Sao Paulo, Sao Paulo; 156 - Pathology, University of Maryland, Seattle, Washington; 157 - TGPS, Lundquist Institute, Torrance, California; 158 - Division of Hematology/Oncology, Harvard University, Boston, Massachusetts; 159 - Genetics, Harvard Medical School, Boston, Massachusetts; 160 - Harvard Medical School, Boston, Massachusetts; 161 - Université Laval, Quebec City; 162 - Pediatrics, Emory University, Atlanta, Georgia; 163 - Human Genetics, Emory University, Atlanta, Georgia; 164 - Medicine/Cardiology, Vanderbilt University, Nashville, Tennessee; 165 - UMass Memorial Medical Center, Worcester, Massachusetts; 166 - University of Saskatchewan, Saskatoon; 167 - University of Michigan; 168 - Biostatistical Sciences, Wake Forest Baptist Health, Winston-Salem, North Carolina; 169 - Genomic Cardiology, University of Colorado at Denver, Aurora, Colorado; 170 - Channing Department of Medicine, Brigham & Women's Hospital, Boston, Massachusetts; 171 - Genetics, Stanford University, Stanford, California; 172 - Institute for Translational Genomics and Populations Sciences, Lundquist Institute, Torrance, California; 173 - University of Washington, Department of Genome Sciences, University of Washington, Seattle, Washington; 174 - Cancer Prevention Division of Public Health Sciences, Fred Hutchinson Cancer Research Center, Seattle, Washington; 175 - Genetics, University of Pennsylvania, Philadelphia, Pennsylvania; 176 - Biostatistics, University of Alabama, Birmingham, Alabama; 177 - Department of Biostatistics, University of Washington, Seattle, Washington; 178 - Pathology & Laboratory Medicine, University of Vermont, Burlington, Vermont; 179 - USC Methylation Characterization Center, University of Southern California, University of Southern California, California; 180 - Brigham & Women's Hospital, Mass General Brigham, Boston, Massachusetts; 181 - Channing Division of Network Medicine, Department of Medicine, Brigham & Women's Hospital, Boston, Massachusetts; 182 - Epidemiology, Indiana University, Indianapolis, Indiana; 183 - Henry Ford Health System, Detroit, Michigan; 184 - Cardiology, Beth Israel Deaconess Medical Center, Cambridge, Massachusetts; 185 - Medicine, University of Pittsburgh, Pittsburgh, Pennsylvania; 186 - Department of Epidemiology, University of Michigan, Ann Arbor, Michigan; 187 -

Department of Population and Quantitative Health Sciences, Case Western Reserve University, Cleveland, Ohio

#### Supplementary Note 9: FinnGen consortium members

Aarno Palotie[1,2], Mark Daly[1], Bridget Riley-Gills[3], Howard Jacob[3], Coralie Viollet[4,5], Slavé Petrovski[4], Chia-Yen Chen[6], Sally John[6], George Okafo[7], Robert Plenge[8], Joseph Maranville[8], Mark McCarthy[9], Rion Pendergrass[9], Jonathan Davitte[10], Kirsi Auro[11], Simonne Longerich[12], Anders Mälarstig [13], Anna Vlahiotis[13], Katherine Klinger[14], Clement Chatelain[14], Jorg Blankenstein[14], Karol Estrada[15], Robert Graham[15], Dawn Waterworth[16], Chris OÂ´Donnell[17], Nicole Renaud[17], Tomi P. Mäkelä[18], Jaakko Kaprio[2], Minna Ruddock[19], Petri Virolainen[20], Antti Hakanen[20], Terhi Kilpi[21], Markus Perola[21], Jukka Partanen[22], Taneli Raivio[23], Jani Tikkanen[24], Raisa Serpi[24], Kati Kristiansson[21,25], Veli-Matti Kosma[26], Jari Laukkanen[27,28], Marco Hautalahti[29,30], Outi Tuovila[31], Jeffrey Waring[3], Bridget Riley-Gillis[3], Fedik Rahimov[3], Ioanna Tachmazidou[4], Zhihao Ding[7], Marc Jung[7], Hanati Tuoken[7], Shameek Biswas[8], Neha Raghavan[12], Adriana Huertas-Vazquez[12], Jae-Hoon Sul[12], Xinli Hu[13], Åsa Hedman[13], Ma´en Obeidat[17], Jonathan Chung[17], Jonas Zierer[17], Mari Niemi[17], Samuli Ripatti[2], Johanna Schleutker[32,33], Mikko Arvas[22], Olli CarpÃ©n[23,34], Reetta Hinttala[24], Johannes Kettunen[24,35], Arto Mannermaa[26], Katriina Aalto- Setälä[36], Mika Kähönen[25,37], Johanna Mäkelä[29,30], Reetta Kälviäinen[38], Valtteri Julkunen[38], Hilkka Soininen[38], Anne Remes[35], Mikko Hiltunen[39], Jukka Peltola[37], Minna Raivio[34], Pentti Tienari[34], Juha Rinne[33], Roosa Kallionpää[33], Juulia Partanen[40], Adam Ziemann[3], Nizar Smaoui[3], Anne Lehtonen[3], Susan Eaton[6], Heiko Runz[6], Sanni Lahdenperä[6], Natalie Bowers[9], Edmond Teng[9], Fanli Xu[41], David Pulford[42], Laura Addis[41], John Eicher[41], Qingqin S Li[43], Karen He[16], Ekaterina Khramtsova[16], Martti Färkkilä[34], Jukka Koskela[34], Sampsa Pikkarainen[34], Airi Jussila[37], Katri Kaukinen[37], Timo Blomster[35], Mikko Kiviniemi[38], Markku Voutilainen[33], Tim Lu[9], Linda McCarthy[41], Amy Hart[16], Meijian Guan[16], Jason Miller[12], Kirsi Kalpala[13], Melissa Miller[13], Kari Eklund[34], Antti PalomÃ¤ki[33], Pia Isomäki[37], Laura Pirilä[33], Oili Kaipiainen-Seppänen[38], Johanna Huhtakangas[35], Nina Mars[2], Apinya Lertratanakul[3], Marla Hochfeld[8], Jorge Esparza Gordillo[41], Fabiana Farias[12], Nan Bing[13], Tarja Laitinen[2,37], Margit Pelkonen[38], Paula Kauppi[34], Hannu Kankaanranta[44], Terttu Harju[35], Riitta Lahesmaa[33], Hubert Chen[9], Joanna Betts[41], Rajashree Mishra[41], Majd Mouded[45], Debby Ngo[45], Teemu Niiranen[46], Felix Vaura[46], Veikko Salomaa[46], Kaj Metsärinne[33], Jenni Aittokallio[33], Jussi Hernesniemi[37], Daniel Gordin[34], Juha Sinisalo[34], Marja-Riitta Taskinen[34], Tiinamaija Tuomi[34], Timo Hiltunen[34], Amanda Elliott[47], Mary Pat Reeve[2], Sanni Ruotsalainen[2], Dirk Paul[4], Audrey Chu[41], Dermot Reilly[48], Mike Mendelson[49], Jaakko Parkkinen[13], Tuomo Meretoja[50], Heikki Joensuu[51], Johanna Mattson[34], Eveliina Salminen[34], Annika Auranen[52], Peeter Karihtala[51], Päivi Auvinen[38],

Klaus Elenius[33], Esa PitkÃ¤nen[2], Relja Popovic[3], Margarete Fabre[5], Jennifer Schutzman[9], Diptee Kulkarni[41], Alessandro Porello[16], Andrey Loboda[12], Heli Lehtonen[13], Stefan McDonough[13], Sauli Vuoti[53], Kai Kaarniranta[54], Joni A Turunen[55], Terhi Ollila[34], Hannu Uusitalo[37], Juha Karjalainen[2], Mengzhen Liu[3], Stephanie Loomis[6], Erich Strauss[9], Hao Chen[9], Kaisa Tasanen[35], Laura Huilaja[35], Katariina Hannula-Jouppi[34], Teea Salmi[37], Sirkku Peltonen[33], Leena Koulu[33], David Choy[9], Ying Wu[13], Pirkko Pussinen[34], Aino Salminen[34], Tuula Salo[34], David Rice[34], Pekka Nieminen[34], Ulla Palotie[34], Maria Siponen[38], Liisa Suominen[38], Päivi Mäntylä[38], Ulvi Gursoy[33], Vuokko Anttonen[35], Kirsi Sipilä[56], Hannele Laivuori[2], Venla Kurra[37], Laura Kotaniemi-Talonen[37], Oskari Heikinheimo[34], Ilkka Kalliala[34], Lauri Aaltonen[34], Varpu Jokimaa[33], Marja Vääräsmäki[35], Outi Uimari[35], Laure Morin-Papunen[35], Maarit Maarit[35], Terhi Piltonen[35], Katja Kivinen[2], Elisabeth Widen[2], Taru Tukiainen[2], Niko Välimäki[57], Eija Laakkonen[58], Jaakko Tyrmi[59], Heidi Silven[60], Eeva Sliz[60], Riikka Arffman[60], Susanna Savukoski[60], Triin Laisk[61], Natalia Pujol[61], Janet Kumar[10], Iiris Hovatta[62], Erkki Isometsä[34], Hanna Ollila[2], Jaana Suvisaari[46], Antti Mäkitie[63], Argyro Bizaki-Vallaskangas[37], Sanna Toppila-Salmi[64], Tytti Willberg[33], Elmo Saarentaus[2], Antti Aarnisalo[34], Elisa Rahikkala[35], Kristiina Aittomäki [65], Fredrik Åberg[66], Mitja Kurki[67], Aki Havulinna[68], Juha Mehtonen[2], Priit Palta[2], Shabbeer Hassan[2], Pietro Della Briotta Parolo[2], Wei Zhou[69], Mutaamba Maasha[69], Susanna Lemmelä[2], Manuel Rivas[70], Aoxing Liu[2], Arto Lehisto[2], Andrea Ganna[2], Vincent Llorens[2], Henrike Heyne[2], Joel Rämö[2], Satu Strausz[2], Tuula Palotie[71], Kimmo Palin[57], Javier Garcia-Tabuenca[72], Harri Siirtola[72], Tuomo Kiiskinen[2], Jiwoo Lee[67], Kristin Tsuo[67], Kati HyvÃ¤rinen[73], Jarmo Ritari[73], Katri Pylkäs [60], Minna Karjalainen[60], Tuomo Mantere[24], Eeva Kangasniemi[25], Sami Heikkinen[39], Nina Pitkänen[20], Samuel Lessard[14], Clément Chatelain[14], Lila Kallio[20], Tiina Wahlfors[21], Eero Punkka[23], Sanna Siltanen[25], Tiina Jokela[27], Anu Jalanko[2], Auli Toivola[2,21], Huei-Yi Shen[2], Risto Kajanne[2], Rodos Rodosthenous[2], Mervi Aavikko[2], Helen Cooper[2], Denise Öller[2], Rasko Leinonen[74], Henna Palin[25], Malla-Maria Linna[23], Masahiro Kanai[69], Zhili Zheng[69], L. Elisa Lahtela[2], Mari Kaunisto[2], Elina Kilpeläinen[2], Tianduanyi Wang[2], Timo P. Sipilä[2], Oluwaseun Alexander Dada[2], Awaisa Ghazal[2], Anastasia Kytölä[2], Rigbe Weldatsadik[2], Jaska Uimonen[2], Kati Donner[2], Anu Loukola[23], PÃ¤ivi Laiho[21], Tuuli Sistonen[21], Essi Kaiharju[21], Markku Laukkanen[21], Elina Järvensivu[21], Sini Lähteenmäki[21], Lotta Männikkö[21], Regis Wong[21], Minna Brunfeldt[21], Sami Koskelainen[21], Tero Hiekkalinna[21], Teemu Paajanen[21], Shuang Luo[2], Shanmukha Sampath Padmanabhuni[2], Marianna Niemi[72], Javier Gracia-Tabuenca[72], Mika Helminen[72], Tiina Luukkaala[72], Iida Vähätalo[72], Iina Laak[2], Saija Haapa-Paananen[30], Sarah Smith[30], Tom Southerington[30], Meri Lähteenmäki[30]

1 - Institute for Molecular Medicine Finland (FIMM), HiLIFE, University of Helsinki, Helsinki, Finland; Broad Institute of MIT and Harvard; Massachusetts General Hospital; 2 - Institute for Molecular

Medicine Finland (FIMM), HiLIFE, University of Helsinki, Helsinki, Finland; 3 - Abbvie, Chicago, IL, United States; 4 - Astra Zeneca, Cambridge, United Kingdom; 5 - AstraZeneca, Cambridge, United Kingdom; 6 - Biogen, Cambridge, MA, United States; 7 - Boehringer Ingelheim, Ingelheim am Rhein, Germany; 8 - Bristol Myers Squibb, New York, NY, United States; 9 - Genentech, San Francisco, CA, United States; 10 - GlaxoSmithKline, Collegeville, PA, United States; 11 - GlaxoSmithKline, Espoo, Finland; 12 - Merck, Kenilworth, NJ, United States; 13 - Pfizer, New York, NY, United States; 14 - Translational Sciences, Sanofi R&D, Framingham, MA, USA; 15 - Maze Therapeutics, San Francisco, CA, United States; 16 - Janssen Research & Development, LLC, Spring House, PA, United States; 17 - Novartis Institutes for BioMedical Research, Cambridge, MA, United States; 18 - HiLIFE, University of Helsinki, Finland, Finland; 19 - Arctic biobank / University of Oulu; 20 - Auria Biobank / University of Turku / Hospital District of Southwest Finland, Turku, Finland; 21 - THL Biobank / Finnish Institute for Health and Welfare (THL), Helsinki, Finland; 22 - Finnish Red Cross Blood Service / Finnish Hematology Registry and Clinical Biobank, Helsinki, Finland; 23 - Helsinki Biobank / Helsinki University and Hospital District of Helsinki and Uusimaa, Helsinki; 24 - Northern Finland Biobank Borealis / University of Oulu / Northern Ostrobothnia Hospital District, Oulu, Finland; 25 - Finnish Clinical Biobank Tampere / University of Tampere / Pirkanmaa Hospital District, Tampere, Finland; 26 - Biobank of Eastern Finland / University of Eastern Finland / Northern Savo Hospital District, Kuopio, Finland; 27 - Central Finland Biobank / University of Jyväskylä / Central Finland Health Care District, Jyväskylä, Finland; 28 - Central Finland Health Care District, Jyväskylä, Finland; 29 - FINBB - Finnish biobank cooperative; 30 - Finnish Biobank Cooperative - FINBB; 31 - Business Finland, Helsinki, Finland; 32 - Auria Biobank / Univ. of Turku / Hospital District of Southwest Finland, Turku, Finland; 33 - Hospital District of Southwest Finland, Turku, Finland; 34 - Hospital District of Helsinki and Uusimaa, Helsinki, Finland; 35 - Northern Ostrobothnia Hospital District, Oulu, Finland; 36 - Faculty of Medicine and Health Technology, Tampere University, Tampere, Finland; 37 - Pirkanmaa Hospital District, Tampere, Finland; 38 - Northern Savo Hospital District, Kuopio, Finland; 39 - University of Eastern Finland, Kuopio, Finland; 40 - Institute for Molecular Medicine Finland, HiLIFE, University of Helsinki, Finland; 41 - GlaxoSmithKline, Brentford, United Kingdom; 42 - GlaxoSmithKline, Stevenage, United Kingdom; 43 - Janssen Research & Development, LLC, Titusville, NJ 08560, United States; 44 - University of Gothenburg, Gothenburg, Sweden/ Seinäjoki Central Hospital, Seinäjoki, Finland/ Tampere University, Tampere, Finland; 45 - Novartis, Basel, Switzerland; 46 - Finnish Institute for Health and Welfare (THL), Helsinki, Finland; 47 - Institute for Molecular Medicine Finland (FIMM), HiLIFE, University of Helsinki, Helsinki, Finland; Broad Institute, Cambridge, MA, USA and Massachusetts General Hospital, Boston, MA, USA; 48 - Janssen Research & Development, LLC, Boston, MA, United

States; 49 - Novartis, Boston, MA, United States; 50 - Department of Breast Surgery, Helsinki University Hospital Comprehensive Cancer Center and University of Helsinki, Helsinki, Finland; 51 - Department of Oncology, Helsinki University Hospital Comprehensive Cancer Center and University of Helsinki, Helsinki, Finland; 52 - Pirkanmaa Hospital District , Tampere, Finland; 53 - Janssen-Cilag Oy, Espoo, Finland; 54 - Northern Savo Hospital District, Kuopio, Finland; Department of Molecular Genetics, University of Lodz, Lodz, Poland; 55 - Helsinki University Hospital and University of Helsinki, Helsinki, Finland; Eye Genetics Group, Folkhälsan Research Center, Helsinki, Finland; 56 - Research Unit of Oral Health Sciences Faculty of Medicine, University of Oulu, Oulu, Finland; Medical Research Center, Oulu, Oulu University Hospital and University of Oulu, Oulu, Finland; 57 - University of Helsinki, Helsinki, Finland; 58 - University of Jyväskylä, Jyväskylä, Finland; 59 - University of Oulu, Oulu, Finland / University of Tampere, Tampere, Finland; 60 - University of Oulu, Oulu, Finland; 61 - Estonian biobank, Tartu, Estonia; 62 - University of Helsinki, Finland; 63 - Department of Otorhinolaryngology - Head and Neck Surgery, University of Helsinki and Helsinki University Hospital, Helsinki, Finland; 64 - University of Eastern Finland and Kuopio University Hospital, Department of Otorhinolaryngology, Kuopio, Finland and Department of Allergy, Helsinki University Hospital and University of Helsinki, Finland; 65 - Department of Medical Genetics, Helsinki University Central Hospital, Helsinki, Finland; 66 - Transplantation and Liver Surgery Clinic, Helsinki University Hospital, Helsinki University, Helsinki, Finland; 67 - Institute for Molecular Medicine Finland (FIMM), HiLIFE, University of Helsinki, Helsinki, Finland; Broad Institute, Cambridge, MA, United States; 68 - Institute for Molecular Medicine Finland (FIMM), HiLIFE, University of Helsinki, Helsinki, Finland; Finnish Institute for Health and Welfare (THL), Helsinki, Finland; 69 - Broad Institute, Cambridge, MA, United States; 70 - University of Stanford, Stanford, CA, United States; 71 - University of Helsinki and Hospital District of Helsinki and Uusimaa, Helsinki, Finland; 72 - University of Tampere, Tampere, Finland; 73 - Finnish Red Cross Blood Service, Helsinki, Finland; 74 - Institute for Molecular Medicine Finland (FIMM), HiLIFE, University of Helsinki, Helsinki, Finland; European Molecular Biology Laboratory, European Bioinformatics Institute, Cambridge, UK

### References

1. Choi SW, O'Reilly PF. PRSice-2: Polygenic Risk Score software for biobank-scale data. Gigascience. 2019 Jul 1;8(7).

2. Wright JD, Folsom AR, Coresh J, Sharrett AR, Couper D, Wagenknecht LE, et al. The ARIC (atherosclerosis risk in communities) study: JACC focus seminar 3/8. J Am Coll Cardiol. 2021 Jun 15;77(23):2939–59.

3. The ARIC Investigators. The Atherosclerosis Risk in Communities (ARIC) Study: design and objectives. . Am J Epidemiol. 1989 Apr;129(4):687–702.

4. Friedman GD, Cutter GR, Donahue RP, Hughes GH, Hulley SB, Jacobs DR, et al. CARDIA: study design, recruitment, and some characteristics of the examined subjects. J Clin Epidemiol. 1988;41(11):1105–16.

5. Redline S, Tishler PV, Tosteson TD, Williamson J, Kump K, Browner I, et al. The familial aggregation of obstructive sleep apnea. Am J Respir Crit Care Med. 1995 Mar;151(3 Pt 1):682–7.

6. Fried LP, Borhani NO, Enright P, Furberg CD, Gardin JM, Kronmal RA, et al. The Cardiovascular Health Study: design and rationale. Ann Epidemiol. 1991 Feb;1(3):263–76.

7. Regan EA, Hokanson JE, Murphy JR, Make B, Lynch DA, Beaty TH, et al. Genetic epidemiology of COPD (COPDGene) study design. COPD. 2010 Feb;7(1):32–43.

8. Dawber TR, Kannel WB, Lyell LP. An approach to longitudinal studies in a community: the Framingham Study. Ann N Y Acad Sci. 1963 May 22;107:539–56.

9. Splansky GL, Corey D, Yang Q, Atwood LD, Cupples LA, Benjamin EJ, et al. The Third Generation Cohort of the National Heart, Lung, and Blood Institute's Framingham Heart Study: design, recruitment, and initial examination. Am J Epidemiol. 2007 Jun 1;165(11):1328–35.

10. Kannel WB, Feinleib M, McNamara PM, Garrison RJ, Castelli WP. An investigation of coronary heart disease in families. The Framingham offspring study. Am J Epidemiol. 1979 Sep;110(3):281–90.

11. Lavange LM, Kalsbeek WD, Sorlie PD, Avilés-Santa LM, Kaplan RC, Barnhart J, et al. Sample design and cohort selection in the Hispanic Community Health Study/Study of Latinos. Ann Epidemiol. 2010 Aug;20(8):642–9.

12. Sorlie PD, Allison MA, Avilés-Santa ML, Cai J, Daviglus ML, Howard AG, et al. Prevalence of hypertension, awareness, treatment, and control in the Hispanic Community Health Study/Study of Latinos. Am J Hypertens. 2014 Jun;27(6):793–800.

13. Pirzada A, Cai J, Heiss G, Sotres-Alvarez D, Gallo LC, Youngblood ME, et al. Evolving science on cardiovascular disease among hispanic/latino adults: JACC international. J Am Coll Cardiol. 2023 Apr 18;81(15):1505–20.

14. Wyatt SB, Diekelmann N, Henderson F, Andrew ME, Billingsley G, Felder SH, et al. A community-driven model of research participation: the Jackson Heart Study Participant Recruitment and Retention Study. Ethn Dis. 2003;13(4):438–55.

15. Taylor HA, Wilson JG, Jones DW, Sarpong DF, Srinivasan A, Garrison RJ, et al. Toward resolution of cardiovascular health disparities in African Americans: design and methods of the Jackson Heart Study. Ethn Dis. 2005;15(4 Suppl 6):S6-4.

16. Bild DE, Bluemke DA, Burke GL, Detrano R, Diez Roux AV, Folsom AR, et al. Multi-Ethnic Study of Atherosclerosis: objectives and design. Am J Epidemiol. 2002 Nov 1;156(9):871–81.

17. Taliun D, Harris DN, Kessler MD, Carlson J, Szpiech ZA, Torres R, et al. Sequencing of 53,831 diverse genomes from the NHLBI TOPMed Program. Nature. 2021;590(7845):290–9.

18. Keenan BT, Kirchner HL, Veatch OJ, Borthwick KM, Davenport VA, Feemster JC, et al. Multisite validation of a simple electronic health record algorithm for identifying diagnosed obstructive sleep apnea. J Clin Sleep Med. 2020 Feb 15;16(2):175–83.

19. Kurniansyah N, Goodman MO, Khan AT, Wang J, Feofanova E, Bis JC, et al. Evaluating the use of blood pressure polygenic risk scores across race/ethnic background groups. Nat Commun. 2023 Jun 2;14(1):3202.

20. Kogan E, Twyman K, Heap J, Milentijevic D, Lin JH, Alberts M. Assessing stroke severity using electronic health record data: a machine learning approach. BMC Med Inform Decis Mak. 2020 Jan 8;20(1):8.

21. Khera AV, Chaffin M, Aragam KG, Haas ME, Roselli C, Choi SH, et al. Genome-wide polygenic scores for common diseases identify individuals with risk equivalent to monogenic mutations. Nat Genet. 2018 Sep;50(9):1219–24.

22. Ni J, Friedman H, Boyd BC, McGurn A, Babinski P, Markossian T, et al. Early antibiotic exposure and development of asthma and allergic rhinitis in childhood. BMC Pediatr. 2019 Jul 5;19(1):225.

23. Gothe H, Rajsic S, Vukicevic D, Schoenfelder T, Jahn B, Geiger-Gritsch S, et al. Algorithms to identify COPD in health systems with and without access to ICD coding: a systematic review. BMC Health Serv Res. 2019 Oct 22;19(1):737.

24. Jalal K, Anand EJ, Venuto R, Eberle J, Arora P. Can billing codes accurately identify rapidly progressing stage 3 and stage 4 chronic kidney disease patients: a diagnostic test study. BMC Nephrol. 2019 Jul 12;20(1):260.

25. Goyal P, Bose B, Creber RM, Krishnan U, Yang M, Brady J, et al. Performance of electronic health record diagnosis codes for ambulatory heart failure encounters. J Card Fail. 2020 Dec;26(12):1060–6.

26. Zhang Z, Cao L, Chen R, Zhao Y, Lv L, Xu Z, et al. Electronic healthcare records and external outcome data for hospitalized patients with heart failure. Sci Data. 2021 Feb 5;8(1):46.
